# Serological evidence of human infection with SARS-CoV-2: a systematic review and meta-analysis

**DOI:** 10.1101/2020.09.11.20192773

**Authors:** Xinhua Chen, Zhiyuan Chen, Andrew S. Azman, Xiaowei Deng, Xinghui Chen, Wanying Lu, Zeyao Zhao, Juan Yang, Cecile Viboud, Marco Ajelli, Daniel T. Leung, Hongjie Yu

**Author notes:** These authors are co-first authors contributed equally to this work (Xinhua Chen, Zhiyuan Chen, Andrew S. Azman). These authors are joint senior authors contributed equally to this work (Daniel T. Leung, Hongjie Yu). Corresponding author: Hongjie Yu, Fudan University, School of Public Health, Key Laboratory of Public Health Safety, Ministry of Education, Shanghai 200032, China. **Disclaimer:** The views expressed are those of the authors and do not necessarily represent the institutions with which the authors are affiliated.

## Abstract

**Background:** A rapidly increasing number of serological surveys for anti-SARS-CoV-2 antibodies have been reported worldwide. A synthesis of this large corpus of data is needed.

**Purpose:** To evaluate the quality of serological studies and provide a global picture of seroprevalence across demographic and occupational groups, and to provide guidance for conducting better serosurveys.

**Data sources:** We searched PubMed, Embase, Web of Science, and 4 pre-print servers for English-language papers published from December 1, 2019 to September 25, 2020.

**Study selection:** Serological studies evaluating SARS-CoV-2 seroprevalence in humans.

**Data extraction:** Two investigators independently extracted data from studies.

**Data Synthesis:** Most of 230 serological studies, representing tests in >1,400,000 individuals, identified were of low quality based on a standardized study quality scale. In the 51 studies of higher quality, high-risk healthcare workers had higher seroprevalence of 17.1% (95% CI: 9.9-24.4%), compared to low-risk healthcare workers and general population of 5.4% (0.7-10.1%) and 5.3% (4.2-6.4%). Seroprevalence varied hugely across WHO regions, with lowest seroprevalence of general population in Western Pacific region (1.7%, 0.0-5.0%). Generally, the young (<20 years) and the old (≥65 years) were less likely to be seropositive compared to middle-aged (20-64 years) populations.

Seroprevalence correlated with clinical COVID-19 reports, with pooled average of 7.7 (range: 2.0 to 23.1) serologically-detected-infections per confirmed COVID-19 case.

**Limitations:** Some heterogeneity cannot be well explained quantitatively.

**Conclusions:** The overall quality of seroprevalence studies examined was low. The relatively low seroprevalence among general populations suggest that in most settings, antibody-mediated herd immunity is far from being reached. Given the relatively narrow range of estimates of the ratio of serologically-detected infections to confirmed cases across different locales, reported case counts may help provide insights into the true proportion of the population infected.

**Primary Funding source:** National Science Fund for Distinguished Young Scholars (PROSPERO: CRD42020198253).

## Introduction

The epidemic of coronavirus disease 2019 (COVID-19), caused by severe acute respiratory syndrome coronavirus 2 (SARS-CoV-2), was first reported in Wuhan, Hubei province in December, 2019 and quickly spread globally (1). As of October 13, 2020, more than 37 million COVID-19 cases, including 1,077,799 deaths, had been reported in 235 countries/regions (2, 3). The real number of SARS-CoV-2 infections is undoubtedly much higher than the officially reported cases due to various factors, including the occurrence of asymptomatic infections, variable healthcare-seeking for clinically mild cases, varied testing strategies in different countries, false-negative virologic tests, and incomplete case reporting. Therefore, the reported COVID-19 cases based on clinical identification with virologic confirmation only represents the “tip of iceberg”, with a large number of asymptomatic and mild infections in the general population who may only be identified by seroepidemiological studies (4).

Serological studies are a useful tool to estimate the proportion of the population previously infected, to quantify the magnitude of transmission of pathogens, estimate the infection fatality rate (5), assess the impact of interventions (6), and when correlates of protection are available, estimate the degree of population immunity (7, 8). Insights from serological surveillance can be valuable for policymakers and health officials when planning public health decision-making.

A large number of serological investigations across the world have been published during the first 9 months of the COVID-19 pandemic, with highly variable estimates of seroprevalence that may largely be due to differences in attack rates, but which also feature heterogeneous sampling strategies and assays used. Two rapid systematic reviews of SARS-CoV-2 seroprevalence were identified (in preprint at the time of writing) but both had a limited scope and did not explore important differences between subpopulations, assess study quality or perform standard meta-analyses techniques (9, 10).

Here, we conduct a systematic review and meta-analysis to summarize serologic surveys for SARS-CoV-2 infections in humans, to comprehensively evaluate the study designs, laboratory methods, and outcome interpretations for each included serological study, and to estimate the risk of infections by populations with different presumed levels of exposure to SARS-CoV-2. We aim for these results to help inform decision makers and researchers alike as plans are made for the next phases of the global pandemic.

## Methods

### Data Sources and Searches

According to the Preferred Reporting Items for Systematic Reviews and Meta-Analyses (PRISMA) guideline (http://www.prisma-statement.org/) (11), we performed a systematic literature review from three peer-reviewed databases (PubMed, Embase and Web of Science) and four preprint severs (medRxiv, bioRxiv, SSRN and Wellcome) with predefined search terms **(Appendix Table 1)**.

### Study Selection and Extraction

Two independent researchers screened titles and abstracts of English-language papers published from December 1, 2019 to September 25, 2020 meeting the following criteria: (1) a report of seroprevalence in either the general population or some other well-defined population of non-COVID clinical cases; (2) conducted after the first reported case in the area and (3) reported the specific assays used. We excluded studies that only reported serologic responses among COVID-19 patients, those using samples with known infection status (e.g. validation studies of assays), and animal experiments. We excluded abstracts of congress meetings or conference proceedings, study protocols, media news, commentaries, reviews or case reports.

### Data Extraction and Quality Assessment

The full text of included studies after initial screening were scrutinized to assess the overall eligibility based on the inclusion and exclusion criteria by two independent researchers. A third researcher was consulted when the two reviewers disagreed on study assessment. For eligible studies, data were extracted by researchers on the number of participants who provided specimens and the number of these who were seropositive to calculate the seroprevalence. When data were inconsistent between reviewers, they were asked to discuss and revisit the article until reaching a consensus. We developed a scoring system on the basis of a seroepidemiological protocol from Consortium for the Standardization of Influenza Seroepidemiology (CONSISE), a previously published scoring system for seroprevalence studies of zoonotic influenza viruses, and a seroepidemiological protocol developed by World Health Organization (WHO), to further assess the study quality (12-14). We comprehensively assessed study design (representativeness of study participants), laboratory assay (whether internal assay validations or a confirmatory assay was performed) and outcome adjustment (correction for demographics and/or test performance) and based on a scoring scheme, an overall score was determined for each included study **(Appendix Table 6)**. From this score, we classified each study’s quality into one of four grades: A, B, C or D. Grade A, the highest quality category, spanned studies with scores ranging from 12 to 10, grade B from 9 to 7, grade C from 4 to 6, and grade D from 0 to 3. We only included grade A and grade B studies in the main analysis but provide additional results with all studies, irrespective of grade, in the supplement. We also described the characteristics, laboratory testing method, and primary outcome for each available study in **Appendix Table 2-5**.

### Data Synthesis and Analysis

Seroprevalence was defined as the prevalence of SARS-CoV-2-specific antibodies at or above a designated antibody titer to define a seropositive result in each original study. For serial cross-sectional studies, we calculated the sum of the total number of participants who provided specimens and total number of seropositive individuals during the whole study period, to avoid repeated inclusion of the same study. Similarly, only data from the first blood collection were analyzed for studies with a longitudinal design. For studies that used multiple serological assays, we used the seropositive results from the assay with the highest sensitivity and specificity (calculated by Youden’s index). We also conducted a sensitivity analysis with results from the other assays **(Appendix Table 10)**. If a study used a confirmatory assay (e.g. microneutralization assay) to validate the positive and/or equivocal result from the initial screening, we used the results from the confirmatory assay. For those studies reporting multiple isotypes including IgG, we included only IgG in the main analyses as they remain elevated for a longer period post-infection compared to IgM and IgA (15). If seropositivity based only on IgG was not reported separately or seropositivity reported was based only on total antibodies, these results were also included. While many studies did adjust for various factors, we decided to use the crude (unadjusted) estimates in our analyses to ease interpretation across different studies.

To reduce heterogeneity between individual seroprevalence estimates and to provide more meaningful summary statistics, we stratified eligible studies by WHO regions (i.e. African region, Region of Americas, Eastern Mediterranean Region, European Region, South-East Asia Region, Western Pacific Region)(16). To estimate seroprevalence by different types of exposures, within each WHO region, we categorized all study participants into five groups: 1) close contacts, 2) high-risk healthcare workers, 3) low-risk healthcare workers, 4) general populations, and 5) poorly-defined populations **(Appendix Table 5)**. The poorly-defined population classification represents populations with undefined or unknown exposure to laboratory-confirmed or suspected COVID-19 patients (e.g., blood donors, residual blood samples from labs, patients of other diseases, etc.), as well as study populations that cannot be categorized into the first four study populations due to limited exposure information [e.g., healthcare workers without reporting usage of PPE (personal protective equipment) or COVID-19-related exposures]. Based on a random-effect meta-analysis model, we used inverse variance method to estimate pooled seroprevalence by WHO regions and different sub-populations, combined with the use of Clopper-Pearson method to calculate 95% confidence intervals (17, 18).

For seroprevalence estimates from the general population, we further explored potential determinants affecting the seroprevalence, such as sex, age, and the reported cumulative incidence of COVID-19 cases for the location. We calculated cumulative incidence of confirmed COVID-19 by dividing the number of cases reported in the same target population as the serosurvey two weeks before the serosurvey mid-point at the location, by the estimated population size. Furthermore, we meta-analyzed the number of serologically-detected infections (the number of individuals with positive SARS-CoV-2 serology) per confirmed case (the number of reported cases in the serologic study’s target population) with available epidemiological data included in the articles, the WHO website (19), the COVID-19 dashboard of Johns Hopkins University (2), and local health authorities (20). Unless reported in the article, we used population size estimates from WorldPop (21) or a local statistics bureau (22). Studies were included in the meta-analyses of the ratio of serologically-detected infections per confirmed cases if they reported seroprevalence in the representative general population (non-convenience sample) with population data and confirmed case data available for the same population.

Variability between studies was determined by the heterogeneity tests with Higgins’ I² statistic. We explored the reasons for variations among eligible studies and examined whether prevalence of SARS-CoV-2 specific antibodies varied by study location, study quality, and level of exposure by multivariable meta-regression models. All statistical analyses were done using R (version 3.6.3), with ‘meta’ package to conduct the meta-analysis. For all statistical tests, two tailed P-value less than 0.05 were considered statistically significant.

### Role of the funding source

The funder had no role in study design, data collection, data analysis, data interpretation, or writing of the report. The corresponding author had full access to all the data in the study and had final responsibility for the decision to submit for publication.

## Results

We identified a total of 8,487 studies after systematically searching multiple data sources with 4,763 coming from peer-reviewed databases, 3,710 from preprint severs and 14 reports from governments or health authorities. After removing duplicates and screening titles and abstracts, 1,535 studies reporting serological evidence of SARS-CoV-2 infections were found. Finally, a total of 230 studies involving 1,445,028 participants were included in our meta-analysis after full-text scrutiny **(Figure 1)**, with the majority of studies conducted in European Region (n=106), Region of the Americas (n=69), and Western Pacific Region (n=34) **(Figure 1, Appendix Figure S9)**. Among 214 studies reporting the exact starting sampling date, seventeen (17/214, 8%) were conducted after more than 75% of the total cases (in that country/states/provinces) had been reported as of Sept 25, 2020, most of which (14/17, 82%) were conducted in China **(Appendix Figure 2-7)**. For those seventeen studies, the serosurveys were conducted a median of 46 days (range: 12-102) after the epidemic peak. Similarly, fifty-three studies (53/214, 25%) were conducted after more than 50% of total cases had been reported, with a median of 40 (range: 2-113) days after the peak. General populations (55/230, 24%) and healthcare workers (both high-risk and low-risk healthcare workers) (78/230, 34%) were sampled with the highest frequency. The age profile of participants varied across studies of different targeted populations **(Appendix Table 2)**. Most studies involved participants aged 18-69 years (222/230 studies) and only seven studies exclusively recruited young people (<18 years), including two papers focusing on children (<10 years), and one exclusively targeting older (>80 years) persons from nursing home **(Appendix Table 2)**.

**Figure 1.**
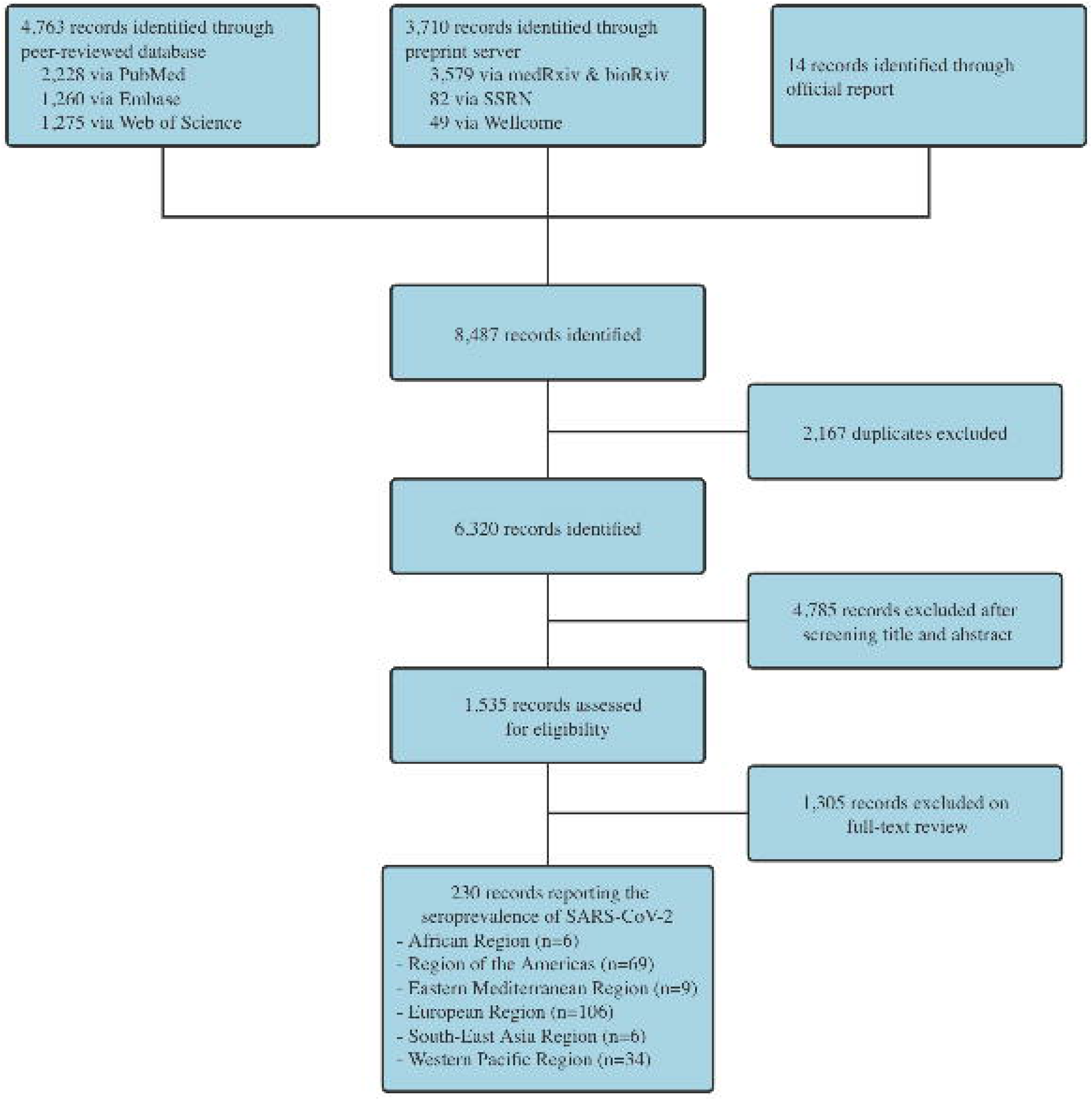
Flowchart of the selection of serological studies of SARS-CoV-2 infection, December 2019-September 2020

The overall quality of studies was low based on our grading system **(Appendix Table 6)**. The majority of studies were categorized as Grades C or D **(Appendix Figure 1)**, including all but one study of high-risk healthcare workers, and all studies of close contacts. Of the 55 general-population-based studies, 5 achieved Grade A, and 19 achieved Grade B.

Approximately two-third of studies (149/230, 65%) described serological results from convenience samples, while only thirty studies (30/230, 13%) used multi-stage/stratified random sampling to select study participants. Most studies measured IgG antibodies using chemiluminescence immunoassays (CLIA, 92/230, 40%), followed by enzyme-linked immunosorbent assays (ELISA, 80/230, 35%), and lateral flow immunoassays (LFIA, 59/230, 26%), with 19% (43/230) of studies using more than one serologic assay. Additionally, nineteen studies utilized a microneutralization assay to detect neutralizing antibodies. Among 165 studies that provided the targeted protein for serological assays, 123 (75%) studies used tests targeting the S-protein, and 104 (63%) studies used tests targeting the N-protein. More than half of the studies (126/230, 55%) reported age/sex-specific seroprevalence or corrected their findings for age and/or sex. Thirty-one studies (31/230, 13%) adjusted for sensitivity and specificity of the serological assay(s). Only twenty-eight studies corrected for both demographic factors and test performance.

Among 51 Grade-A and Grade-B studies, seroprevalence varied across WHO regions and study populations. Generally, high-risk healthcare workers had a higher seroprevalence (17.1%, 95% CI: 9.9-24.4%), compared to low-risk healthcare workers (5.4%, 95% CI: 0.7-10.1%) and general populations (5.3%, 95% CI: 4.2-6.4%) **(Figure 3A, Appendix Table 9)**. The seroprevalence of the populations in studies that did not specify exposure was 4.1% (95% CI: 3.0-5.1%) **(Figure 3A, Appendix Table 9)**. Estimates of seroprevalence from the general population was highest in the one study conducted in the African Region (25.4%, 95% CI:19.1-31.7%), followed by Eastern Mediterranean Region (2 studies, 14.4%, 95% CI:2.2-26.7%), Region of the Americas (9 studies, 5.2%, 95% CI:2.9-7.5%), and European Region (10 studies, 4.2%, 95% CI:2.7-5.8%), with the lowest seroprevalence in studies conducted in the Western Pacific Region (2 studies, 1.7%, 95% CI:0.0-5.0%) **(Figure 3B-G, Appendix Table 9)**. Sensitivity analysis for studies using multiple assays showed that few changes were observed in seroprevalence by different WHO regions and populations compared to our main analysis **(Appendix Table 10)**.

**Figure 2.**
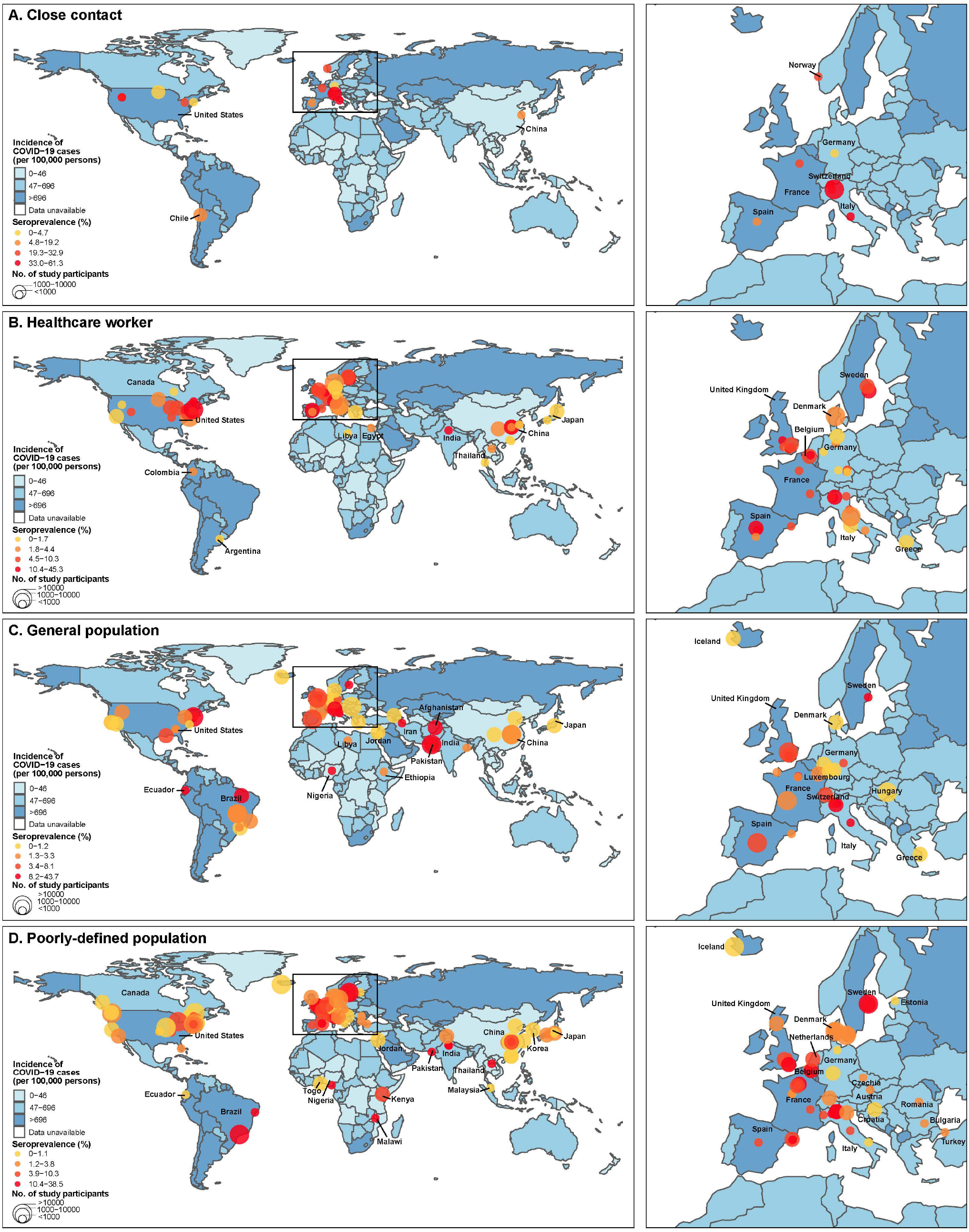
Geographical distribution of SARS-CoV-2 serosurveys in humans by study populations, December 2019-September 2020. The color of the map indicates the cumulative incidence of reported cases (19) with darker colors representing higher values. **(A)** Close contact. **(B)** Healthcare workers. **(C)** General population. **(D)** Poorly-defined population

**Figure 3.**
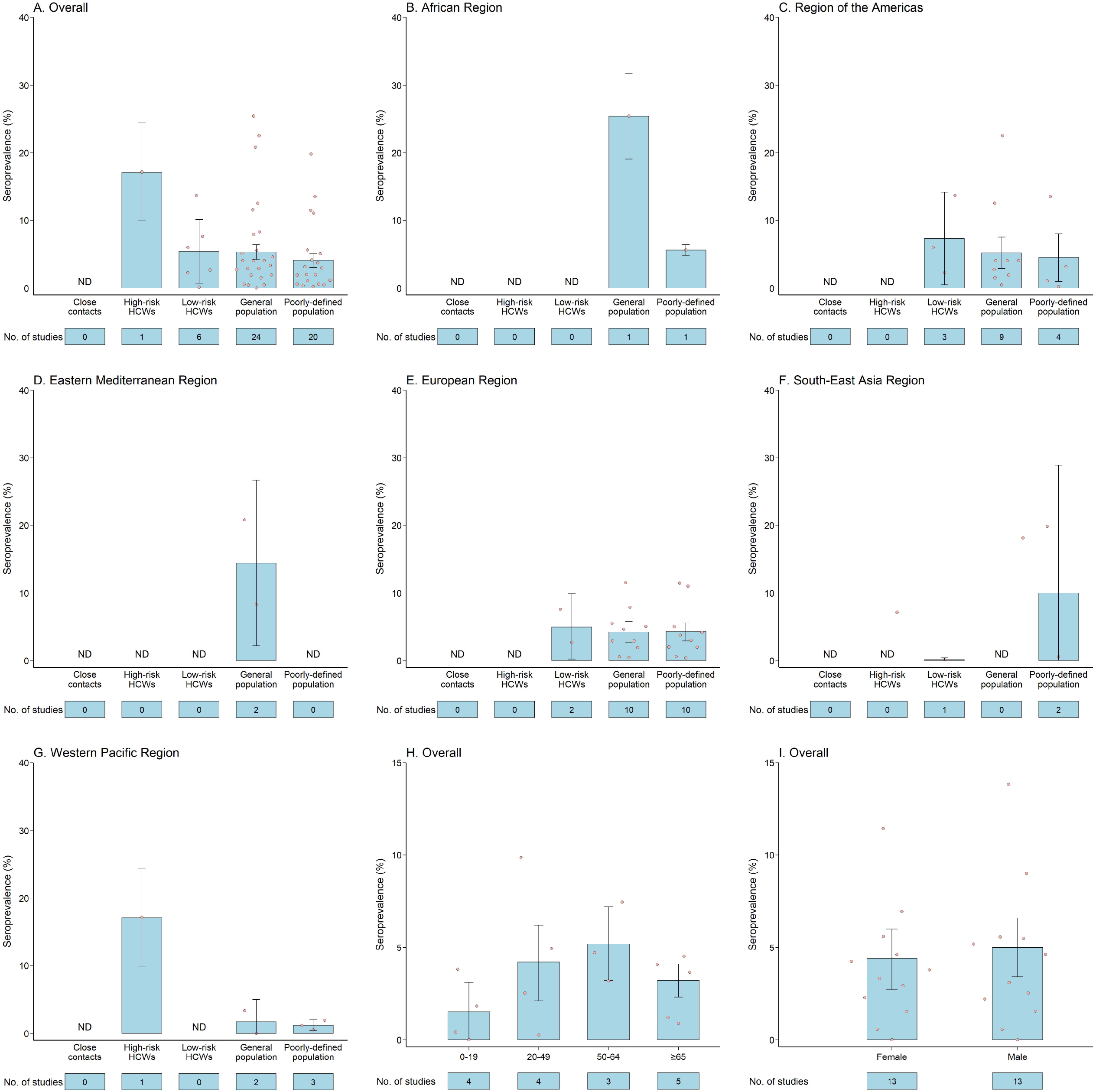
Estimated seroprevalence by WHO regions and study populations. **(A)** Overall estimated seroprevalence by study populations; **(B)** Estimated seroprevalence in African region; **(C)** Estimated seroprevalence in Region of the Americas; **(D)** Estimated seroprevalence in Eastern Mediterranean region; **(E)** Estimated seroprevalence in European region; **(F)** Estimated seroprevalence in South-East Asia region; **(G)** Estimated seroprevalence in Western Pacific region; **(H)** Overall estimated seroprevalence by age groups; **(I)** Overall estimated seroprevalence by sexes (one study were not shown due to the y-axis limit); The bar represents the pooled estimates. The vertical line represents the 95%CI. Each dot represents the result of one single study.

The seroprevalence increased with age for study participants under 65 years, constituting 1.5% (95% CI: 0.0-3.1%), 4.2% (95% CI: 2.1-6.2%) and 5.2% (95% CI: 3.2-7.2%) for study participants younger than 20 years, 20-49 years, and 50-64 years among the 51 studies graded as A or B **(Figure 3H)**. A similar tendency was also observed when analyzing by WHO regions **(Appendix Figure 10B-C)**. After further (imperfectly) merging age groups, the relative risk of seropositivity in the young (<20 years) was approximately 20% lower than that of the middle-age group (20-64 years) (RR=0.76, 95%CI: 0.69-0.85), suggesting young individuals may be less susceptible to SARS-CoV-2 infections or may mount a less robust antibody response **(Appendix Table 12)**. The risk of seropositivity in the old (≥ 65 years) was also lower compared to the middle-age group (RR=0.71, 95%CI: 0.54-0.94) **(Appendix Table 12)**.

Pooled seroprevalence of males (5.0%, 95% CI: 3.4-6.6%) and females (4.4%, 95% CI: 2.7-6.0%) were similar, with 58% (7/12) of studies reporting sex-specific seroprevalence having higher point estimates of seroprevalence in males than females (Figure 3I). Similarly, pooling sex-specific relative risks across studies to adjust for the differences in risk across settings revealed no significant increase in risk of seropositivity in males (RR=1.03, 95% CI: 0.95-1.13) **(Appendix Table 12)**, with similar estimates across WHO regions (**Appendix Table 12**).

The relationship between reported COVID-19 incidence (confirmed cases reported from public sources) and the number of infections identified through serologic surveys can be useful for trying to understand the evolution of the pandemic without serologic surveillance in each and every locale. For studies of the general population, the cumulative reported incidence of COVID-19 correlated with seroprevalence across locations (Pearson correlation coefficient: 0.80), though this is largely driven by one study **(Appendix Figure 11)**. For studies including individuals from general populations, the ratio of serologically-detected infections to virologically-confirmed cases varied across locations, with a pooled ratio of 7.7 (95% CI: 5.5-10.9), suggesting that for each virologically confirmed SARS-COV-2 infection, at least 6 infections remained undetected by surveillance systems. This ratio was similar between the Region of the Americas (7.9, 95% CI: 1.7-35.3) and the European Region (7.7, 95%CI:5.8-10.2) **(Figure 4)**.

**Figure 4.**
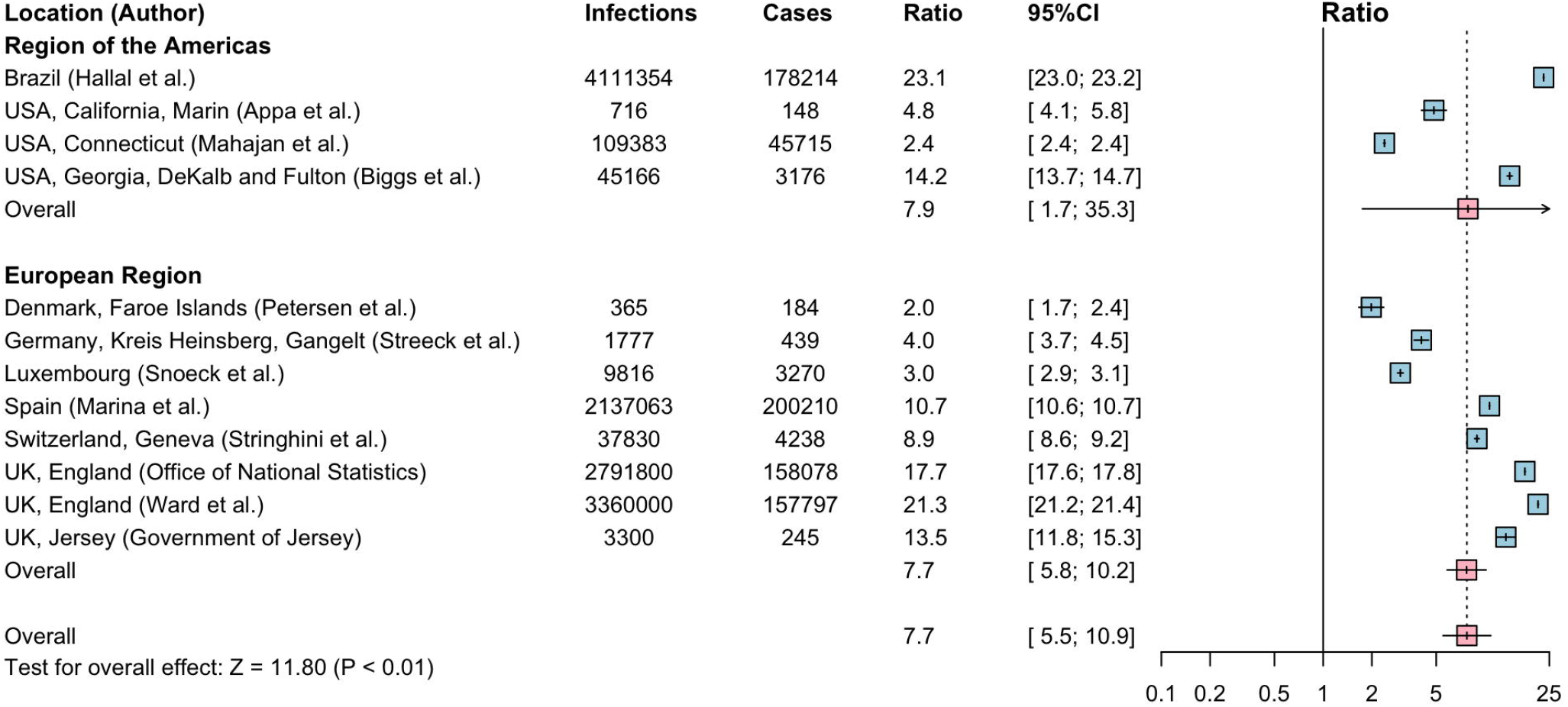
Estimated ratio of human infections with SARS-CoV-2 per COVID-19 confirmed cases. The size of boxes represents the weight for each study. The whisker represents the 95%CI.

## Discussion

With the increasing availability of serological assays for SARS-CoV-2, a large body of literature describing seroprevalence studies in different populations has also emerged. In this study, we examine the quality and results of over two hundred reports of seroprevalence studies from around the globe, both published, and in preprint form. In general, the quality of existing serological studies was low, involving less rigorous sampling strategies, poorly-validated and non-standardized laboratory methods, and lack of statistical correction in analyses. As expected, we found that high-risk healthcare workers had higher prevalence of SARS-CoV-2 specific antibodies than that of low-risk healthcare workers and the general population. Young individuals (<20 years) and older persons (≥65 years) are less likely to be seropositive than those of middle age (20-64 years), while there was not a significant difference in seroprevalence between males and females. Additionally, we find that the ratio of infections per confirmed case is generally on the same order of magnitude across populations with very different surveillance and healthcare systems.

Representative serosurveys can provide useful snapshots of the infection history of a population. However, very few studies (30/230, 13%) provided representative estimates for their target population. An optimal study design for estimating seroprevalence includes a detailed sampling framework, rigorous sampling methods (i.e. multi-stage, stratified sampling), and adjustments for selection bias and assay performance (12).

Various detection assays were used for determination of seropositivity. We found large variations in test performance, targeted antigens and immunoglobulin isotypes, and threshold used. Furthermore, more than half of the studies lacked independent validations of the sensitivity and specificity of the diagnostic kits prior to assessment of serosurvey samples to verify their initial results (23-25). Notably, although the WHO established a generic population-based serological study protocol, there is a lack of standardized guidelines and procedures for laboratory testing, which may contribute towards such heterogeneity in both performance and reporting of results. We call on national and international governance bodies to develop standardized antibody testing protocols and reporting practices and create biobanks of reference standards (e.g. monoclonal antibodies), to reduce lab-to-lab variations, thus facilitating the comparability and interpretability amongst seroprevalence studies. Despite the WHO recommendations, the estimates described by many of the population-based serosurveys did not adjust for demographic structure of the target population (14), nor the testing performance (sensitivity and specificity) of the assay, which made the comparison among studies difficult.

Most of the high-quality serological surveys identified were conducted in the Region of the Americas and the European Region, predominantly on general populations. There were a very limited number of high-quality studies of exposed populations, especially for healthcare workers and close contacts, and studies to address this knowledge gap are needed. For the other four WHO regions examined, there was a paucity of high-quality studies across all populations examined, suggesting that attention should be paid to optimize the design of future sero-epidemiological studies to include good representativeness of samples, standardized laboratory methods, and reasonable adjustments. Higher quality studies provide more accurate measures of disease burden and transmission, to better inform public health efforts against COVID-19.

We found high seroprevalence among high-risk healthcare workers, defined as those who provided routine medical care to COVID-19 patients without PPE. On the contrary, low-risk healthcare workers, defined as those wearing adequate PPE or those who provided care for non-COVID-19 patients, had seroprevalence estimates closer to (and with overlapping confidence intervals) that of the general population (26). We found a pooled seroprevalence of 5.3% in the general population, suggesting that globally, the number of persons infected thus far is unlikely to approach that satisfying estimates of what it would take to achieve antibody-mediated herd immunity (27-29). The seroprevalence in the general population also varied across WHO regions, indicating different levels of community transmission of SARS-CoV-2 in different locales. In addition, the very limited number of high-quality studies in all but two of the WHO regions underlines our incomplete understanding of COVID-19 burden in much of the world. (30) We also found notable differences in seroprevalence between age groups, with the seroprevalence increasing with age among participants younger than 65 years. We found that the young (<20 years) were less likely to be seropositive compared to middle-aged (20-64 years) individuals, consistent with reports of lower numbers of virally-confirmed cases in children compared to other age groups (31). In some areas, this may have been due to the effects of lockdowns, limiting exposures of school-aged populations, in contrast to adult-aged essential workers who had continued community exposure (32, 33). The prevalence of SARS-CoV-2 specific antibodies and relative risk of being seropositive among older persons (≥65 years) was also low, which may be explained by poorer serologic responses after infection (34), lower rates of infection as a result of biological differences or, perhaps more likely, due to behavior changes leading to reduced frequency of infectious contacts.

High heterogeneity was observed for all study populations. Although meta regression could partly explain such varied seroprevalence between different study populations with different type of exposures, the internal heterogeneity for the same population should be interpreted carefully. The seroprevalence of the general population is likely a reflection of the duration and intensity of community transmissions. We showed using a regression analysis that higher cumulative incidence of reported cases is associated with higher seroprevalence. For locations where data on the number of confirmed cases were available, we found that the number of infections per confirmed case varied (2.0-23.1) but the order of magnitude was quite consistent across locales. Such evidence indicates the existence of a large number of undetected cases in the community and provides a range of scaling factors for translating the observed confirmed cases into unobserved infections in the community.

Our study has several limitations. First, although we have performed meta-regression and subgroup analysis to explore heterogeneity of varied seroprevalence for different populations, there are still some factors that we have not taken into account, so that some heterogeneity cannot be well explained quantitatively. Second, misclassification bias may occur due to the limited information on exposures for the study populations, especially for the “Poorly-defined” populations. For some key information (e.g., the use of PPE for healthcare workers) that cannot be extracted from the original articles, we have tried to contact the authors, but the response rate was low. Third, our findings involving cumulative incidence of confirmed cases are limited by the inherently heterogeneous incidence across locations, due to the diversity of testing strategy, case tracing and identification, and public health interventions. Fourth, only raw data without correcting for assay performance or sampling design were used in the analysis to ensure comparability within studies. Fifth, the seroprevalence of IgG was possibly overestimated due to the inability to isolate IgG-based seroprevalence from total antibodies-based seroprevalence in some studies. Lastly, our infection-to-case estimates did not account for age-specific differences in exposure risk and symptom severity.

In conclusion, the overall quality of the existing seroprevalence studies of SARS-CoV-2 is low and international efforts to standardize study design and assays are urgently needed (14). Pooled estimates of SARS-CoV-2 seroprevalence based on currently available data demonstrate a higher infection risk among close contacts and healthcare workers lacking PPE, while the relatively low prevalence of SARS-CoV-2 specific antibodies among general populations suggests that the majority of examined populations have not been infected. Therefore, antibody-mediated herd immunity is likely far from being reached in most settings, and continuous serological monitoring is necessary to inform public health decision making.

## Data Availability

All data referred to in the manuscript was available in appendix.

## Contributors

H.Y. designed and supervised the study. X.C. and Z.C. did the literature search, set up the database and did all statistical analyses. X.C., Z.C., A.S.A., and D.T.L. co-drafted the first version of the article. X.C, Z.C., X.D., W.L, Z.Z, and X.C. helped with checking data and did the figures. D.T.L., A.S.A., J.Y, M.A., C.V. and H.Y. commented on the data and its interpretation, revised the content critically. All authors contributed to review and revision and approved the final manuscript as submitted and agree to be accountable for all aspects of the work.

## Declaration of interests

H.Y. has received investigator-initiated research funding from Sanofi Pasteur, GlaxoSmithKline, and Yichang HEC Changjiang Pharmaceutical Company; M.A. has received research funding from Seqirus; D.T.L. and A.S.A. has received research funding from the US National Institutes of Health. None of those research funding is related to COVID-19. All other authors report no competing interests.

## Acknowledgments

We thank Wei Wang, Qianhui Wu, Yongli Zhang, Junbo Chen, Qianli Wang, and Yuxia Liang from the Fudan University. This study was funded by the National Science Fund for Distinguished Young Scholars (grant no. 81525023), Program of Shanghai Academic/Technology Research Leader (grant no. 18XD1400300), National Science and Technology Major project of China (grant no. 2018ZX10713001-007, 2017ZX10103009-005, 2018ZX10201001-010), the US National Institutes of Health (R01 AI135115 to D.T.L. and A.S.A.)

## Notes

### Author Declarations

This review was preregistered with the protocol available in the PROSPERO database (ID: CRD42020198253).

