## Supplementary material for "Serological evidence of human infection with SARS-CoV-2: a systematic review and meta-analysis": SI

#### Methods and results

*Data extraction and variable list*

We screened all eligible studies to determine: 1) study characteristics, study population and related types/levels of exposure to SARS-CoV-2; 2) antibody detection assays used; 3) predefined outcomes, i.e. SARS-CoV-2 antibody seroprevalence

The following variables were extracted from qualified studies, including the author's name, publication date, study design, sampling period, study period, study population and location, age and occupation of participants, exposure setting, frequency and type of exposure, the use of personal protective equipment (PPE) for healthcare workers, laboratory methodology for serologic confirmation of SARS-CoV-2 infections (including assay methods, the manufacturer and related agency authorization, targeted immunoglobulin and antigens, days from exposure to sampling, experiment validations, sensitivity and specificity of the validated assay, cross-reactivity with other coronaviruses, seropositive threshold value, confirmatory assay and definitions of serological infections for each study), assessment of participants' symptom (including the number of symptomatic and asymptomatic serological infections), and predefined outcomes (i.e. the total number of participants, the number of participants provided single or paired sera, the number of seropositive participants, adjustments of the results and potential risk factors for serological infections).

*Rationale for modifying scoring systems for antibody detection assays that focused on human infection with avian influenza SARS-CoV-2 virus*

In consideration of a published sero-epidemiological study protocol by Consortium for the Standardization of Influenza Seroepidemiology (CONSISE)，an established scoring system for serological study concerning animal influenza exposure in humans, and a population-based seroepidemiological investigation protocol for COVID-19 from the WHO, we developed a modified scoring system to develop a more appropriate system to weigh the serological evidence for SARS-CoV-2 infections in humans (12-14). In our scoring system, study design, laboratory assay and outcome adjustment are three main considerations. Studies reporting the method to recruit participants or sampling methods (e.g. convenient sample or randomly-selected samples) receive a higher score. Specifically, studies with detailed sampling framework, or using stratified/multi-stage sampling are assessed the highest points (3 points), followed by those studies with simplified random (2 points) or convenience sampling (1 point). If a study does not report how they recruited their study participants, the study receives zero points.

Multiple serological assays are available to detect SARS-CoV -2 antibodies, with various test performance (different sensitivity and specificity), different targeted antigens, immunoglobulin isotypes, and various positive threshold or cut-off values. It is difficult to compare the performance of different serological assays without a uniform "gold standard". Studies using well-validated (previously evaluated in published paper) in-house serological assays, as well as those using detection kits approved by GPC/WHO-recognized national regulatory authority [e.g. Food and Drug administration (FDA), Conformité Européenne-In Vitro Diagnostics (CE-IVD), and National Medical Products Administration (NMPA)], are assigned 1 point. Additionally, if internal validations (using their own specimens to evaluate sensitivity and specificity) were performed prior to assay of population samples, they are assigned 2 points. Similarly, if a study used confirmatory assays, such as microneutralization assay (2 points) or other serological methods (1 point), to validate their initial screening results, additional points were given. If a study only used microneutralization assay to detect SARS-CoV-2 antibodies, 2 points were assigned to it.

For the outcome analysis, adjustment for the local demographic factors (mainly including age and sex) or test performance is of great importance to interpret the serological results. A total of four points are assigned for studies accounting for these two adjustments at the same time, with 2 points for each adjustment.

*Quality assessment of serological studies*

Based on their overall score, the study quality was further classified into four Grades, A, B, C and D, according to their quartiles. Grade A spanned studies with a scores ranging from 10 to 12, Grade B from 7 to 9, Grade C from 4 to 6, and Grade D from 0 to 3.

*Rationale for assessing asymptomatic and symptomatic SARS-CoV-2 infections*

We evaluated all included studies according to whether a study reported any acute respiratory illness (i.e. fever or respiratory symptoms) among participants during the COVID-19 epidemic. To distinguish symptomatic and asymptomatic serological infections among different populations, we recorded serologically-confirmed number of symptomatic or asymptomatic individuals for studies assessing participants' fever or other COVID-19 related respiratory symptoms. Specifically, seroprevalence of symptomatic ($p_{sym} = \frac{Number\ of\ symptomatic\ infections}{Total\ number\ of\ participants\ provided\ specimens}$) and asymptomatic infections ($p_{asym} = \frac{Number\ of\ asymptomatic\ infections}{Total\ number\ of\ participants\ provided\ specimens}$) were calculated based on the total number of participants who provided specimens. If a study reported participants' non-COVID-19 symptoms or reported symptoms before the start of SARS-CoV-2 epidemic in their country, corresponding symptomatic of asymptomatic serological results would not be included in analysis.

#### Appendix Tables

#### Appendix Table 1. Search strategy for three peer-reviewed databases and four preprint severs

| Database | Step | Searching strategy | Number of articles* |
|---|---|---|---|
| PubMed | #1 | 2019-nCoV OR "coronavirus disease 2019" OR COVID-19 OR "severe acute respiratory syndrome coronavirus 2" OR SARS-CoV-2 | 54,901 |
| | #2 | seroprevalen* OR seroincidenc* OR seroconversion OR seronegative OR seropositive* OR seroepidemiolog* OR serolog* OR serosurvey* OR antibod* OR (infection* AND ("attack rate" OR "cumulative incidence")) | 977,351 |
| | #3 | 2019/12/01-2020/09/25 | 1,298,115 |
| | #4 | Language: English | 26,819,608 |
| | #5 | #1 **AND** #2 **AND** #3 AND #4 | 2,228 |
| Web of Science | #1 | TS = (2019-nCoV OR "coronavirus disease 2019" OR COVID-19 OR "severe acute respiratory syndrome coronavirus 2" OR SARS-CoV-2) | 37,456 |
| | #2 | TS= (seroprevalen* OR seroincidenc* OR seroconversion OR seronegative OR seropositive OR seropositivity OR seroepidemiolog* OR serolog* OR serosurvey* OR antibod* OR (infection* AND ("attack rate" OR "cumulative incidence"))) | 1,050,283 |
| | #3 | 2019/01/01-2020/09/25 | - |
| | #4 | Language: English | - |
| | #5 | #1 **AND** #2 **AND** #3 AND #4 | 1,275 |
| Embase | #1 | 2019-nCoV OR "coronavirus disease 2019" OR COVID-19 OR "severe acute respiratory syndrome coronavirus 2" OR SARS-CoV-2 | 34,636 |

| | | | |
|---|---|---|---|
| | #2 | seroprevalen* OR seroincidenc* OR seroconversion OR seronegative OR seropositive* OR seroepidemiolog* OR serolog* OR serosurvey* OR antibod* OR (infection* AND ("attack rate" OR "cumulative incidence")) | 1,066,142 |
| | #3 | 2019/12/01-2020/09/25 | - |
| | #4 | Language: English | - |
| | #5 | #1 **AND** #2 **AND** #3 AND #4 | 1,260 |
| medRxiv & bioRxiv | #1 | COVID-19 OR SARS-CoV-2 | 9,773 |
| | #2 | sero* OR antibod* | 38,307 |
| | #3 | 2019/12/01-2020/09/25 | 45,309 |
| | #4 | #1 AND #2 AND #3 | 3,579 |
| SSRN* | #1 | COVID-19 AND antibody, COVID-19 AND seroprevalence | 82 |
| Wellcome* | #1 | COVID-19 | 49 |

*Databases do not permit Boolean operator OR, extensive search was done for SSRN and Wellcome.

#### Appendix Table 2. Descriptive characteristics of serological studies included in the systematic review

| Reference | Location (country) | Study period | Study type | Study population | No. of participants | Age of participants (Median, range/mean±SD) | Exposures | Symptom assessment | [illegible] |
|---|---|---|---|---|---|---|---|---|---|
| Peer-reviewed database | | | | | | | | | |
| Victoria et al., 2020 | Washington, USA | Jan 2020 | Longitudinal study | Office co-workers, waiting room contacts, healthcare contacts | 11 office co-workers<br>20 waiting room contacts, and<br>7 healthcare workers | Office co-workers: 39 (24-62) years old;<br>Waiting room contacts: 53.5 (<1-78) years old;<br>Healthcare contacts: 36 (30-56) years old;<br>All contacts: 45 (0-78) years old | Office co-workers: being an office co-worker of the case-patient with close contact of any duration;<br>Waiting room contacts: sharing a healthcare waiting room or area during the same time and up to 2 hours after the case-patient was present;<br>Healthcare contacts: any face-to-face interaction between healthcare personnel and the case-patient without wearing the full personal protective equipment (i.e., gown, gloves, eye protection, and N95 respirator) or potential contact with the case-patients' secretions by HCP without wearing full PPE. | Yes | [illegible] |
| To et al., 2020 | Hongkong, China | Jan 2020 | Cross-sectional study for general population;<br>Longitudinal study for Hong residents evacuated from Hubei | General population;<br>Hongkong residents evacuated from Hubei | 1938 general population (specimens collected from clinical biochemistry laboratory);<br>469 Hongkong residents evacuated from Hubei | General population: 0-80 years old;<br>Hongkong residents evacuated from Hubei: 41 years old | Poorly-defined exposures for both populations | Symptom of Hongkong residents evacuated from Hubei was assessed. | [illegible] |
| Liang et al., 2020 | Guangdong, China; Wuhan, Hubei, China | Jan 2020 | Cross-sectional study | Inpatients and their healthy companions | Guangzhou: 8782;<br>Wuhan: 8272 | Guangzhou: 54 (44-62) years old<br>Wuhan: 55 (38-67) years old | Poorly-defined exposures | Yes (The seropositive individuals had no history of COVID-19 symptoms, and therefore regarded as asymptomatic or mild) | [illegible] |

| Reference | Location (country) | Study period | Study type | Study population | No. of participants | Age of participants (Median, range/mean±SD) | Exposures | Symptom assessment | Se se |
|---|---|---|---|---|---|---|---|---|---|
| Hallowell et al., 2020 | USA | Feb 2020 | Cross-sectional study | Evacuees from Wuhan in a repatriation | 193 | 42 (0–74) years old | Among participants with serological results: 1 person had close contact with laboratory-confirmed COVID-19 case-patient in past 2 months; 30 had close contact with person with fever and/or acute respiratory illness in past 2 month | Yes (9/193 of evacuees reported having experienced signs or symptoms associated with COVID-19 in the previous 2 weeks, and 24/193 of evacuees reported signs/symptoms associated with COVID-19 in the previous 2 months) | Ye |
| Sam et al., 2020 | Kuala Lumpur and Selangor state, Malaysia | Jan 2020 | Cross-sectional study | Residual serum samples collected at a teaching hospital | 588 | All ages | Residual serum with poorly-defined exposures | No | Ye |
| Chen et al., 2020 | Nanjing, China | Feb 2020 | Cross-sectional study | Healthcare worker | 105 | 30.0 (26.0-39.5) years old | Direct contact with four COVID-19 patients | Yes (12/105 of participants reported general symptoms, including fever, headache, sore throat,etc.) | Ye |
| Cavicchiolo et al., 2020 | Veneto, Italy | Feb 2020 | Cross-sectional study | Neonates | 75 | - | - | No | Ye |
| Plebani et al., 2020 | Padova, Italy | Feb 2020 | Cross-sectional study | Healthcare workers | 8285 | 43.2±11.6 years old | - | No | Ye |
| Cox et al., 2020 | Bergen, Norway | Feb 2020 | Cross-sectional study | Household members of confirmed COVID-19 cases | 77 | - | Household contact | No | Ye |

| Reference | Location (country) | Study period | Study type | Study population | No. of participants | Age of participants (Median, range/mean±SD) | Exposures | Symptom assessment | Se se |
|---|---|---|---|---|---|---|---|---|---|
| Brandstetter et al., 2020 | Regensburg, Germany | Mar 2020 | Cross-sectional study | Hospital staff | 180 hospital staff with different levels of exposures | 18-65 years old | Close contact: unprotected contact with a distance of less than 2 meters for 15 minutes or longer;<br>Moderate contact: contact with a distance of less than 2 meters while using personal protective equipment or unprotected contact with a distance of more than 2 meters;<br>No contact: not aware of any contact to a COVID-19 patient. | Yes | Ye H H |
| Solodky et al., 2020 | Lyon, France | Mar 2020 | Cross-sectional study | Healthcare worker;<br>cancer patients | 244<br>85 | - | Poorly-defined exposures | Healthcare worker: Yes<br>Cancer patients: No | Ye pa |
| Zhang et al., 2020 | Guangdong, China | Mar 2020 | Cross-sectional study | Healthy individuals returning to Shenzhen | 1589 | 36.4 (11–89) years old | - | Yes (All were asymptomatic) | Ye |
| Suda et al., 2020 | Japan | Mar 2020 | Cross-sectional study | Outpatients with liver disease | 700 | 20-84 years old | Poorly-defined exposure | Yes (All were asymptomatic) | Ye |
| Bogogiannidou et al., 2020 | Greece | Mar 2020 | Cross-sectional study | Leftover blood samples from nationwide labs | 6586 | All ages | Poorly-defined exposure with residual blood samples | No | Ye |
| Xu et al., 2020 | Hubei, Guangdong, China | Mar 2020 | Cross-sectional study | Hemodialysis Patients;<br>Healthcare worker | Hemodialysis Patients: 1542<br>Healthcare worker: 3205 | - | - | Yes | Ye he |
| Vena et al., 2020 | Liguria and Lombardia, Italy | Mar 2020 | Cross-sectional study | non-hospitalized participants in an outpatient setting. | 3609 | Median (IQR): 51 (41–63) years old | - | Yes | Ye |
| Ng et al., 2020 | San Francisco Bay Area, USA | Mar 2020 | Cross-sectional study | Blood donors;<br>Hospitalized patients admitted for non-respiratory indications | Blood donors: 1000;<br>Hospitalized patients admitted for non-respiratory indications: 387 | - | - | Yes | Ye pa in |
| Dingens et al., 2020 | Seattle, USA | Mar 2020 | Cross-sectional study | Residual serum samples from Seattle Children's Hospital | 1076 | - | - | Yes | Ye |
| Fischer et al., 2020 | North Rhine-Westphalia, Lower-Saxony, Hesse, German | Mar 2020 | Cross-sectional study | Blood donors | 3186 | 18-65 years old | - | Yes | Ye |

| Reference | Location (country) | Study period | Study type | Study population | No. of participants | Age of participants (Median, range/mean±SD) | Exposures | Symptom assessment | [illegible] |
|---|---|---|---|---|---|---|---|---|---|
| Brown et al., 2020 | USA | Mar 2020 | Cross-sectional study | Student who contacted with infected teacher | 21 | 17 (5-18) years old | Interactive classroom contact (mean in-class time was 108 minutes); noninteractive classroom contact (mean in-class time was 50 minutes). | Yes | [illegible] |
| Han et al., 2020 | Wuhan, Hubei, China | Mar 2020 | Cross-sectional study | Persons during work resumption screening | 22633 | - | - | Yes (All were asymptomatic) | [illegible] |
| De et al., 2020 | Belgium | Mar 2020 | Longitudinal study | Hemodialysis patients | 282 | - | - | Yes (All seropositive individuals are symptomatic) | [illegible] |
| Zhou et al., 2020 | Wuhan, Hubei, China | Mar 2020 | Cross-sectional study | Hospital staff | 3674 | Older than 18 years old | - | Yes | [illegible] |
| Tu et al., 2020 | Wuhan, Hubei, China | Mar 2020 | Cohort study | Pediatric medical workers | 325 | - | Contact with confirmed and/or suspected cases of COVID-19 | No | [illegible] |
| Fuereder et al., 2020 | Vienna, Austria | Mar 2020 | Cross-sectional study | Healthcare professionals; cancer patients | Healthcare professionals: 62; cancer patients: 84 | 41 (23–59) years old; median: 61 years old | - | Yes | [illegible] |
| Fusco et al., 2020 | Naples, Italy | Mar 2020 | Cross-sectional study | Healthcare worker | 120 | Median (IQR): 43 (32-51.5) years old | Direct contact with patient or patients' environment | Yes | [illegible] |
| Havers et al., 2020 | USA | Mar 2020 | Cross-sectional study | Residual patient sera collected for routine screening | 3264 (Washington)<br>2482 (New York City)<br>1184 (Louisiana)<br>1742 (South Florida)<br>824 (Pennsylvania)<br>1882 (Missouri)<br>1132 (Utah)<br>1224 (California)<br>1431 (Connecticut)<br>860 (Minnesota) | All ages | - | No | [illegible] |
| Xu et al., 2020 | Guangdong, China | Mar 2020 | Cross-sectional study | Blood donors | 2199 | 34 (18-59) years old | - | No | [illegible] |
| Behrens et al., 2020 | Hannover, Germany | Mar 2020 | Longitudinal study | First line health care professional | 217 | Mean (range): 36.5 (18-63) years old | Direct contact with a confirmed SARS-CoV-2 infected person | Yes | [illegible] |

| Reference | Location (country) | Study period | Study type | Study population | No. of participants | Age of participants (Median, range/mean±SD) | Exposures | Symptom assessment | Se se |
|---|---|---|---|---|---|---|---|---|---|
| Loconsole et al., 2020 | Bari, Italy | Mar 2020 | Cross-sectional study | Patients admitted to Emergency Department | 819 | Median (IQR): 66 (52–80) years old | - | Yes | Ye |
| Mansour et al., 2020 | New York, USA | Mar 2020 | Cross-sectional study | Healthcare worker | 285 | 18-84 years old | Exposure to aerosolized SARS-CoV-2 or direct patient exposure (emergency medicine, critical care, anesthesiology; direct contact with patients) | Yes | Ye |
| Gallian et al., 2020 | France | Mar 2020 | Cross-sectional study | Bloor donors | 998 | Median: 41 years old | Poorly-defined exposure | Yes (no history of fever or symptom of respiratory infection in the previous 2 weeks) | Ye |
| Korth et al., 2020 | Essen, Germany | Mar 2020 | Cross-sectional study | High-risk healthcare worker; intermediated-risk healthcare worker;<br>low-risk healthcare worker | 244 high-risk healthcare workers; 37 intermediated-risk healthcare workers and 35 low-risk healthcare workers | High-risk healthcare worker: 36.7±10.7 years old intermediated-risk healthcare worker: unk low-risk healthcare worker: 42.3±3.2 years old | High-risk healthcare: daily COVID-19 patient contact; intermediated-risk healthcare worker: daily non-COVID-19 patient contact;<br>low-risk healthcare worker: without daily patient contact; | Yes | Ye in 3! ) |
| Bielecki et al., 2020 | Switzerland | Mar 2020 | Cross-sectional study | Two soldier cohorts at a Swiss Army Base | Company 1: 154<br>Company 2 and 3: 354 | Company 1: 20.4 (18-27) years old<br>Company 2: 18-28 years old | Company 1: without any COVID-19 cases; Company 2 and 3: heavily affected by COVID-19 | Yes | Ye Co |
| Tsaneva et al., 2020 | Varna, Bulgaria | Mar 2020 | Cross-sectional study | Outpatients | 586 | 3-92 years old | Poorly-defined exposure | Yes | Ye |
| Houlihan et al., 2020 | London, UK | Mar 2020 | Longitudinal study | First-line healthcare worker | 200 | Median (IQR): 34 (29-44) years old | Contact with COVID-19 patients | Yes | Ye |
| Basteiro et al., 2020 | Barcelona, Spain | Mar 2020 | Cross-sectional study | Health care workers | 578 | 43.8±11.1 years old | - | Yes | Ye |
| Isherwood et al., 2020 | UK | Mar 2020 | Cross-sectional study | Patients in a tertiary acute general surgical unit;<br>Healthcare staff in the same healthcare setting | Patients in a tertiary acute general surgical unit: 1964<br>Healthcare staff in the same healthcare setting: 215 | Healthcare staff: 20-69 years old | - | No | Ye su th |

| Reference | Location (country) | Study period | Study type | Study population | No. of participants | Age of participants (Median, range/mean±SD) | Exposures | Symptom assessment | [illegible] |
| --- | --- | --- | --- | --- | --- | --- | --- | --- | --- |
| Xu et al., 2020 | Hubei, Chongqing, Sichuan, Guangdong, China | Mar 2020 | Cross-sectional study | Healthcare worker; healthcare worker relative; Hemodialysis patient; Outpatient; Hotel staff member; Community resident; Factory worker | 714 healthcare workers in Wuhan; 3091 healthcare workers in Hubei, 319 healthcare workers in Chongqing; 260 healthcare workers in Guangdong; 219 Healthcare worker relatives in Wuhan; 979 Hemodialysis patients in Hubei; 993 outpatients in Chongqing; 563 Hemodialysis patients in Guangdong; 346 Hotel staff member in Hubei; 9442 Community residents in Sichuan; 442 Factory workers in Guangdong | Healthcare worker in Wuhan: 33 (28, 39) years old; Healthcare worker in Hubei :35 (29, 47) years old; Healthcare worker in Chongqing: 33 (28, 50) years old; Healthcare worker in Guangdong: 32 (27, 40) years old; Healthcare worker relative in Wuhan: 42 (31, 56) years old; Hemodialysis patient in Hubei: 57 (48, 67) years old; Hemodialysis patient in Guangdong:59 (47, 70) years old; Outpatient in Chongqing:52 (36, 64) years old; Hotel staff member in Wuhan: 46 (37, 50) years old; Community resident in Sichuan:56 (40, 69) years old Factory workers in Guangdong: 29 (25, 32) years old | Healthcare workers in Wuhan engaged in COVID-19 patients' management | No | [illegible] |
| Milani et al., 2020 | Milan, Italy | Mar 2020 | Cross-sectional study | Personnel of the University of Milan | 197 | - | Poorly-defined exposure | Yes | [illegible] |
| Bryan et al., 2020 | Idaho, USA | Apr 2020 | Cross-sectional study | Community resident | 4856 residents | All ages | - | No | [illegible] |

| Reference | Location (country) | Study period | Study type | Study population | No. of participants | Age of participants (Median, range/mean±SD) | Exposures | Symptom assessment | Se se |
|---|---|---|---|---|---|---|---|---|---|
| Hains et al., 2020 | Indianapolis, USA | Apr 2020 | Cohort study | Hemodialysis patients; Healthcare worker | 13 hemodialysis patients and 25 healthcare workers | Hemodialysis patients: 13 (2-16) years old Healthcare worker: 40.5 (25-61) years old | Potential contact with a hemodialysis patient diagnosed with COVID-19 in the unit. | Yes | Ye H |
| Liu et al., 2020 | Guangdong, China | Apr 2020 | Cross-sectional study | Healthcare worker deployed to Wuhan, and healthcare professionals at home hospital | 116 Doctors, 304 nurses and 77 control healthcare professionals | Doctors: 42.2 years old; Nurses: 33.4 years old; Control healthcare professionals: 57.8 years old | Caring for patients with critical disease and operating aerosol generating procedure (AGPs),<br>Control healthcare professionals: without exposure to COVID-19 patients at home hospital | Yes | Ye Co pr |
| Malickova et al., 2020 | Czech Republic | Apr 2020 | Cross-sectional study | Inflammatory bowel disease healthcare professionals | 92 | 45 (38-57) years old | Poorly-defined exposure | Yes | Ye |
| Lackermair et al., 2020 | Bavaria, Germany | Apr 2020 | Cross-sectional study | Healthcare worker | 151 | 38 (26–47) years old | - | Yes | Ye |
| Sotgiu et al., 2020 | Milan, Italy | Apr 2020 | Cross-sectional study | Healthcare worker | 202 | Median (IQR): 45 (35-54) years old | Contact with Covid-19 patients | No | Ye |
| Wu et al., 2020 | Wuhan, Hubei, China | Apr 2020 | Cross-sectional study | People applying for a permission of resume;<br>hospitalized patients | 1021 persons applying for a permission of resume;<br>381 hospitalized patients | - | Poorly-defined exposure | Yes | Ye re 38 |
| Stubblefield et al., 2020 | Tennessee, USA | Apr 2020 | Cross-sectional study | Healthcare worker worked in COVID-19 units | 249 | 33 (21–70) years old | Direct contact with COVID-19 patients | Yes | Ye |
| Self et al., 2020 | 12 states, USA | Apr 2020 | Cross-sectional study | Frontline Health care personnel | 3248 | Median: 36 years old | Cared for patients with COVID-19, | Yes | Ye |
| Patel et al., 2020 | Tennessee, USA | Apr 2020 | Longitudinal study | Health care personnel | 249 | 33 (21-70) years old | - | Yes | Ye |
| Flannery et al., 2020 | Pennsylvania, USA | Apr 2020 | Cross-sectional study | Pregnant women presenting for delivery | 1293 | Median (IQR): 31 (27-35) years old | - | No | Ye |

| Reference | Location (country) | Study period | Study type | Study population | No. of participants | Age of participants (Median, range/mean±SD) | Exposures | Symptom assessment | Se se |
|---|---|---|---|---|---|---|---|---|---|
| Stock et al., 2020 | New York, USA | Apr 2020 | Cross-sectional study | Adult clinicians | 98 | 37.6±10.6 years old | - | Yes | Ye |
| Goldberg et al., 2020 | Massachusetts, USA | Apr 2020 | Cross-sectional study | Staff members at a Skilled Nursing Facility;<br>residents at a Skilled Nursing Facility | Staff members at a Skilled Nursing Facility: 97;<br>residents at a Skilled Nursing Facility: 56; | Staff members at a Skilled Nursing Facility: 45 years old<br>residents at a Skilled Nursing Facility: 83 (54-102) years old | Poorly-defined exposure | Yes (All were asymptpmatic) | Ye<br>Fa<br>N |
| Stringhini et al., 2020 | Geneva, Switzerland | Apr 2020 | Cross-sectional study | General population | Week1: 341;<br>Week2: 469;<br>Week3: 577;<br>Week4: 604;<br>Week5: 775;<br>Overall: 2766 | Older than 5 years old | - | Yes | Ye<br>w<br>w |
| Erikstrup et al., 2020 | Denmark | Apr 2020 | Cross-sectional study | Blood donors | 20640 | 17–69 years old | Poorly-defined exposure | Yes (Donors must self-defer for two weeks if they develop fever with upper respiratory symptom) | Ye |
| Lahner et al., 2020 | Rome, Italy | Apr 2020 | Cross-sectional study | Healthcare worker | 2057 | 46 (16–69) years old | Exposure to SARS-CoV-2-positive subjects | Yes | Ye |
| Pallett et al., 2020 | London, UK | Apr 2020 | Cross-sectional study | Healthcare worker | 1704 | Symptomatic healthcare worker (mean): 38.2 years old<br>Asymptomatic healthcare worker (mean): 42.4 years old | Delivered direct clinical care to SARS-CoV-2- positive inpatients in cohort areas or isolation rooms involving aerosol-generating procedures | Yes | Ye |
| Sood et al., 2020 | California, USA | Apr 2020 | Cross-sectional study | General population | 863 | Older than 18 years old | - | Yes | Ye |
| Madsen et al., 2020 | Utah, USA | Apr 2020 | Cross-sectional study | ED employees | 279 | - | - | No | Ye |
| Crovetto et al., 2020 | Barcelona, Spain | Apr 2020 | Cross-sectional study | Pregnant women attending first trimester screening | 874 | - | - | Yes | Ye |

| Reference | Location (country) | Study period | Study type | Study population | No. of participants | Age of participants (Median, range/mean±SD) | Exposures | Symptom assessment | [illegible] |
|---|---|---|---|---|---|---|---|---|---|
| Gudbjartsson et al., 2020 | Iceland | Apr 2020 | Cross-sectional study | Persons contact with the Icelandic health care system for reasons other than Covid-19;<br>Icelanders in the greater Reykjavik area;<br>Residents of Vestmannaeyjar;<br>Icelanders had been quarantined | Persons contact with the Icelandic health care system for reasons other than Covid-19: 18609;<br>Icelanders in the greater Reykjavik area: 4843;<br>Residents of Vestmannaeyjar: 663;<br>Icelanders had been quarantined: 4222 | Persons contact with the Icelandic health care system for reasons other than Covid-19: 56±20 years old;<br>Icelanders in the grea Reykjavik area: 48 ±13 years old;<br>Residents of Vestmannaeyjar: 52 ±18 years old;<br>Icelanders had been quarantined: 47±17 years old | - | Yes | [illegible] |
| Naranbhai et al., 2020 | Massachusetts, USA | Apr 2020 | Cross-sectional study | Asymptomatic residents | 200 | Median (IQR): 46 (27-55) years old | - | Yes | [illegible] |
| Martin et al., 2020 | Brussels, Belgium | Apr 2020 | Longitudinal study | Staff members worked in a tertiary reference hospital for infectious diseases | 532 | 37 (21-66) years old | All staff members worked in the Covid-19 highly exposed units | Yes | [illegible] |
| Amendola et al., 2020 | Milan, Italy | Apr 2020 | Cross-sectional study | Healthcare worker | 663 | Median: 44 years old | - | Yes | [illegible] |
| Iversen et al., 2020 | Denmark | Apr 2020 | Cross-sectional study | Healthcare worker;<br>Blood donors | Healthcare worker: 28792;<br>Blood donors: 4672 | Healthcare worker: 44.4±12.6<br>Blood donors: 40.7±13.4 | - | Yes | [illegible] |
| Olalla et al., 2020 | Marbella, Spain | Apr 2020 | Cross-sectional study | Health care workers | 498 | Mean: 41.5 years old | Contact with CoVID-19 cases inside or outside the workplace | Yes | [illegible] |
| Cosma et al., 2020 | Piedmont, Italy | Apr 2020 | Cross-sectional study | Pregnant women | 138 | 32.6 ± 3.54 for seropositive individuals;<br>33.9 ± 4.63 for seronegative individuals | - | Yes | [illegible] |
| Caban-Martinez et al., 2020 | South Florida, USA | Apr 2020 | Cross-sectional study | Frontline firefighter/paramedic workforce | 203 | Older than 21 years old | - | Yes | [illegible] |

| Reference | Location (country) | Study period | Study type | Study population | No. of participants | Age of participants (Median, range/mean±SD) | Exposures | Symptom assessment | [illegible] |
|---|---|---|---|---|---|---|---|---|---|
| Poletti et al., 2020 | Lombardy, Italy | Apr 2020 | Cross-sectional study | Close contacts of COVID-19 cases | 5484 | Median (IQR): 50 (30–61) years old | Contact with COVID-19 cases | No | [illegible] |
| Racine-Brzostek et al., 2020 | New York, USA | Apr 2020 | Cross-sectional study | Health care workers | 2274 | 37 (31-48) years old | Patient-facing for physicians | Yes | [illegible] |
| Rosenberg et al., 2020 | New York, USA | Apr 2020 | Cross-sectional study | General population | 15626 | Older than 18 years old | - | No | [illegible] |
| Daniel et al., 2020 | USS Theodore Roosevelt aircraft carrier, USA | Apr 2020 | Cross-sectional study | Service member | 382 | 18-59 years old | Service member may contact with the 1000 service members who were previously determined to be infected with SARS-CoV-2 during the period. | Yes | [illegible] |
| Schmidt et al., 2020 | Lower Saxony, Germany | Apr 2020 | Cross-sectional study | Clinic staff | 406 | Older than 18 years old | - | Yes | [illegible] |
| Moscola et al., 2020 | New York, USA | Apr 2020 | Cross-sectional study | Health Care Personnel in the New York City Area | 46117 | Median (IQR): 42 (31.5–34.5) years old | Working in a COVID-19–positive unit | No | [illegible] |
| Montenegro et al., 2020 | Barcelona, Spain | Apr 2020 | Longitudinal study | Community individuals; patients consulting the primary care physician | Community individuals: 311; patients consulting the primary care physician: 743 | Community individuals: 43.7±21.79; patients consulting the primary care physician: 46.97±20.0 years old | - | Yes | [illegible] |
| Steensels et al., 2020 | Belgium | Apr 2020 | Cross-sectional study | Hospital staff | 3056 | 39.5 ± 13.1 for seropositive individuals; 41.3 ± 12.4 for seronegative individuals | Contact with COVID-19 patients | Yes | [illegible] |

| Reference | Location (country) | Study period | Study type | Study population | No. of participants | Age of participants (Median, range/mean±SD) | Exposures | Symptom assessment | Se[illegible] se[illegible] |
|---|---|---|---|---|---|---|---|---|---|
| Soriano et al., 2020 | Madrid, Spain | Apr 2020 | Cross-sectional study | University employees; University employees' relatives; Social services and health care workers; Individuals living in communities; Other people | 175 University employees;<br>85 University employees' relatives;<br>108 Social services and health care workers;<br>234 Individuals living in communities;<br>72 other people | University employees: 44 (31, 67) years old;<br>University employees' relatives: 41 (18, 76) years old;<br>Social services and health care workers: 42 (21, 79) years old;<br>Individuals living in communities: 60 (20, 89) years old;<br>Other: 53 (18, 76) years old | - | Yes | Ye[illegible]<br>U[illegible]<br>Se[illegible]<br>1[illegible]<br>2[illegible] |
| Eyre et al., 2020 | Oxford, UK | Apr 2020 | Cross-sectional study | Health care workers | 10610 | Older than 18 years old | Contact with a confirmed or suspected case. | Yes | Y[illegible] |
| Shields et al., 2020 | Birmingham, UK | Apr 2020 | Cross-sectional study | Health-care workers | 516 | Median (IQR): 42 (30-51) years old | - | Yes | Y[illegible] |
| Menachemi et al., 2020 | Indiana, USA | Apr 2020 | Cross-sectional study | Indiana residents (random sample); Indiana residents (non-random sample) | 3658<br><br>898 | Older than 12 years old | - | No | Y[illegible] |
| Marina et al., 2020 | Spain | Apr 2020 | Cross-sectional study | General population | 61075 | All ages | - | Yes | Y[illegible] |
| Petersen et al., 2020 | Faroe Islands, Denmark | Apr 2020 | Cross-sectional study | Inhabitants of the Faroe Islands | 1500 | 42.1±23.1 years old | - | Yes | Y[illegible] |
| Biggs et al., 2020 | Georgia, USA | Apr 2020 | Cross-sectional study | Community household residents | 696 | All ages | - | Yes | Y[illegible] |
| Sydney et al., 2020 | New York, USA | Apr 2020 | Cross-sectional study | Healthcare workers | 1700 | - | - | Yes | Y[illegible] |
| Hunter et al., 2020 | Indiana, USA | Apr 2020 | Cross-sectional study | Healthcare worker | 734 | Mean: 43 years old | - | No | Y[illegible] |

| Reference | Location (country) | Study period | Study type | Study population | No. of participants | Age of participants (Median, range/mean±SD) | Exposures | Symptom assessment | Se se |
|---|---|---|---|---|---|---|---|---|---|
| Josè et al., 2020 | Foggia, Italy | May 2020 | Cross-sectional study | Healthy blood donors | 904 | 18-65 years old | - | Yes | Y |
| Paderno et al., 2020 | Italy | May 2020 | Cross-sectional study | Healthcare worker in otolaryngology unit | 58 | Mean: 41 years old | Contacts with infected patients in hospital and outside hospital | Yes | Y |
| Merkely et al., 2020 | Hungary | May 2020 | Cross-sectional study | Hungarian population | 10504 | 48.7±18.0 years old | - | Yes | Y |
| Dioscoridi et al., 2020 | Milan, Italy | May 2020 | Cohort study | Family members of healthcare workers; healthcare workers | Family members of healthcare workers: 81; health care workers: 38 | Family members of healthcare workers: unk; healthcare workers: 47±18 years old | Health care workers: working in a COVID-19 hospital; Family members lived in the same house with healthcare workers | Yes | Y w |
| Péré et al., 2020 | Paris, France | May 2020 | Cross-sectional study | Healthcare workers | 3569 | Median: 39.6 years old | - | No | Y |
| Torres et al., 2020 | Santiago, Chile | May 2020 | Cross-sectional study | Students; Staff members | 1029 students and 240 staff members | Students: 10.8±4.1 years old; Staff members: 42.8±10.4 years old | Contact with more than 1 confirmed Covid-19 case | Yes | Y St |
| Poulikakos et al., 2020 | North West England, UK | May 2020 | Cross-sectional study | Healthcare workers | 281 | - | Directly involved in patient care | Yes | Y |
| Veerus et al., 2020 | Estonia | May 2020 | Cross-sectional study | Pregnant women | 433 | 31±5.89 years old | - | No | Y |
| Feehan et al., 2020 | Louisiana, USA | May 2020 | Cross-sectional study | General population | 2640 | Mean: 50.6 years old | - | No | Y |
| Sutton et al., 2020 | Oregon, USA | May 2020 | Cross-sectional study | Patients visiting ambulatory, emergency, or inpatient health care setting | 897 | All ages | - | No | Y |
| Bampoe et al., 2020 | London, UK | May 2020 | Cross-sectional study | Maternity healthcare workers | 200 | Older than 18 years old | Patient-facing | Yes | Y |
| Tong et al., 2020 | Jiangsu, China | May 2020 | Cross-sectional study | Medical staff who went to Wuhan city for support | 222 | 32 (24-58) years old | Directly involved in patient care | Yes | Y |

| Reference | Location (country) | Study period | Study type | Study population | No. of participants | Age of participants (Median, range/mean±SD) | Exposures | Symptom assessment | S[illegible] s[illegible] |
|---|---|---|---|---|---|---|---|---|---|
| Mughal et al., 2020 | New Jersey, USA | May 2020 | Cross-sectional study | Healthcare personnel in the ICU setting. | 134 | Median (IQR) :39.2 (28.0-48.5) years old | Exposed to critically ill COVID-19 patients in ICU unit | Yes (All participants were asymptomatic) | Y[illegible] |
| Zhang et al., 2020 | Jiangsu, China | May 2020 | Cross-sectional study | Close contacts of COVID-19 patients | 284 | - | Contact with COVID-19 patients | Yes | Y[illegible] |
| Akinbami et al., 2020 | Michigan, USA | May 2020 | Cross-sectional study | Healthcare, First Response, and Public Safety Personnel | 16397 | Range: 19-82 years old | - | No | Y[illegible] |
| Kempen et al., 2020 | Addis Ababa, Ethiopia | May 2020 | Cross-sectional study | Resident in Addis Ababa | 99 | Older than 14 years old | - | Yes | Y[illegible] |
| Pagani et al., 2020 | Lombardy, Italy | May 2020 | Cross-sectional study | Population of Castiglione D'Adda | 562 | All ages | Contact with verified case | Yes | Y[illegible] |
| Blairon et al., 2020 | Belgium | May 2020 | Cross-sectional study | Healthcare worker | 1485 | - | 215 workers (14.3%) reported having a function with no contact with patients while 1138 (75.9%) have had regular or occasional contact | Yes | Y[illegible] |
| Noh et al., 2020 | Southwestern Seoul, Korea | May 2020 | Cross-sectional study | Outpatients | 1500 | 0-92 years old | - | No | Y[illegible] |
| Lidström et al., 2020 | North of Stockholm, Sweden | May 2020 | Cross-sectional study | Healthcare staff | 8679 | 18-85 years old | - | Yes (All were asymptomatic) | Y[illegible] |
| Haizler-Cohen et al., 2020 | New York state, USA | May 2020 | Cross-sectional study | Pregnant women | 1671 | - | - | No | Y[illegible] |
| Kassem et al., 2020 | Egypt | June 2020 | Cross-sectional study | Healthcare workers employed in the gastroenterology | 74 | Median: 32 years old | - | Yes | Y[illegible] |
| Dimcheff et al., 2020 | Michigan and Ohio, USA | June 2020 | Cross-sectional study | Employees of a Veterans Affairs Healthcare System | 1476 | Older than 18 years old | An exposure was defined as close contact (within six feet) with an individual with confirmed COVID-19 for greater than 15 minutes with the example being exposed to a family member at home who has had a positive COVID-19 nasal swab. | Yes | Y[illegible] |

| Reference | Location (country) | Study period | Study type | Study population | No. of participants | Age of participants (Median, range/mean±SD) | Exposures | Symptom assessment | Se se |
|---|---|---|---|---|---|---|---|---|---|
| Dodd et al., 2020 | 44 states, USA | June 2020 | Cross-sectional study | Blood donors | 160328 | Older than 16 years old | - | No | Y |
| Lundkvist et al., 2020 | Stockholm, Sweden | June 2020 | Cross-sectional study | Residents in Norra Djurgårdsstadena and Tensta | Residents in Norra Djurgårdsstaden: 123<br>Residents in Tensta: 90 | Mean:<br>Residents in Norra Djurgårdsstaden: 37 years old;<br>Residents in Tensta: 50 years old | - | No | Y<br>1 |
| Younas et al., 2020 | Karachi, Pakistan | June 2020 | Cross-sectional study | Blood donors | 370 | 30.6±6.3 years old | - | No | Y |
| Del Brutto et al., 2020 | Atahualpa, Ecuador | May 2020 | Cross-sectional study | Inhabitants in Atahualpa | 673 | 59.2±12.8 years old | - | Yes | Y |
| Preprint database | | | | | | | | | |
| Sughayer et al., 2020 | Amman, Jordan | Jan 2020 | Cross-sectional study | Healthy blood donors | 746 | 18-63 years old | Poorly-defined exposures | Yes | Y |
| Germain et al., 2020 | France | Jan 2020 | Cross-sectional study | Tissue donors | 144 | Median (IQR): 68 (57-79) years old | - | No | Y |
| Chang et al., 2020 | Hubei, Hebei, Guangdong, China | Jan 2020; | Cross-sectional study | Blood donors | Wuhan, Hubei, China: 17794;<br>Shijiazhuang, Hebei, China: 13540;<br>Shenzhen, Guangdong, China: 6810 | Wuhan, Hubei, China: 33 (IQR 19-47) years old;<br>Shijiazhuang, Hebei, China: 40 (IQR 33-48) years old;<br>Shenzhen, Guangdong, China: 36 (IQR 19-53) years old | Poorly-defined exposures | Yes | Y<br>H<br>C |
| Li et al., 2020 | Shanghai, China | Feb 2020 | Cross-sectional study | Individuals with different ocular diseases | 1331 | Median (IQR): 58 (36-68) years old | - | No | Y |
| Buss et al., 2020 | Manaus, São Paulo, Brazil | Feb 2020 | Cross-sectional study | Blood donor | 13867 | 16-70 years old | - | No | Y |
| Stadlbauer et al., 2020 | New York, USA | Feb 2020 | Cross-sectional study | Patients unrelated to COVID-19 | 3412 | All ages | Poorly-defined exposures | No | Y |

| Reference | Location (country) | Study period | Study type | Study population | No. of participants | Age of participants (Median, range/mean±SD) | Exposures | Symptom assessment | Se se |
|---|---|---|---|---|---|---|---|---|---|
| Xiong et al., 2020 | Wuhan, Hubei, China | Feb 2020 | Longitudinal study | Healthcare workers with intensive exposure to COVID-19 patients | 797 | 31 (23–53) years old | Close contact with COVID-19 patients | Yes (All were asymptomatic) | Y |
| Valenti et al., 2020 | Milan, Italy | Feb 2020 | Cross-sectional study | Blood donors | 789 | 18-70 years old | - | Yes | Y |
| Yu et al., 2020 | Hubei, China | Feb 2020 | Cross-sectional study | Health Care Workers | 1184 | 33 (20-68) years old | Contact with confirmed COVID-19 patient | Yes | Y |
| Liu et al., 2020 | Wuhan, Hubei, China | Feb 2020 | Cross-sectional study | Healthcare providers;<br>general workers;<br>other patients | Healthcare providers: 3832<br>general workers: 19555<br>other patients: 1616 | Mean age:<br>Healthcare providers: 37.1 years old;<br>general workers: 41.6 years old;<br>other patients: 53.3 years old | Most of the healthcare providers were exposed to SARS-CoV-2 during the first few months of the outbreak when use of personal protection equipment was sparse as person-to-person transmission was not suspected; | No | Y ge pa |
| Tubiana et al., 2020 | Paris, France | Mar 2020 | Longitudinal study | Healthcare workers | 154 | Median (IQR): 35 (29.0-46.8) years old | Exposed to COVID-19 index | No | Y |
| Skowronski et al., 2020 | British Columbia, Canada | Mar 2020 | Cross-sectional study | Anonymized residual sera were obtained from patients | Mar 2020: 870;<br>May 2020: 889 | Median: 45 years old | Poorly-defined exposures | No | Y 8 |
| Thompson et al., 2020 | Scottish, UK | Mar 2020 | Cross-sectional study | Blood donors | 1000 | 18-75 years old | - | Yes | Y |
| Dietrich et al., 2020 | Louisiana, USA | Mar 2020 | Cohort study | Children from a Children's Hospital | 812 | Median (IQR): 11 (4–15) years old | - | No | Y |
| Brehm et al., 2020 | Hamburg, Germany | Mar 2020 | Longitudinal study | Health care workers;<br>non-health care workers | Health care workers: 1026;<br>non-health care workers: 227 | Mean (range): 38.4 (16-69) years old | Health care workers: Contact to covid-19 cases | Yes | Y h |
| Tang et al., 2020 | Wuhan, Hubei, China | Mar 2020 | Cross-sectional study | Outpatients in Zhongnan Hospital, Wuhan University | 2952 | All ages | - | Yes | Y |
| Augusto et al., 2020 | London, UK | Mar 2020 | Cross-sectional study | healthcare workers | 400 | 36.7 (10.4) years old | Contact with confirmed COVID-19 patient, Contact with confirmed COVID-19 colleague. | Yes | Y |

| Reference | Location (country) | Study period | Study type | Study population | No. of participants | Age of participants (Median, range/mean±SD) | Exposures | Symptom assessment | [illegible] |
|---|---|---|---|---|---|---|---|---|---|
| Wang et al., 2020 | Anhui, China | Mar 2020 | Cross-sectional study | Healthcare workers deployed to Wuhan;<br>Healthcare workers who remained in Hefei | Healthcare workers deployed to Wuhan: 142<br>Healthcare workers who remained in Hefei: 284 | Over 20 years old | Provided care for patients with COVID-19. | No | [illegible] |
| Ling et al., 2020 | Wuhan, Hubei, China | Mar 2020 | Cross-sectional study | Persons experiencing back-to-work medical examinations | 18721 | 40 (42–50) years old | - | Yes | [illegible] |
| Paradiso et al., 2020 | Bari, Italy | Mar 2020 | Longitudinal study | Healthcare worker | 606 | 47 (20-73) years old | Direct contact with individuals with suspected COVID-19 disease in the last two weeks | Yes | [illegible] |
| Herzog et al., 2020 | Belgium | Mar 2020 | Cross-sectional study | Persons with blood samples collected from clinical lab | Period 1: 3910<br>Period 2: 3397<br>Period 3: 3242<br>Period 4: 2960<br>Period 5: 3023 | Mean: 55 years old<br>Mean: 49 years old<br>-<br>-<br>- | - | No | [illegible] |
| Dopico et al., 2020 | Stockholm, Sweden | Mar 2020 | Cross-sectional study | Blood donor and pregnant women | 1900 | - | - | No | [illegible] |
| Streeck et al., 2020 | Heinsberg, Germany | Mar 2020 | Cross-sectional study | Local inhabitants | 1007 | 53 (1-90) years old | - | Yes | [illegible] |
| Doi et al., 2020 | Kobe, Japan | Mar 2020 | Cross-sectional study | Patients who visited outpatient clinics with blood samples | 1000 | All ages | - | No | [illegible] |
| Tosato et al., 2020 | Italy | Apr 2020 | Cross-sectional study | Healthcare professionals | 133 | 51 (39–55) years old | - | Yes | [illegible] |
| Shakiba et al., 2020 | Guilan, Iran | Apr 2020 | Cross-sectional study | Residents in Guilan | 551 | All ages | - | Yes | [illegible] |
| Bastiani et al., 2020 | Italy | Apr 2020 | Cross-sectional study | Adults who lived in Italy | 472 | Female: 49 ± 15.0 years old<br>Male: 52 ± 14.1 years old | - | Yes | [illegible] |
| Carozzi et al., 2020 | Tuscany, Italy | Apr 2020 | Cross-sectional study | Health care workers | 17098 | - | - | Yes | [illegible] |

| Reference | Location (country) | Study period | Study type | Study population | No. of participants | Age of participants (Median, range/mean±SD) | Exposures | Symptom assessment | [illegible] |
|---|---|---|---|---|---|---|---|---|---|
| Siddiqui et al., 2020 | New Delhi, India | Apr 2020 | Longitudinal study | Staff of a tertiary care hospital; individuals visiting that hospital for COVID-19 testing | Staff of a tertiary care hospital: 448; individuals visiting that hospital for COVID-19 testing: 332 | Older than 18 years old | Contact with symptomatic/suspected person | Yes | [illegible] |
| Kammon et al., 2020 | Alzintan, Libya | Apr 2020 | Cross-sectional study | Community residents; Healthcare workers | 142 community residents; 77 healthcare workers: | All ages | - | Yes | [illegible] |
| Bendavid et al., 2020 | California, USA | Apr 2020 | Cross-sectional study | Local residents | 3439 | All ages | - | Yes | [illegible] |
| Egerup et al., 2020 | Copenhagen, Denmark | Apr 2020 | Cross-sectional study | Parturient women; partners of parturient women; newborns | Parturient women: 1361; partners of parturient women: 1236; newborns: 1342 | - | - | Yes | [illegible] |
| Krähling et al., 2020 | Frankfurt, Germany | Apr 2020 | Cross-sectional study | Employees in the Frankfurt metropolitan area | 1000 | 18-65 years old | - | Yes | [illegible] |
| Nopsopon et al., 2020 | Thailand | Apr 2020 | Cross-sectional study | Hospital staff; patients who needed procedural treatment or operation | Hospital staff: 675<br>patients who needed procedural treatment or operation: 182 | Hospital staff: median (IQR): 36.5 (28–45) years old<br>patients who needed procedural treatment or operation: median (IQR): 37 (25–53) years old | Some of the hospital staff and patients have the history of travel to high risk area and of close contact confirmed case | Yes | [illegible] |
| Leidner et al., 2020 | Oregon, USA | Apr 2020 | Longitudinal study | Healthcare workers | 10019 | 42 (18-82) years old | Direct patient contact, contact with patient biospecimens or patient linens | Yes | [illegible] |
| Fujita et al., 2020 | Kyoto, Japan | Apr 2020 | Cross-sectional study | Healthcare workers | 92 | Older than 20 years old | Treat suspected COVID-19 cases | Yes | [illegible] |
| Psichogiou et al., 2020 | Greece | Apr 2020 | Cross-sectional study | Healthcare workers from two hospitals (Hospital-1 was involved in the care of COVID-19 patients while hospital-2 was not) | Hospital-1 HCWs: 906<br>Hospital-2 HCWs: 589 | Older than 18 years old | First-line health care workers (FL-HCWs), defined as personnel whose activities involve contact with patients. | Yes | [illegible] |
| Thomas et al., 2020 | Minnesota, USA | Apr 2020 | Cross-sectional study | Health Care Workers; Asymptomatic outpatients | 1282<br>2379 | 49 (0.17-93) years old<br>41 (18-73) years old | With confirmed and non-confirmed COVID-19 exposures ≥14 days prior<br>Potential COVID-19 exposures or history of prior symptoms consistent with COVID-19 ≥14 days prior. | Yes | [illegible] |

| **Reference** | **Location (country)** | **Study period** | **Study type** | **Study population** | **No. of participants** | **Age of participants (Median, range/mean±SD)** | **Exposures** | **Symptom assessment** | **[illegible]** |
|---|---|---|---|---|---|---|---|---|---|
| Cohen et al., 2020 | Paris, France | Apr 2020 | Cross-sectional study | Children consulting an ambulatory pediatrician | h | 4.9±3.9 years old | Contact with confirmed/ suspected COVID-19 | Yes | [illegible] |
| Rudberg et al., 2020 | Stockholm, Sweden | Apr 2020 | Cross-sectional study | Healthcare workers | 2149 | 44±12 years old | Exposed to patients infected with covid-19 | Yes | [illegible] |
| Sikora et al., 2020 | UK | Apr 2020 | Cross-sectional study | Cancer centre staff | 161 | Mean: 43 years old | - | No | [illegible] |
| Galán et al., 2020 | Madrid, Spain | Apr 2020 | Cross-sectional study | Healthcare workers | 2919 | 43.8 ±11.1 years old | Direct contact with COVID-19 patients | Yes | [illegible] |
| Frank et al., 2020 | Belgium | Apr 2020 | Cross-sectional study | Residents of nursing home; Staff of nursing home | 130 residents and 112 staff | Mean: 86 years old for nursing home residents - | - | No | [illegible] |
| Garralda et al.., 2020 | Madrid, Spain | Apr 2020 | Cross-sectional study | Health care workers | 2439 | mean (range): 42.1 (18-65) years old | Unsafe contact or exposure to a confirmed case | Yes | [illegible] |
| Snoeck et al., 2020 | Luxembourg | Apr 2020 | Cohort study | General population | 1862 | 47±15 years old and 18–84 years old | - | Yes | [illegible] |
| Comar et al., 2020 | Trieste, Italy | Apr 2020 | Cross-sectional study | Healthcare workers | 727 | 22-77 years old | - | Yes | [illegible] |
| Nisar et al., 2020 | Karachi, Pakistan | Apr 2020 | Cross-sectional study | Households | April: 1000; June: 1004 | All ages | - | No | [illegible] |
| Wang et al., 2020 | Beijing, China | Apr 2020 | Cross-sectional study | Communities residents | 2184 | 42.3±19.5 years old | - | Yes | [illegible] |
| Waterfield et al., 2020 | UK | Apr 2020 | Longitudinal study | Healthy children of healthcare workers | 1007 | 10.1 (2-15) years old | - | Yes | [illegible] |
| Zou et al., 2020 | Atlanta, USA | Apr 2020 | Cross-sectional study | Local residents | 142 | - | - | No | [illegible] |

| Reference | Location (country) | Study period | Study type | Study population | No. of participants | Age of participants (Median, range/mean±SD) | Exposures | Symptom assessment | [illegible] |
|---|---|---|---|---|---|---|---|---|---|
| Nopsopon et al., 2020 | Ranong, Thailand | Apr 2020 | Cross-sectional study | Hospital staff | 844 | 42 (32-50) years old | History of travel to the high-risk area was 2.5%, history of close contact PCR confirmed COVID-19 case was 2.0%, history of close contact suspected case was 38.1%. | Yes | [illegible] |
| McDade et al., 2020 | USA | Apr 2020 | Longitudinal study | Household members of essential workers | 202 | Range: 18-70 years old | - | Yes | [illegible] |
| Appa et al., 2020 | California, USA | Apr 2020 | Cross-sectional study | Residents and county essential workers | 1880 | Older than 4 years old | - | Yes | [illegible] |
| Tönshoff et al., 2020 | Baden-Württemberg, Germany | Apr 2020 | Cross-sectional study | Children and their parents | 4964 | Children: 1-10 years old;<br>Parents: 23-66 years old | - | Yes | [illegible] |
| Jerković et al., 2020 | Croatia | Apr 2020 | Cross-sectional study | Industry workers | 1494 | 46 (18–79) years old | - | Yes | [illegible] |
| Alessandro et al., 2020 | Lombardy, Italy | Apr 2020 | Cross-sectional study | General population<br>Healthcare Workers | 1792<br>2415 | 44±16 years old<br>48±10 years old | Contacts with patients | No | [illegible] |
| Halatoko et al., 2020 | Lomé, Togo | Apr 2020 | Cross-sectional study | Healthcare (doctors, nurses, pharmacy auxiliaries, hospital administrators), air transport, police, road transport (taxi and moto-taxi drivers) and informal (market sellers and craftsmen); air transport, police, road transport (taxi and moto-taxi drivers) and informal (market sellers and craftsmen) | Healthcare (doctors, nurses, pharmacy auxiliaries, hospital administrators), air transport, police, road transport (taxi and moto-taxi drivers) and informal (market sellers and craftsmen): 370;<br>air transport, police, road transport (taxi and moto-taxi drivers) and informal (market sellers and craftsmen): 585 | Median (IQR): 36 (32-43) years old | High probability of being in close contact with travelers or with Covid-19 patients | No | [illegible] |
| Dillner et al., 2020 | Stockholm, Sweden | Apr 2020 | Cross-sectional study | Healthy hospital employees | 14057 | All ages | - | No | [illegible] |
| Aziz et al., 2020 | Bonn, Germany | Apr 2020 | Cross-sectional study | Community residents | 4771 | 30-100 years old | - | No | [illegible] |
| Chamie et al., 2020 | San Francisco, USA | Apr 2020 | Cross-sectional study | All residents (>4 years) and workers in census tract | 3953 | Older than 4 years old | - | Yes | [illegible] |

| Reference | Location (country) | Study period | Study type | Study population | No. of participants | Age of participants (Median, range/mean±SD) | Exposures | Symptom assessment | Se se |
|---|---|---|---|---|---|---|---|---|---|
| Nesbitt et al., 2020 | Rhode Island, USA | Apr 2020 | Cross-sectional study | Blood donor | 2008 | Median: 56 years old | - | No | Y |
| Wells et al., 2020 | London & South-East England, UK | Apr 2020 | Cross-sectional study | Members of the Twins UK cohort | 431 | 48.38±28 years old | - | Yes | Y |
| Fontanet et al., 2020 | Paris, France | Apr 2020 | Cross-sectional study | Pupils, their parents and relatives, and staff of primary schools | 1340 | The pupils: 6-11 years old;<br>Parents: 40 (37-44) years old;<br>Teachers: 47.5 (40-51) years old;<br>Non-teaching staff: 47.5 (32-54) years old | - | Yes | Y |
| Sandri et al., 2020 | Lombardy, Italy | Apr 2020 | Cross-sectional study | Healthcare workers | 3985 | Older than 20 years old | - | Yes | Y |
| Uyoga et al., 2020 | Kenya | Apr 2020 | Cross-sectional study | Blood donors | 3174 | 15-64 years old | - | No | Y |
| Brant et al., 2020 | California, USA | May 2020 | Cross-sectional study | Health care workers | 3013 | 42.62±12.12 years old | - | No | Y |
| Addetia et al., 2020 | Washington, USA | - | Cross-sectional study | Ship's crew | 122 | - | - | No | Y |
| Barallat et al., 2020 | Barcelona, Spain. | May 2020 | Cross-sectional study | Healthcare worker | 7563 | 43.81±12.43 years old | Hospital admitted for COVID was low | Yes | Y |
| Tess et al., 2020 | São Paulo, Brazil | May 2020 | Cross-sectional study | Local inhabitants | 517 | Older than 18 years old | - | Yes | Y |
| Takita et al., 2020 | Tokyo, Japan | May 2020 | Cross-sectional study | Community residents | 1071 | All ages | - | Yes | Y |
| Mattern et al., 2020 | Paris, France | May 2020 | Cross-sectional study | All patients admitted to the delivery room | 272 | Median (IQR): 31 (30.5-37) for seropositive individuals;<br>Median (IQR): 33 (29-36) for seronegative individuals; | - | Yes | Y |

| Reference | Location (country) | Study period | Study type | Study population | No. of participants | Age of participants (Median, range/mean±SD) | Exposures | Symptom assessment | Se se |
|---|---|---|---|---|---|---|---|---|---|
| Carrat et al., 2020 | Ile-de-France, Grand Est, Nouvelle-Aquitaine, France | May 2020 | Cross-sectional study | General adult population | 14628 | - | - | Yes | Y |
| McBride et al., 2020 | New York City, USA | May 2020 | Cross-sectional study | Outpatients coming into the Department of Radiation Oncology | 919 | 62 (6-96) years old | | Yes | Y |
| Ebinger et al., 2020 | California, USA | May 2020 | Cross-sectional study | Health Care Workers | 6062 | All ages | Regular contact with Covid-19 patients; work on a unit housing/caring for Covid-19 patients | Yes | Y |
| Hurk et al., 2020 | Netherlands | May 2020 | Cross-sectional study | Blood donor | 8275 | Range: 18-73 years old | - | Yes | Y |
| Weis et al., 2020 | Jena, Germany | May 2020 | Cross-sectional study | Community residents | 626 | Adult: 58.1 years old, Children: 9.62 years old | - | Yes | Y |
| Rigatti et al., 2020 | USA | May 2020 | Cross-sectional study | Life insurance applicants | 50025 | Median (IQR): 42 (34-54) years old | - | Yes | Y |
| Gomes et al., 2020 | Espírito Santo, Brazil | May 2020 | Cross-sectional study | General population | 4612 | All ages | - | Yes | Y |
| Hallal et al., 2020 | Brazil | Apr 2020 | Cross-sectional study | Community residents | 24995 | All ages | - | No | Y |
| Nakamura et al., 2020 | Iwate, Japan | May 2020 | Cross-sectional study | Healthcare workers | 1000 | 40±11 years old | - | No | Y |
| Jespersen et al., 2020 | Central Denmark Region, Denmark | May 2020 | Cross-sectional study | All healthcare workers and administrative personnel at the hospitals (including the pre-hospital services) and specialist practitioner clinics | 17987 | All ages | - | No | Y |
| Tsertsvadze et al., 2020 | Tbilisi, Georgia | May 2020 | Cross-sectional study | Adult residents of capital city of Tbilisi | 1068 | Older than 18 years old | Contact with suspected or confirmed case, History of international travel | Yes | Y |
| Chibwana et al., 2020 | Blantyre City, Malawi | May 2020 | Cross-sectional study | Health care workers | 500 | 31 (20-64) years old | Involved in clinical work related to COVID-19 | Yes | Y |

| Reference | Location (country) | Study period | Study type | Study population | No. of participants | Age of participants (Median, range/mean±SD) | Exposures | Symptom assessment | Se<br>se |
|---|---|---|---|---|---|---|---|---|---|
| Armann et al., 2020 | Saxony, Germany | May 2020 | Cross-sectional study | Students grade 8–11 and their teachers; | Students and teachers without household contact of COVID-19 patients: 2021<br>Students and teachers with household contact of COVID-19 patients: 24 | Median: 15 years old (students)<br>Median: 51 years old (teachers) | - | No | Ye<br>(g<br>gr<br>24 |
| Hibino et al., 2020 | Tokyo, Japan | May 2020 | Longitudinal study | Healthy volunteers working for a Japanese company | 650 | Range: 19-69 years old | - | No | Ye |
| Wilkins et al., 2020 | Illinois, USA | May 2020 | Cross-sectional study | Healthcare workers | 6714 | 40.6±12.0 years old | - | No | Ye |
| Alkurt et al., 2020 | Istanbul and Kocaeli, Turkey | May 2020 | Cross-sectional study | Healthcare workers | 813 | - | - | Yes | Ye |
| Vassallo et al., 2020 | USA | Jun 2020 | Cross-sectional study | Blood Donors | 189656 | Older than 16 years old | - | No | Ye |
| Melo et al., 2020 | Sergipe, Brazil | Jun 2020 | Cross-sectional study | Healthcare workers | 471 | - | - | No | Ye |
| Favara et al., 2020 | Eastern Region, UK | Jun 2020 | Longitudinal study | Staff involved in treating cancer patients | 434 | 40 (19-66) years old | Working within the oncology department ward or out-patient setting and not primarily within a dedicated SARS-CoV-2 in-patient ward | Yes | Ye |
| Silva et al., 2020 | Buenos Aires, Argentina | Jun 2020 | Cross-sectional study | Healthcare workers from public facilities | 738 | Older than 18 years old | Close contact with a confirmed case of COVID- 19 | Yes | Ye |
| Ray et al., 2020 | New Delhi, India | Jun 2020 | Cross-sectional study | Patients who were admitted to the medicine wards and intensive care unit (ICU) | 212 | 41.2±15.4 years old | - | No | Ye |
| Bardai et al., 2020 | Montreal, Canada | Jun 2020 | Cross-sectional study | Children patients;<br>accompanying persons;<br>hospital employees | Children patients: 39;<br>accompanying persons: 61;<br>hospital employees: 99 | Median (IQR):<br>Children patients: 15.6 (13.4-16.8) years old;<br>accompanying persons: 47.1 (41.4; 50.8) years old;<br>hospital employees: 42.5 (32.5; 52.5) | - | Yes | Ye<br>ac<br>er |

| Reference | Location (country) | Study period | Study type | Study population | No. of participants | Age of participants (Median, range/mean±SD) | Exposures | Symptom assessment | Se se |
|---|---|---|---|---|---|---|---|---|---|
| Mahajan et al., 2020 | Connecticut, USA | Jun 2020 | Cross-sectional study | Community residents | 567 | 50.1±17.2 years old | - | Yes | Ye |
| Nishida et al., 2020 | Osaka Prefecture, Japan | Jun 2020 | Cross-sectional study | Hospital staff | 926 | 40.0±11.8 years old | Direct contact with confirmed or suspected COVID-19 patients | Yes | Ye |
| Nawa et al., 2020 | Tochigi, Japan | Jun 2020 | Cross-sectional study | Households randomly selected from Utsunomiya City's basic resident registry | 2290 | All ages | - | Yes | Ye |
| Qutob et al., 2020 | West Bank, Palestine | May 2020 | Cross-sectional study | Individuals visiting medical laboratories;<br>Palestinian population residing in the West Bank | Individuals visiting medical laboratories: 1136;<br>Palestinian population residing in the West Bank: 1355 | Older than 15 years old | - | No | Ye 1[illegible] in |
| Ulyte et al., 2020 | Zurich, Switzerland | Jun 2020 | Cross-sectional study | School children | 2585 | 6-16 years old | - | No | Ye |
| Asuquo et al., 2020 | Calabar, Nigeria | Jun 2020 | Cross-sectional study | Clinic staff and patients | 66 | Older than 18 years old | - | No | Ye |
| Ward et al., 2020 | England, UK | Jun 2020 | Cross-sectional study | Community adults | 105651 | Older than 18 years old | Contact with confirmed or suspected patient | Yes | Ye |
| Menezes et al., 2020 | Brazil | Jun 2020 | Cross-sectional study | Community residents | 33205 | All ages | - | Yes | Ye |
| Ariza et al., 2020 | Bogotá, Colombia | Jun 2020 | Longitudinal study | medical trainees or medical doctors | 351 | - | - | Yes | Ye |
| Majiya et al., 2020 | Niger State, Nigeria | Jun 2020 | Cross-sectional study | Residents | 185 | All ages | Travel overseas, contact with overseas returnee | Yes | Ye |
| Javed et al., 2020 | Peshawar and Quetta, Pakistan | - | Cross-sectional study | Working population | 24210 | 18-65 years old | - | Yes | Ye |
| Buonsenso et al., 2020 | Rome, Italy | - | Cross-sectional study | Household contacts of index patients | 80 | 0-56 years old | Household contacts of index patients | No | Ye |

| Reference | Location (country) | Study period | Study type | Study population | No. of participants | Age of participants (Median, range/mean±SD) | Exposures | Symptom assessment | Se[illegible] se[illegible] |
|---|---|---|---|---|---|---|---|---|---|
| Khan et al., 2020 | Kashmir, India | Jul 2020 | Cross-sectional study | Hospital visitors | 2923 | Older than 18 years old | - | Yes | Ye[illegible] |
| Satpati et al., 2020 | West Bengal, India | Jul 2020 | Cross-sectional study | Population of Paschim Medinipur District | 458 | All ages | - | Yes | Ye[illegible] |
| Silva et al., 2020 | Maranhão, Brazil | Jul 2020 | Cross-sectional study | General population | 3289 | Older than 1 year old | - | Yes | Ye[illegible] |
| Calife et al., 2020 | Baixada Santista metropolitan area, Brazil | - | Cross-sectional study | Residents | 2342 | 37.78±19.98 years old | - | No | Ye[illegible] |
| Kumar et al., 2020 | Ontario, Canada | - | Cross-sectional study | Health care workers | 996 | 40.8±11.1 years old | Directly looked after COVID patient in the last 2 weeks . | Yes | Ye[illegible] |
| Official report | | | | | | | | | |
| Public Health Ontario, Canada, 2020 | Ontario, Canada | Mar 2020 | Cross-sectional study | Serum or plasma left over after diagnostic testing | Mar 2020: 827;<br>May 2020: 1061;<br>Jun 2020: 7014 | All ages | - | No | Ye[illegible] 1[illegible] |
| Office of National Statistics, UK, 2020 | UK | Apr 2020 | Cohort study | General population | 5248 | Older than 2 years old | - | Yes | Ye[illegible] |
| The Government of Jersey, UK, 2020 | UK | Apr 2020 | Longitudinal study | Adult resident population living in private households in Jersey | Round 1: 855; | Older than 16 years old | - | Yes | Ye[illegible] |
| Canadian Blood Services, 2020 | Canada | May 2020 | Cross-sectional study | Blood donor | 37737 | Older than 17 years old | - | No | Ye[illegible] |
| Ministry of Health, Labour and Welfare, Japan, 2020 | Tokyo, Japan; Osaka, Japan; Miyagi, Japan | Jun 2020 | Cross-sectional study | Residents | Tokyo: 1971;<br>Osaka: 2970;<br>Miyagi: 3009 | - | - | No | Ye[illegible] 2[illegible] |
| Islamic Republic of Afghanistan Ministry of Public Health, Afghanistan, 2020 | Afghanistan | Jul 2020 | Cross-sectional study | General population | 9514 | Mean: 27 years old | - | No | Ye[illegible] |

| Reference | Location (country) | Study period | Study type | Study population | No. of participants | Age of participants (Median, range/mean±SD) | Exposures | Symptom assessment | Se se |
|---|---|---|---|---|---|---|---|---|---|
| Public Health England, 2020 | England, UK | Aug 2020 | Cross-sectional study | Blood donor | 7899 | Older than 17 years old | - | No | Y |
| MedLife, Romania, 2020 | Romania | - | Cross-sectional study | Healthcare workers | 371 | - | Contact with patients: average 25 people per day (two thirds were patients)2020 | - | Y |

Abbreviations: IQR: interquartile range; PPE: personal protective equipment; unk: unknown; CLIA: Chemiluminescent immunoassay

#### Appendix Table 3. Summary of antibody detection assays to identify human infection with SARS-CoV-2 included in systematic review

| Reference | Paired serums | Days from last possible exposure to sampling (median/range) | Assay methods (screening methods/confirmatory methods) | Antibodies measured | Targeted antigen | Test performance (sensitivity, specificity)* | Reported positive cut-off value | Comments |
|---|---|---|---|---|---|---|---|---|
| Peer-reviewed databases | | | | | | | | |
| Victoria et al., 2020 | Yes | First: 14 days<br>Second: ≈6 weeks | ELISA | Total antibodies | SP | Sensitivity: 96.0%<br>Specificity: >99.0% | The cut-off OD value is 0.4;<br>Total anti-SARS-CoV-2 antibody titers >400 was considered to be seropositive. | RT-PCR were also performed for participants; the ELISA were well-validated (1). |
| To et al., 2020 | Yes | 1-13 days for Hongkong residents evacuated from Hubei | ELISA | IgG, | NP, SP | Sensitivity: 57.8%-73.3%;<br>Specificity: 100.0% | The cut-off OD values were 0.610 for anti-nucleoprotein IgG and 0.573 for anti-RBD IgG | RT-PCR were also performed for participants |
| | | | MN | Neutralizing antibodies | - | Sensitivity: 91.1% (95%CI 78.8%-97.5%);<br>Specificity: 100.0% | Titer > 1:20 were considered to be seropositive. | |
| Liang et al., 2020 | No | - | CLIA | IgG, IgM | NP, SP | - | - | - |

| Reference | Paired serums | Days from last possible exposure to sampling (median/range) | Assay methods (screening methods/confirmatory methods) | Antibodies measured | Targeted antigen | Test performance (sensitivity , specificity)* | Reported positive cut-off value | Comments |
|---|---|---|---|---|---|---|---|---|
| | | | LFIA | IgG, IgM | - | - | - | This antibody assay was approved by the Chinese Food and Drug Administration and pended approval by US FDA. |
| Hallowell et al., 2020 | No | Within 14 days | ELISA | IgG, IgM, IgA | SP | - | Any specimens with titers ≥400 were considered positive by ELISA | RT-PCR were also performed for participants; Serum samples that were positive by ELISA were confirmed by microneutralization test. |
| | | | MN | Neutralizing antibodies | - | - | - | |
| Sam et al., 2020 | No | - | ELISA | IgG | SP | Sensitivity: 97.1% Specificity: 88.6 % | - | - |
| | | | MN | Neutralizing antibodies | - | Sensitivity: 100% Specificity: 100 % | - | - |
| Chen et al., 2020 | No | Within 14 days | ELISA | IgG, IgM | NP, SP | Sensitivity: 93.3% Specificity: 100.0 % | The cut off was determined if OD of 1:20 diluted serum was | RT-PCR were also performed for participants |

| Reference | Paired serums | Days from last possible exposure to sampling (median/range) | Assay methods (screening methods/confirmatory methods) | Antibodies measured | Targeted antigen | Test performance (sensitivity , specificity)* | Reported positive cut-off value | Comments |
|---|---|---|---|---|---|---|---|---|
| | | | | | | | above the cut-off values for either IgM or IgG against both RBD and NP protein | |
| | | | MN | Neutralizing antibodies | - | - | - | |
| Cavicchiolo et al., 2020 | No | - | CLIA | IgG, IgM | - | - | - | RT-PCR were also performed for participants |
| Plebani et al., 2020 | No | - | CLIA | IgG | NP, SP | Sensitivity: 73.0% Specificity: 98.0 % | - | - |
| Cox et al., 2020 | No | | ELISA | IgG | SP | - | - | The laboratory method was described before (2). |
| Brandstetter et al., 2020 | No | 15-28 days | ELISA | IgG, IgA | SP | - | The OD ratio >1 was considered positive (The OD was detected at 450 nm and the OD-ratio of the measurement of each sample to the | RT-PCR were also performed for participants |

| Reference | Paired serums | Days from last possible exposure to sampling (median/range) | Assay methods (screening methods/confirmatory methods) | Antibodies measured | Targeted antigen | Test performance (sensitivity , specificity)* | Reported positive cut-off value | Comments |
|---|---|---|---|---|---|---|---|---|
| | | | | | | | supplied calibrator was calculated) | |
| Solodky et al., 2020 | No | 15 days or more | LFIA | IgG | - | - | - | RT-PCR were also performed for participants |
| Zhang et al., 2020 | No | - | ELISA | IgG, IgA, IgM | - | - | The cutoff value of this test was defined by receiver operating characteristic curves | - |
| Suda et al., 2020 | No | - | Immunochromatographic test | IgG | | Specificity: 98.0% | - | - |
| | | | CLIA | IgG | NP | Specificity: 100.0% | cutoff index ≥ 1.0 indicates a positive diagnosis | |
| Bogogiannidou et al., 2020 | No | - | CLIA | IgG | NP | Sensitivity: 84.0% Specificity: 99.7% | - | All positive samples, as well as 100 randomly chosen negative samples were confirmed with ELISA |
| Xu et al., 2020 | No | - | CLIA | IgG | NP, SP | Sensitivity: 83.0% Specificity: 100.0% | An S/CO value of >1.0 for either IgG or IgM | - |

| Reference | Paired serums | Days from last possible exposure to sampling (median/range) | Assay methods (screening methods/confirmatory methods) | Antibodies measured | Targeted antigen | Test performance (sensitivity , specificity)* | Reported positive cut-off value | Comments |
|---|---|---|---|---|---|---|---|---|
| | | | | | | | was regarded as positive. | |
| Vena et al., 2020 | No | - | CLIA, LFIA | IgG, IgM | NP, SP | - | - | - |
| Ng et al., 2020 | No | - | CLIA | IgG | NP | - | - | Seropositive samples were confirmed by MN and chemiluminescent immunoassay. |
| Dingens et al., 2020 | No | - | ELISA | IgG | SP | - | - | Seropositive samples were validated by Abbott CLIA and MN. |
| Fischer et al., 2020 | No | - | ELISA | IgG | SP | - | The OD ratio ≥ 1.1 were considered to be seropositive. | Seropositive results were confirmed using the Architect SARS-CoV-2 IgG targeting the viral nucleocapsid and the LIAISON SARS-CoV-2 S1/S2 IgG assay targeting the SARS-CoV-2 spike protein. |

| Reference | Paired serums | Days from last possible exposure to sampling (median/range) | Assay methods (screening methods/confirmatory methods) | Antibodies measured | Targeted antigen | Test performance (sensitivity , specificity)* | Reported positive cut-off value | Comments |
|---|---|---|---|---|---|---|---|---|
| Brown et al., 2020 | No | 14 days | ELISA | - | SP | - | Antibody titers of >400 was considered to be seropositive | - |
| Han et al., 2020 | No | - | Colloidal gold-based immunochromatographic strip assay | IgG, IgM | - | - | - | RT-PCR were also performed for participants |
| De et al., 2020 | Yes | - | MN | IgG | - | | - | RT-PCR were also performed for participants |
| Zhou et al., 2020 | No | - | CLIA | IgG, IgM | NP, SP | - | IgM or IgG level ≥10.0 AU/ml was designated as positive. | - |
| Tu et al., 2020 | No | - | ELISA | IgG, IgM | NP, SP | - | Adding the average absorbance of the negative control plus 0.042 | - |
| | No | - | Dual-target immuno-fluorescence assay | IgG | NP, SP | - | RBD/N fluorescence value was > 2000 | - |
| Fuereder et al., 2020 | No | - | CLIA | Total antibodies | NP | - | A cut-off index >1 is regarded as positive | Seropositive samples were validated by Abbott CLIA (An index |

| Reference | Paired serums | Days from last possible exposure to sampling (median/range) | Assay methods (screening methods/confirmatory methods) | Antibodies measured | Targeted antigen | Test performance (sensitivity, specificity)* | Reported positive cut-off value | Comments |
|---|---|---|---|---|---|---|---|---|
| | | | | | | | | (S/C) >1.4 is regarded as positive); RT-PCR were also performed for participants. |
| Fusco et al., 2020 | Yes | - | CLIA | IgG, IgM | NP, SP | | - | - |
| Havers et al., 2020 | No | - | ELISA | - | - | Sensitivity: 96.0% (95%CI 89.98 – 98.89%); Specificity: 99.3% (95%CI 98.32 – 99.88%). | A specimen was considered reactive if, on confirmatory testing, at a background corrected optical density (OD) of 0.4 and at a serum dilution of 1:100, it had a signal to threshold ratio of >1. | - |
| Xu et al., 2020 | No | - | ELISA | IgG, IgA | NP, SP | - | - | - |
| Behrens et al., 2020 | No | Mean: 30.4 days | ELISA | IgG, IgA | NP, SP | IgG: Specificity: 99.3% IgA: Specificity: 97.5% | IgG ratio >1.1 were seropositive | - |

| Reference | Paired serums | Days from last possible exposure to sampling (median/range) | Assay methods (screening methods/confirmatory methods) | Antibodies measured | Targeted antigen | Test performance (sensitivity , specificity)* | Reported positive cut-off value | Comments |
|---|---|---|---|---|---|---|---|---|
| | | | Neutralization assays | Neutralizing antibodies | - | - | - | - |
| Loconsole et al., 2020 | No | - | LFIA | IgG, IgM | - | - | - | - |
| Mansour et al., 2020 | No | 2 weeks | ELISA | IgG | SP | - | Antibody titers of⩾320 was considered to be seropositive | - |
| Gallian et al., 2020 | No | - | Neutralization test | Neutralizing antibodies | - | Specificity: 100.0% | - | - |
| Korth et al., 2020 | No | - | ELISA | IgG | SP | - | - | RT-PCR were also performed for participants |
| Bielecki et al., 2020 | No | - | ELISA | IgG, IgM, IgA | - | Sensitivity: 83.0% Specificity: 100.0% | The OD ratio >1.1 was considered positive | RT-PCR were also performed for participants |
| Tsaneva et al., 2020 | Yes | ⩾7 days | LFIA | IgG, IgM | - | - | - | Two of the COVID-19 positive women were tested with a pair of serum samples |

| Reference | Paired serums | Days from last possible exposure to sampling (median/range) | Assay methods (screening methods/confirmatory methods) | Antibodies measured | Targeted antigen | Test performance (sensitivity , specificity)* | Reported positive cut-off value | Comments |
|---|---|---|---|---|---|---|---|---|
| Houlihan et al., 2020 | Yes | - | ELISA | - | SP | - | The cut-off OD values of 0.9 were considered positive | RT-PCR were also performed for participants |
| | | | Flow cytometry | - | SP | - | - | |
| Basteiro et al., 2020 | Yes | ⩾10 days | A serological assay based on the Luminex technique | IgG, IgA, IgM | SP | IgG:<br>Sensitivity: 97.0%<br>Specificity: 98.0%<br>IgA:<br>Sensitivity: 97.0%<br>Specificity: 98.0%<br>IgM:<br>Sensitivity: 75.0%<br>Specificity: 98.0% | Assay cutoff was calculated as 10 to the mean plus 3 standard deviations of log10-transformed median fluorescent intensities (MFIs) of 47 negative controls. | RT-PCR were also performed for participants |
| Isherwood et al., 2020 | - | - | CLIA | IgG | NP | - | - | - |
| Xu et al., 2020 | No | - | CLIA | IgG, IgM | NP, SP | IgG:<br>Sensitivity: 95.0%<br>Specificity: 93.3%<br>IgM: | Antibody levels were expressed as the ratio of the chemiluminescence signal over the cutoff (S/CO) value. An S/CO | RT-PCR were also performed for participants |

| Reference | Paired serums | Days from last possible exposure to sampling (median/range) | Assay methods (screening methods/confirmatory methods) | Antibodies measured | Targeted antigen | Test performance (sensitivity, specificity)* | Reported positive cut-off value | Comments |
|---|---|---|---|---|---|---|---|---|
| | | | | | | Sensitivity: 95%<br>Specificity: 100.0% | value higher than 1.0 for either IgG or IgM was regarded as positive. | |
| Milani et al., 2020 | No | - | ELISA | IgG, IgM, and total antibodies | SP | - | - | RT-PCR were also performed for participants |
| Bryan et al., 2020 | No | - | CLIA | IgG | NP | Sensitivity: 96.9% (89.5-99.5%) at 14 days, and 100% (95.1%-100%) at day 17;<br>Specificity: 99.9% | The index value cutoff of 1.40 was considered positive (according to manufacturer's recommended) | - |
| Hains et al., 2020 | Yes | Within 21 days | ELISA | IgG, IgM | SP | - | A positive ELISA result at 0.14 were considered seropositive. Participants were considered to have seroconverted if positive for IgM or IgG. | RT-PCR were also performed for participants |

| Reference | Paired serums | Days from last possible exposure to sampling (median/range) | Assay methods (screening methods/confirmatory methods) | Antibodies measured | Targeted antigen | Test performance (sensitivity , specificity)* | Reported positive cut-off value | Comments |
|---|---|---|---|---|---|---|---|---|
| Liu et al., 2020 | No | 2 weeks or more | CLIA | IgG, IgM | NP, SP | IgG:<br>Sensitivity: 97.8%<br>Specificity: 97.9%<br>IgM:<br>Sensitivity: 88.2%<br>Specificity: 99.0% | - | RT-PCR were also performed for participants |
| Malickova et al., 2020 | No | - | ELISA | IgG | SP | - | An OD ratio >1.1 was considered positive. | RT-PCR were also performed for participants |
| Lackermair et al., 2020 | No | - | ELISA | IgG | SP | - | - | RT-PCR were also performed for participants |
| Sotgiu et al., 2020 | No | - | Colloidal gold-based assay | IgG, IgM | - | - | Forming a red M line was considered positive. | - |
| Wu et al., 2020 | No | - | Colloidal gold based 2019-nCoV Ab Test | IgG, IgM | - | - | - | RT-PCR were also performed for participants |
| Stubblefield et al., 2020 | No | - | ELISA | IgG, IgM, IgA (pan immunoglobulin) | - | - | - | - |
| Self et al., 2020 | No | - | ELISA | IgG, IgM, IgA | SP | Sensitivity: 96.0%<br>Specificity: 99.0% | A specimen was considered reactive if it | - |

| Reference | Paired serums | Days from last possible exposure to sampling (median/range) | Assay methods (screening methods/confirmatory methods) | Antibodies measured | Targeted antigen | Test performance (sensitivity , specificity)* | Reported positive cut-off value | Comments |
|---|---|---|---|---|---|---|---|---|
| | | | | | | | had a signal to threshold ratio >1.0 at a serum dilution of 1:100, correcting for background. | |
| Patel et al., 2020 | Yes | - | ELISA | Total antibodies | SP | - | - | - |
| Flannery et al., 2020 | No | - | ELISA | IgG, IgM | SP | Sensitivity: 100.0% Specificity: 98.9% | Either IgG or IgM level >0.48 arbitrary units | RT-PCR were also performed for participants |
| Stock et al., 2020 | No | - | ELISA | IgG | NP | - | The cutoff OD value was 1.1 | - |
| Goldberg et al., 2020 | No | - | LFIA | IgG, IgM | - | Sensitivity: 90.0% Specificity: 99.47% (IgM); 99.74% (IgG); | - | RT-PCR were also performed for participants; The assay had validated by EUA |
| Stringhini et al., 2020 | No | - | ELISA | IgG | SP | Sensitivity: 93.0% Specificity: 100.0% | The index value cutoff of 1.10 was considered positive (according to manufacturer's recommended) | - |

| Reference | Paired serums | Days from last possible exposure to sampling (median/range) | Assay methods (screening methods/confirmatory methods) | Antibodies measured | Targeted antigen | Test performance (sensitivity , specificity)* | Reported positive cut-off value | Comments |
|---|---|---|---|---|---|---|---|---|
| Erikstrup et al., 2020 | No | - | LFIA | IgG, IgM | NP, SP | Sensitivity: 82.6% (95%CI 75.7%-88.2%); Specificity: 99.5% (95%CI 98.7%-99.9%) | Samples were concluded as reactive if the IgM, the IgG, or both bands were visible. | - |
| Lahner et al., 2020 | No | - | CLIA | IgG, IgM | - | 14days: Sensitivity: 80.0% 20days: Sensitivity:100.0% | - | RT-PCR were also performed for participants |
| Pallett et al., 2020 | No | More than 14days | LFIA | IgG, IgM | - | IgG: Sensitivity: (95%CI 88.2%-93.4%); Specificity: (95%CI 94.0%-99.0%) IgM: Sensitivity: (95%CI 88.2%-93.4%); | - | - |

| Reference | Paired serums | Days from last possible exposure to sampling (median/range) | Assay methods (screening methods/confirmatory methods) | Antibodies measured | Targeted antigen | Test performance (sensitivity , specificity)* | Reported positive cut-off value | Comments |
|---|---|---|---|---|---|---|---|---|
| Sood et al., 2020 | No | - | LFIA | IgG, IgM | - | Sensitivity: 82.7% (95%CI 76.0%-88.4%); Specificity: 99.5% (95%CI 99.2%-99.7%) | - | The unweighted and weighted proportions of positive tests (either IgM or IgG) in the analysis sample were calculated. |
| Madsen et al., 2020 | No | - | ELISA | IgG | SP | Sensitivity: 95.4% Specificity: 98.3% | - | - |
| Crovetto et al., 2020 | No | - | ELISA | IgG, IgM, IgA | - | - | - | - |
| Gudbjartsson et al., 2020 | - | - | CLIA | Total antibodies | SP, NP | - | - | Positive results for both assays for a test result to be considered positive |
| | | | ELISA | Total antibodies | SP, NP | - | - | |
| Naranbhai et al. | - | | LFIA | IgG, IgM | - | Sensitivity: IgG:85% IgM:80% IgG or IgM:90% Specificity: >99% | - | - |

| Reference | Paired serums | Days from last possible exposure to sampling (median/range) | Assay methods (screening methods/confirmatory methods) | Antibodies measured | Targeted antigen | Test performance (sensitivity , specificity)* | Reported positive cut-off value | Comments |
|---|---|---|---|---|---|---|---|---|
| Martin et al., 2020 | Yes | - | ELISA | IgG | SP | - | - | RT-PCR were also performed for participants |
| Amendola et al., 2020 | No | - | ELISA | IgG | SP | - | - | - |
| Iversen et al., 2020 | No | - | Point-of-care test. | IgG, IgM | | Sensitivity: 82.5% Specificity: 99.5% | - | RT-PCR were also performed for participants |
| Olalla et al., 2020 | No | | LFIA | IgG | SP | - | - | RT-PCR were also performed for participants |
| Cosma et al., 2020 | - | - | LFIA | IgG, IgM | SP, NP | - | The cut-off index (COI) in which a COI > 1.1 indicates a positive result. | RT-PCR were also performed for participants |
| | - | - | CLIA | IgG | SP | - | The antibody concentration is expressed as arbitrary units (AU/mL) and grades the results as | RT-PCR were also performed for participants |

| Reference | Paired serums | Days from last possible exposure to sampling (median/range) | Assay methods (screening methods/confirmatory methods) | Antibodies measured | Targeted antigen | Test performance (sensitivity , specificity)* | Reported positive cut-off value | Comments |
|---|---|---|---|---|---|---|---|---|
| | | | | | | | positive when ≥ 15 AU/mL (CLIA). | |
| Caban-Martinez et al., 2020 | No | | LFIA | IgG, IgM | - | - | - | - |
| Poletti et al., 2020 | No | - | CLIA | IgG | SP | - | A positive result (>15 AU/mL) indicates the presence of IgG antibodies | - |
| Racine-Brzostek et al., 2020 | - | - | CEFA | IgG, IgM | - | - | - | - |
| Rosenberg et al., 2020 | No | - | Microsphere immunoassay | IgG | NP | Sensitivity: 87.9% (95%CI 83.7%-92.1%); Specificity: 99.8% | The mean MFI (median fluorescence intensity) of 90-100 negative DBS was used to set cut-offs | - |
| Daniel et al., 2020 | No | 22 (IQR 15-26) days | ELISA | Total antibodies | SP | - | The cut-off OD values of 1.0 were considered positive. | RT-PCR were also performed for participants |
| | | | MN | Neutralizing antibodies | - | - | Titer > 1:40 were considered to be seropositive. | |

| Reference | Paired serums | Days from last possible exposure to sampling (median/range) | Assay methods (screening methods/confirmatory methods) | Antibodies measured | Targeted antigen | Test performance (sensitivity , specificity)* | Reported positive cut-off value | Comments |
|---|---|---|---|---|---|---|---|---|
| Schmidt et al., 2020 | No | - | ELISA | IgG | SP | - | The cut-off OD values of 1.0 were considered positive | - |
| Moscola et al., 2020 | No | - | ELISA | IgG | SP | Sensitivity:<br>< 10 days after onset of symptoms: 33.3% (1/3)<br>> 10 days after onset of symptoms 80% (4/5); Specificity: 98.5% (197/200) | - | - |
| | No | - | ELISA | IgG | NP, SP | Sensitivity: 95% (19/20);<br>Sensitivity: 98.3% (118/120) | - | - |
| | No | - | CLIA | IgG | NP | Sensitivity:<br>8-13 days: 91.18% (31/34)<br>>14 days: 100% (73/73) ； | - | - |

| Reference | Paired serums | Days from last possible exposure to sampling (median/range) | Assay methods (screening methods/confirmatory methods) | Antibodies measured | Targeted antigen | Test performance (sensitivity , specificity)* | Reported positive cut-off value | Comments |
|---|---|---|---|---|---|---|---|---|
| | | | | | | Specificity: 99.63% (1066/1070) | | |
| | No | | Immunometric | IgG | SP | Sensitivity: 87.5% (42/48) Specificity: 100% (407/407) | - | - |
| | No | | Immunometric | - | SP | Sensitivity: 83.3% (30/36) Specificity: 100% (400/400) | - | - |
| | No | | CLIA | IgG | SP | Sensitivity: 6-14 days: 89.80% (44/49) >15 days: 97.56% (40/41) Specificity: 99.3% (1082/1090) | - | - |
| | No | | CLIA | - | NP | Sensitivity: | - | - |

| Reference | Paired serums | Days from last possible exposure to sampling (median/range) | Assay methods (screening methods/confirmatory methods) | Antibodies measured | Targeted antigen | Test performance (sensitivity , specificity)* | Reported positive cut-off value | Comments |
|---|---|---|---|---|---|---|---|---|
| | | | | | | 7-13 days: 88.10% (52/59)<br>>14 days: 100% (29/29)<br>Specificity: 99.3% (5262/5272) | | |
| Montenegro et al., 2020 | - | - | LFIA | IgG, IgM | - | - | - | - |
| Steensels et al., 2020 | No | - | LFIA | IgG | NP | Sensitivity: 92.2%<br>Specificity: 97.0% | - | - |
| Soriano et al., 2020 | No | - | Immunochromatography | IgG, IgM | SP, NP | - | - | The test has granted EU and FDA approval |
| Eyre et al., 2020 | No | - | CIIA | IgG | NP | - | Abbott Architect (CLIA) with a manufacturer's signal-to-cut-off index of 1.4 | - |
| | | | ELISA | IgG | SP | - | Using net-normalised signal cut-off of 8 | - |

| Reference | Paired serums | Days from last possible exposure to sampling (median/range) | Assay methods (screening methods/confirmatory methods) | Antibodies measured | Targeted antigen | Test performance (sensitivity , specificity)* | Reported positive cut-off value | Comments |
|---|---|---|---|---|---|---|---|---|
| | | | | | | | million as a positive cut-off for ELISA | |
| Shields et al., 2020 | No | - | ELISA | IgG, IgM, IgA | SP | - | - | RT-PCR were also performed for participants |
| Menachemi et al., 2020 | No | - | CIIA | IgG | - | - | - | - |
| Marina et al., 2020 | No | - | LFIA | IgG, IgM | SP | IgG:<br>Sensitivity: 82.1%<br>Specificity: 100.0% | - | - |
| | No | - | CLIA | IgG | NP | Sensitivity: 89.7%<br>Specificity: 100.0% | The amount of IgG antibodies to SARS-CoV-2 in each sample is determined by comparing its chemiluminescent relative light unit (RLU) to the calibrator RLU (index S/C). | - |

| Reference | Paired serums | Days from last possible exposure to sampling (median/range) | Assay methods (screening methods/confirmatory methods) | Antibodies measured | Targeted antigen | Test performance (sensitivity , specificity)* | Reported positive cut-off value | Comments |
|---|---|---|---|---|---|---|---|---|
| Petersen et al., 2020 | No | - | ELISA | IgG, IgM | - | - | - | - |
| Biggs et al., 2020 | No | - | CLIA | Total antibodies | SP | Sensitivity: 93.2%<br>Specificity: 99.0% | - | - |
| Sydney et al., 2020 | No | - | CLIA | IgG | NP | - | - | - |
| Hunter et al., 2020 | No | - | CLIA | IgG | NP | - | - | - |
| Josè et al., 2020 | No | - | CLIA | IgG, IgM | NP, SP | - | IgG: The RLU-ratio of 1.1 positive were considered positive;<br>IgM: The RLU-ratio of 1.0 positive were considered positive; | RT-PCR were also performed for participants |
| Paderno et al., 2020 | No | - | CLIA | IgG | SP | - | - | RT-PCR were also performed for participants;<br>Positive cases were defined as those with positive IgG serology |

| Reference | Paired serums | Days from last possible exposure to sampling (median/range) | Assay methods (screening methods/confirmatory methods) | Antibodies measured | Targeted antigen | Test performance (sensitivity , specificity)* | Reported positive cut-off value | Comments |
|---|---|---|---|---|---|---|---|---|
| | | | | | | | | and/or positive nasal/ pharyngeal swab. |
| Merkely et al., 2020 | No | - | CLIA | IgG | NP | - | - | RT-PCR were also performed for participants; |
| Dioscoridi et al., 2020 | Yes | - | ELISA | IgG, IgM | - | - | - | - |
| Péré et al., 2020 | - | - | CLIA | IgG | NP | - | - | - |
| Torres et al., 2020 | No | - | Colloidal gold immunoassay | IgG, IgM | - | - | - | - |
| Poulikakos et al., 2020 | No | - | CLIA | IgG | NP, SP | - | - | - |
| Veerus et al., 2020 | - | - | CLIA | IgG | NP | - | The IgG antibody level above 1.4 Index (S/C) was defined as a positive result. | - |
| Feehan et al., 2020 | No | - | CLIA | IgG | NP | - | - | RT-PCR were also performed for participants |
| Sutton et al., 2020 | No | - | CLIA | IgG | NP | - | - | - |

| Reference | Paired serums | Days from last possible exposure to sampling (median/range) | Assay methods (screening methods/confirmatory methods) | Antibodies measured | Targeted antigen | Test performance (sensitivity , specificity)* | Reported positive cut-off value | Comments |
|---|---|---|---|---|---|---|---|---|
| Bampoe et al., 2020 | No | - | CLIA | IgG | NP | - | A relative light index > 1.4 was considered to be a positive result | - |
| Tong et al., 2020 | No | - | CLIA | IgG, IgM | NP, SP | - | - | - |
| Mughal et al., 2020 | No | - | LFIA | IgG, IgM | - | - | - | - |
| Zhang et al., 2020 | No | - | ELISA | IgG, IgM, IgA | SP | - | - | RT-PCR were also performed for participants |
| Akinbami et al., 2020 | No | - | CLIA | IgG | SP | - | Signal-to-cutoff ratio >1.0 was considered positive | - |
| Kempen et al., 2020 | No | - | CLIA | IgG | NP | - | - | - |
| Pagani et al., 2020 | - | - | CLIA | IgG | NP | - | - | - |
| Blairon et al., 2020 | No | - | CLIA | IgG | SP | Sensitivity: 100% | A positive result (>15 AU/mL) indicates the presence of IgG antibodies | Confirmed by a semi-quantitative ELISA method |

| Reference | Paired serums | Days from last possible exposure to sampling (median/range) | Assay methods (screening methods/confirmatory methods) | Antibodies measured | Targeted antigen | Test performance (sensitivity , specificity)* | Reported positive cut-off value | Comments |
|---|---|---|---|---|---|---|---|---|
| Noh et al., 2020 | No | - | CLIA | IgG | NP | | A cutoff index (COI, signal sample/cutoff) of ≥ 1.0 was considered positive | |
| | | | Plaque reduction neutralization test | Neutralizing antibodies | - | - | - | - |
| Lidström et al., 2020 | No | - | CLIA | IgG | NP | - | A positive/ negative cut-off of 1.4 S/C was used in line with the manu-facturer's instructions. | - |
| Haizler-Cohen et al., 2020 | - | - | CLIA | IgG | NP | - | - | - |
| Kassem et al., 2020 | No | - | LFIA | IgG, IgM | - | - | - | - |
| Dimcheff et al., 2020 | - | - | CLIA | IgG | NP | | A value greater than or equal to 1.4 RLU is considered a positive antibody response. | - |
| Dodd et al., 2020 | - | - | CLIA | Total antibodies | SP | - | - | - |
| Lundkvist et al., 2020 | No | - | LFIA | IgG | - | Sensitivity: 100.0% Specificity: | - | - |

| Reference | Paired serums | Days from last possible exposure to sampling (median/range) | Assay methods (screening methods/confirmatory methods) | Antibodies measured | Targeted antigen | Test performance (sensitivity, specificity)* | Reported positive cut-off value | Comments |
|---|---|---|---|---|---|---|---|---|
| | | | | | | IgG: 95.0%<br>IgM: 100.0% | | |
| Younas et al., 2020 | - | - | ECLIA | IgG, IgM, IgA | NP | - | Result reported as reactive if cutoff index (COI)>1.0 and non-reactive for COI<1.0 | Seropositive samples were confirmed by ELISA |
| Del Brutto et al., 2020 | No | - | LFIA | IgG, IgM | - | - | - | As IgM and IgG responses in SARS-CoV-2 develop with only a few days of difference, we defined seropositivity as a positive response to any of them. |
| Preprint servers | | | | | | | | |
| Sughayer et al., 2020 | No | - | CLIA | Total antibodies | NP | - | - | - |
| Germain et al., 2020 | - | - | ELISA | Total antibodies | SP | - | Wantai total antibodies positivity threshold ≥ 1.1 | - |

| Reference | Paired serums | Days from last possible exposure to sampling (median/range) | Assay methods (screening methods/confirmatory methods) | Antibodies measured | Targeted antigen | Test performance (sensitivity , specificity)* | Reported positive cut-off value | Comments |
|---|---|---|---|---|---|---|---|---|
| | | | CLIA | IgG | NP | - | Abbott Architect IgG positivity threshold ≥ 1.4 | - |
| Chang et al., 2020 | Yes | - | ELISA | Total antibodies | SP, NP | - | Those IgG or IgM positive samples with the signal to the cutoff ratio (S/CO) ≥10 were further diluted (1:10, 1:40, 1:160..., and 1:40960 by normal saline and tested again. | - |
| | Yes | - | Neutralization test | Neutralizing antibodies | - | - | An ID50 ≥20 was determined as a cutoff value for the presence of neutralizing antibodies. | - |
| Li et al., 2020 | - | - | CLIA | IgG, IgM | SP, NP | - | An S/CO value higher than 1.0 for either IgG or IgM was regarded as positive. | - |

| Reference | Paired serums | Days from last possible exposure to sampling (median/range) | Assay methods (screening methods/confirmatory methods) | Antibodies measured | Targeted antigen | Test performance (sensitivity , specificity)* | Reported positive cut-off value | Comments |
|---|---|---|---|---|---|---|---|---|
| Buss et al., 2020 | - | - | CLIA | IgG | NP | Sensitivity: 77.4%<br>Specificity: 99.9% | 1.4 S/C threshold to define positive result in the main analysis | - |
| Stadlbauer et al., 2020 | Yes | | ELISA | IgG | - | Sensitivity: 95.0%<br>Specificity: 100.0% | - | Confirmatory ELISA was also conducted. |
| Xiong et al., 2020 | Yes | 27-32 days | ELISA | IgG, IgM | - | - | - | RT-PCR were also performed for participants |
| Valenti et al., 2020 | No | - | LFIA | IgG, IgM | NP | IgG:<br>Sensitivity: 100.0%<br>Specificity: 99.2%<br>IgM:<br>Sensitivity: 68.0% | - | - |

| Reference | Paired serums | Days from last possible exposure to sampling (median/range) | Assay methods (screening methods/confirmatory methods) | Antibodies measured | Targeted antigen | Test performance (sensitivity , specificity)* | Reported positive cut-off value | Comments |
|---|---|---|---|---|---|---|---|---|
| | | | | | | Specificity: 99.2% | | |
| Yu et al., 2020 | No | - | CLIA | IgG, IgM | NP, SP | - | - | - |
| Liu et al., 2020 | No | ≥21 days | CLIA | IgG, IgM | NP, SP | - | - | RT-PCR were also performed for participants |
| | | | Colloidal gold test card | IgG, IgM | - | - | - | |
| Tubiana et al., 2020 | | - | ELISA | IgG | NP, SP | - | - | - |
| Skowronski et al., 2020 | No | - | CLIA | Total antibodies | NP, SP | - | Resulted signal to cut-off (S/C) ratios of 1) ≥1.00 considered reactive for Ortho-Clinical Diagnostics; 2) ≥1.40 considered reactive for Abbott Laboratories; 3) ≥1.00 considered reactive for Siemens Healthineers; | All positive specimens were further assessed by gold-standard neutralization assay. |
| | | | MN | Neutralizing antibodies | - | - | | |

| Reference | Paired serums | Days from last possible exposure to sampling (median/range) | Assay methods (screening methods/confirmatory methods) | Antibodies measured | Targeted antigen | Test performance (sensitivity , specificity)* | Reported positive cut-off value | Comments |
|---|---|---|---|---|---|---|---|---|
| Thompson et al., 2020 | No | - | MN | Neutralizing antibodies | - | Sensitivity: 94.11% (95%CI 79.2-100.0%); Specificity: 100.0% (95%CI 98.10-100%) Relative to result from RT-PCR | - | - |
| | | | ELISA | IgG | - | - | - | A second ELISA based assay was used to confirm the analysis |
| Dietrich et al., 2020 | Yes | - | ELISA | IgG | SP | - | Positive reactions were defined as a net OD reading > 0.7 | - |
| Brehm et al., 2020 | Yes | | ELISA | IgG | SP | Specificity: 99.1% | OD ratio >1.5 were seropositive | - |
| Tang et al., 2020 | No | - | Immuno-colloidal gold technology | IgG, IgM | - | - | Both the control line and the test line appear simultaneously | RT-PCR were also performed for participants |

| Reference | Paired serums | Days from last possible exposure to sampling (median/range) | Assay methods (screening methods/confirmatory methods) | Antibodies measured | Targeted antigen | Test performance (sensitivity , specificity)* | Reported positive cut-off value | Comments |
|---|---|---|---|---|---|---|---|---|
| Augusto et al., 2020 | No | - | ELISA | IgG | SP | - | - | RT-PCR were also performed for participants |
| Wang et al., 2020 | No | - | CLIA | IgG, IgA, IgM | NP, SP | | - | RT-PCR were also performed for participants;<br>The detected chemiluminescent signal over background signal was calculated as relative light units (RLU), COI was the ratio of RLU to statistically determined cut-off. |
| Ling et al., 2020 | No | - | Colloidal gold-based immunoassay | IgG, IgM | - | IgG:<br>Sensitivity: 86.7%<br>Specificity: 98.0%<br>IgM:<br>Sensitivity: 76.2%<br>Specificity: 99.0% | - | RT-PCR were also performed for participants |

| Reference | Paired serums | Days from last possible exposure to sampling (median/range) | Assay methods (screening methods/confirmatory methods) | Antibodies measured | Targeted antigen | Test performance (sensitivity , specificity)* | Reported positive cut-off value | Comments |
|---|---|---|---|---|---|---|---|---|
| Paradiso et al., 2020 | Yes | - | Rapid serological test (VivaDiag) | IgG, IgM | - | - | The presence of SARS-CoV-2 IgG and IgM antibodies is indicated by a red/purple line that appears in the specific region | RT-PCR were also performed for participants |
| | | | CLIA. | IgG, IgM | - | - | signal/cutoff (S/C) ratio was 1 | |
| Herzog et al., 2020 | - | - | ELISA | IgG | SP | - | The cut-off OD values of 1.1 and above were considered positive | - |
| Dopico et al., 2020 | No | - | ELISA | IgG | SP | - | - | - |
| Streeck et al., 2020 | - | - | ELISA | IgG, IgA | SP | Specificity: 98.3% | The cut-off OD values of 1.1 and above were considered positive. | RT-PCR were also performed for participants |
| | | | Neutralization assays | Neutralizing antibodies | - | - | The cut-off OD values of 1.1 and above were considered positive. | RT-PCR were also performed for participants |
| Doi et al., 2020 | No | - | Immunochromatographic assay | IgG | - | - | - | - |

| Reference | Paired serums | Days from last possible exposure to sampling (median/range) | Assay methods (screening methods/confirmatory methods) | Antibodies measured | Targeted antigen | Test performance (sensitivity, specificity)* | Reported positive cut-off value | Comments |
|---|---|---|---|---|---|---|---|---|
| Tosato et al., 2020 | No | - | CLIA | IgG, IgM | NP, SP | - | ⩾1.000 kAU/L were considered seropositive (IgM);<br>⩾1.100 kAU/L were considered seropositive (IgG). | RT-PCR were also performed for participants |
| Shakiba et al., 2020 | No | - | Rapid colloidal gold-based immunochromatographic test | IgG, IgM | - | Sensitivity: 63.3%<br>Specificity: 100.0% | - | - |
| Bastiani et al., 2020 | No | - | - | IgG, IgM | - | - | - | - |
| Carozzi et al., 2020 | No | At least 14 days after a diagnostic PCR-positive assay result | LFIA | IgG, IgM | | Sensitivity: 97.0%-99.0%<br>Specificity: 92.0%-95.0% | Presence of the expected control line and of a line at the IgG or IgM position | RT-PCR were also performed for participants |
| Siddiqui et al., 2020 | No | - | CLIA | Total antibodies | NP | - | - | - |
| Kammon et al., 2020 | No | - | LFIA | IgG, IgM | - | - | The presence of only the control line indicates a negative result and valid test; the presence of both the control line and | - |

| Reference | Paired serums | Days from last possible exposure to sampling (median/range) | Assay methods (screening methods/confirmatory methods) | Antibodies measured | Targeted antigen | Test performance (sensitivity , specificity)* | Reported positive cut-off value | Comments |
|---|---|---|---|---|---|---|---|---|
| | | | | | | | the IgM or IgG antibody line indicates a positive result for IgM or IgG antibody, respectively. | |
| Bendavid et al., 2020 | No | - | LFIA | IgG, IgM | SP | Sensitivity: 82.8% (95%CI 76.0%-88.4%) Specificity: 99.5% (95%CI 99.2%-99.7%) | - | - |
| Egerup et al., 2020 | - | - | CLIA | IgG, IgM | NP, SP | | A positive result was defined as values ≥ 8 AU/mL for IgM and ≥ 10 AU/mL for IgG | - |
| Krähling et al., 2020 | No | - | ELISA | IgG | SP | - | The cut-off value was calculated as the average of the OD values plus 4 standard deviations. | RT-PCR were also performed for participants |
| Krähling et al., 2020 | No | - | Neutralization test | Neutralizing antibodies | - | - | - | RT-PCR were also performed for participants |

| Reference | Paired serums | Days from last possible exposure to sampling (median/range) | Assay methods (screening methods/confirmatory methods) | Antibodies measured | Targeted antigen | Test performance (sensitivity , specificity)* | Reported positive cut-off value | Comments |
|---|---|---|---|---|---|---|---|---|
| Nopsopon et al., 2020 | No | - | Locally developed rapid COVID-19 IgG/IgM test kit | IgG, IgM | - | Sensitivity: 94.1% Specificity: 98.0% | - | - |
| Leidner et al., 2020 | No | - | ELISA | IgG | NP | Sensitivity: 80.0% Specificity: 100.0% | - | |
| Fujita et al., 2020 | No | - | ELISA | IgG | NP | - | - | - |
| Psichogiou et al., 2020 | No | - | CLIA | IgG, IgM | SP | - | Samples were concluded as reactive if the IgM or the IgG or both bands were positive. | - |
| Thomas et al., 2020 | No | Days post symptom onset or exposure > 14 | ELISA | Total antibodies | SP | Sensitivity: 100.0% (Days post symptom onset >14) Specificity: 100.0% (Days post symptom onset >14) | The Antibody Index (AI) was calculated by dividing each sample's OD450nm by the serum pooled control mean. Antibody indices were categorized as follows: Negative, ≤ 2.5; Equivocal, 2.51-4.0; Positive > 4.0 | - |
| Cohen et al., 2020 | No | - | CLIA | IgG, IgM | - | - | - | Positive serology was defined as a case |

| Reference | Paired serums | Days from last possible exposure to sampling (median/range) | Assay methods (screening methods/confirmatory methods) | Antibodies measured | Targeted antigen | Test performance (sensitivity , specificity)* | Reported positive cut-off value | Comments |
|---|---|---|---|---|---|---|---|---|
| | | | | | | | | positive for IgM and negative for IgG or positive for IgM and IgG or negative for IgM and positive for IgG. RT-PCR were also performed for participants |
| Rudberg et al., 2020 | No | - | Multi-Antigen IgG Assay | IgG | NP, SP | Sensitivity: 99.4% Specificity: 99.1% | Cutoff was defined as signals above the mean+6 SD | - |
| Sikora et al., 2020 | No | - | Immunochromatographic test | IgG, IgM | - | - | - | - |
| Galán et al., 2020 | No | - | ELISA | IgG | NP, SP | - | The cut-off OD values of 1.1 and above were considered positive | RT-PCR were also performed for participants |
| Frank et al., 2020 | No | - | LFIA | IgG, IgM | - | Sensitivity: 85-100% Specificity: 95%-99% | A test was considered positive for IgM/IgG (IgM/IgG +) if at least IgM or IgG was positive. | RT-PCR were also performed for participants; |

| Reference | Paired serums | Days from last possible exposure to sampling (median/range) | Assay methods (screening methods/confirmatory methods) | Antibodies measured | Targeted antigen | Test performance (sensitivity , specificity)* | Reported positive cut-off value | Comments |
|---|---|---|---|---|---|---|---|---|
| Garralda Fernandez et al., 2020 | No | - | CLIA | IgG, IgM | NP, SP | - | - | RT-PCR were also performed for participants |
| Snoeck et al., 2020 | No | - | ELISA | IgG, IgA | SP | IgG:<br>Sensitivity: 85.7%<br>Specificity: 97.8%<br>IgA:<br>Sensitivity: 92.9%<br>Specificity: 89.2% | The cut-off OD values of 1.1 and above were considered positive | RT-PCR were also performed for participants |
| Comar et al., 2020 | No | - | LFIA | IgG, IgM | | Sensitivity: 95.6%<br>Specificity: 95.2% | - | RT-PCR were also performed for participants |
| Nisar et al., 2020 | No | - | CLIA | IgG, IgM | NP | - | - | - |
| Wang et al., 2020 | - | - | Colloidal gold test | IgG, IgM | | Sensitivity:100.0%<br>Specificity: 100.0% | - | - |
| | | | Neutralization assay | Neutralizing antibodies | | - | A titer of 1:4 or higher indicated seropositivity | - |
| Waterfield et al., 2020 | - | - | CLIA | IgG, Total antibodies | NP, SP | - | Abbott: 1.4 S/C; Roche: 1.0 COI; DiaSorin: 15.0 AU/ml | T |
| Zou et al., 2020 | No | - | ELISA | IgG, IgM | SP | - | - | - |

| Reference | Paired serums | Days from last possible exposure to sampling (median/range) | Assay methods (screening methods/confirmatory methods) | Antibodies measured | Targeted antigen | Test performance (sensitivity , specificity)* | Reported positive cut-off value | Comments |
|---|---|---|---|---|---|---|---|---|
| Nopsopon et al., 2020 | Yes | - | Rapid IgG/IgM test kit | IgG, IgM | - | Sensitivity: 94.1%<br>Specificity: 98.0% | - | - |
| McDade et al., 2020 | Yes | - | ELISA | IgG | SP | - | - | RT-PCR were also performed for participants |
| Appa et al., 2020 | No | - | CLIA | IgG | NP | - | - | - |
| | | | ELISA | IgG | SP | - | - | - |
| | | | ELISA | IgG, IgA | SP | - | | |
| | | | MN | Neutralizing antibodies | - | - | Titer >1:4 were considered positive | |
| Tönshoff et al., 2020 | - | - | ELISA | IgG | - | - | | Unclear or discordant results were further assessed by CLIA or a second ELISA |
| | | | Immunofluorescence | IgG | - | - | | |
| | | | Neutralization assay | Neutralizing antibodies | | - | | |
| Jerković et al., 2020 | No | - | Rapid immunochromatographic assay | IgG, IgM | - | - | - | - |
| Alessandro et al., 2020 | No | - | CLIA | IgG | SP | - | Positive or negative results were established by the following cuts off: | - |

| Reference | Paired serums | Days from last possible exposure to sampling (median/range) | Assay methods (screening methods/confirmatory methods) | Antibodies measured | Targeted antigen | Test performance (sensitivity , specificity)* | Reported positive cut-off value | Comments |
|---|---|---|---|---|---|---|---|---|
| | | | | | | | <12: Negative; ≥15: positive. | |
| Halatoko et al., 2020 | No | - | LFIA | IgG, IgM | NP, SP | Sensitivity: 77.1%<br>Specificity: 95.4% | - | RT-PCR were also performed for participants;<br>This test was validated by Laboratory Department of the ministry of health in Togo |
| Dillner et al., 2020 | - | - | Multiplex, microsphere-based assay | IgG | NP, SP | Sensitivity: 99.2%<br>Specificity: 99.8% | - | - |
| Aziz et al., 2020 | - | - | ELISA | IgG | SP | - | cut-off of >1.1 | Two additional confirmatory tests in all those individuals whose ELISA assay results were either positive (i.e. >1·1) or borderline (i.e. between 0·8 and 1·1) were performed |

| Reference | Paired serums | Days from last possible exposure to sampling (median/range) | Assay methods (screening methods/confirmatory methods) | Antibodies measured | Targeted antigen | Test performance (sensitivity , specificity)* | Reported positive cut-off value | Comments |
|---|---|---|---|---|---|---|---|---|
| | | | In-house immunofluorescent test | - | - | - | - | - |
| | | | Plaque reduction neutralization test | Neutralizing antibodies | - | - | - | - |
| Chamie et al., 2020 | No | - | CLIA | IgG | NP | - | - | RT-PCR were also performed for participants |
| Nesbitt et al., 2020 | No | - | LFIA | IgG, IgM | - | - | - | - |
| | | | CLIA | Total antibodies | SP | - | - | - |
| Wells et al., 2020 | No | - | ELISA | IgG | SP, NP | Sensitivity: 90.0% (95%CI 60.0%-99.0%) Specificity: 100.0% (95%CI 93.0%-100.0%) | A participant was considered seropositive if an IgG response (OD value) to both N and S was detected that was 4-fold above the background of the assay. This cut-off is based on the analysis of 300+ pre-COVID-19 serum samples | RT-PCR were also performed for participants |

| Reference | Paired serums | Days from last possible exposure to sampling (median/range) | Assay methods (screening methods/confirmatory methods) | Antibodies measured | Targeted antigen | Test performance (sensitivity , specificity)* | Reported positive cut-off value | Comments |
|---|---|---|---|---|---|---|---|---|
| Fontanet et al., 2020 | No | - | Flowcytometry-based serological test | - | SP | - | - | - |
| Sandri et al., 2020 | No | - | CLIA | IgG | SP | - | > 15.0 AU/mL was considered positive. | RT-PCR were also performed for participants |
| Uyoga et al., 2020 | No | - | ELISA | IgG | SP | Sensitivity: 83.0% (95%CI 59.0-96.0%) Specificity: 99.0% (95%CI 98.1-99.5%) | we defined anti- SARS-CoV-2 IgG seropositivity as an OD ratio >2 | - |
| Brant-Zawadzki et al., 2020 | No | - | CLIA | IgG | SP | - | - | - |
| Addetia et al., 2020 | No | - | CLIA | IgG | NP | - | - | The presence of anti-Spike and neutralizing antibodies was analyzed in pre-departure sera samples from individuals that were positive in the Abbott assay screening through four different methods |

| Reference | Paired serums | Days from last possible exposure to sampling (median/range) | Assay methods (screening methods/confirmatory methods) | Antibodies measured | Targeted antigen | Test performance (sensitivity , specificity)* | Reported positive cut-off value | Comments |
|---|---|---|---|---|---|---|---|---|
| | | - | Neutralization assay | Neutralizing antibodies | - | - | - | - |
| Barallat et al., 2020 | No | - | CLIA | IgG (spike);<br>IgG (nucleocapsid) | NP, SP | - | IgG (spike):> 15.0 AU/mL were considered positive;<br>IgG (nucleocapsid): antibody levels were expressed as the ratio of the chemiluminescence signal over the cutoff (S/CO) value. An S/CO value higher than 1.4 for either IgG positive. | - |
| Tess et al., 2020 | No | - | CLIA | IgG, IgM | NP, SP | - | IgG: reagent >1.1 UA/mL,<br>IgM:<br>reagent >1.0UA/mL | Individuals who were reactive to either IgM or IgG were considered positive |
| Takita et al., 2020 | No | - | Point-of-care immunodiagnostic test | IgG | - | Sensitivity: 76.4%<br>Specificity: 100.0% | - | - |

| Reference | Paired serums | Days from last possible exposure to sampling (median/range) | Assay methods (screening methods/confirmatory methods) | Antibodies measured | Targeted antigen | Test performance (sensitivity , specificity)* | Reported positive cut-off value | Comments |
|---|---|---|---|---|---|---|---|---|
| Mattern et al., 2020 | No | - | CLIA | IgG | NP | - | Positive if the IgG index value was 1.40 | RT-PCR were also performed for participants |
| Carrat et al., 2020 | - | - | ELISA | IgG | NP, SP | - | ELISA-positive with an optical density ratio ≥ 1.1 | - |
| | | | Neutralization assay | Neutralizing antibodies | - | - | Positive SN defined as a titer ≥40 | - |
| McBride et al., 2020 | No | - | CLIA | IgG | NP | - | - | - |
| Ebinger et al., 2020 | No | - | CLIA | IgG | NP | - | Manufacturer's signal-to-cut-off index of 1.4 | - |
| Hurk et al., 2020 | No | - | ELISA | Total antibodies | SP | Sensitivity: 98.7% Specificity: 99.6% | OD/CO ratio >1.0 were considered positive | - |
| Weis et al., 2020 | No | - | ELISA | IgG | - | - | - | - |
| | | | CLIA | IgG | - | - | - | - |
| Rigatti et al., 2020 | | | CLIA | Total antibodies | NP | - | - | - |
| Gomes et al., 2020 | No | - | Immunochromatographic test | IgG, IgM | - | - | - | - |
| Hallal et al., 2020 | No | - | LFIA | IgG, IgM | - | Sensitivity: 77.1% Specificity: 98.0% | - | - |
| | No | - | CLIA (Abbott) | IgG | NP | - | - | |

| Reference | Paired serums | Days from last possible exposure to sampling (median/range) | Assay methods (screening methods/confirmatory methods) | Antibodies measured | Targeted antigen | Test performance (sensitivity , specificity)* | Reported positive cut-off value | Comments |
|---|---|---|---|---|---|---|---|---|
| Nakamura et al., 2020 | | | CLIA (Roche) | IgG, IgM | NP | - | - | Weak signals for IgM and IgG, together or separate, were considered positive. |
| | | | POC qualitative test | IgG, IgM | NP | - | - | |
| Jespersen et al., 2020 | No | - | ELISA | Total antibodies | SP | - | The sample absorbance (A) value was divided by a cut- off (CO) value for the ELISA plate based on an average absorbance value for 3 negative kit controls. A/CO values ≥ 1.1 were considered positive. | - |
| Tsertsvadze et al., 2020 | - | - | LFIA | IgG | - | - | - | - |
| Chibwana et al., 2020 | No | - | ELISA | IgG | SP, NP | | The assay interpretation was as follows; positive result (OD 0.6), indeterminate result (OD 0.55 to < 0.6) and negative (OD < 0.55) | - |

| Reference | Paired serums | Days from last possible exposure to sampling (median/range) | Assay methods (screening methods/confirmatory methods) | Antibodies measured | Targeted antigen | Test performance (sensitivity , specificity)* | Reported positive cut-off value | Comments |
|---|---|---|---|---|---|---|---|---|
| Armann et al., 2020 | No | - | CLIA | IgG | SP | - | Antibody levels > 15.0 AU/ml were considered positive | - |
| | | | ELISA | IgG | SP | - | A ratio > 1.1 was considered positive | - |
| | | | CLIA | IgG | NP | - | An index (S/C) of >/= 1.4 was considered positive | - |
| Hibino et al., 2020 | Yes | - | LFIA | IgG, IgM | - | - | - | - |
| Wilkins et al., 2020 | - | - | CLIA | IgG | NP | - | - | - |
| Alkurt et al., 2020 | No | - | CLIA | IgG | NP | - | Cut-off value of 1.40 S/C was considered positive | - |
| Vassallo et al., 2020 | - | - | CLIA | IgG, IgM, IgA | SP | - | Results with signal-to-cutoff (S/C) ratios ≥1 are reported as positive | - |
| Melo et al., 2020 | - | - | Immunofluorescence assays | IgG, IgM | - | - | - | - |
| Favara et al., 2020 | No | - | Rapid point-of-care assay | IgG, IgM | NP, SP | - | - | RT-PCR were also performed for participants |
| | | | Microsphere-based assay | IgG | NP, SP | - | - | |

| Reference | Paired serums | Days from last possible exposure to sampling (median/range) | Assay methods (screening methods/confirmatory methods) | Antibodies measured | Targeted antigen | Test performance (sensitivity , specificity)* | Reported positive cut-off value | Comments |
|---|---|---|---|---|---|---|---|---|
| Silva et al., 2020 | - | - | ELISA | IgG | SP | Sensitivity: 74.0%<br>Specificity: 100.0% | | |
| Ray et al., 2020 | No | - | ELISA | IgG | SP | Sensitivity: 88.2%<br>Specificity: 99.8% | - | - |
| Bardai et al., 2020 | - | - | ELISA | IgG | NP, SP | - | - | - |
| Mahajan et al., 2020 | No | - | CLIA | IgG | SP | - | Antibody levels were expressed as the ratio of the chemiluminescence signal over the cutoff (S/CO) value. An S/CO value≥1.00 was reported as positive | - |
| Nishida et al., 2020 | - | - | CLIA | IgG | NP | - | - | - |
| Nawa et al., 2020 | No | - | CLIA | IgG | NP, SP | - | A cut-off value of 10 AU/ml was considered positive | - |
| Qutob et al., 2020 | - | - | CLIA | Total antibodies | NP | - | - | - |
| Ulyte et al., 2020 | - | - | Microsphere-based assay | IgG, IgM, IgA | NP, SP | Sensitivity: 93.3%<br>Specificity: 99.6% | - | - |

| Reference | Paired serums | Days from last possible exposure to sampling (median/range) | Assay methods (screening methods/confirmatory methods) | Antibodies measured | Targeted antigen | Test performance (sensitivity , specificity)* | Reported positive cut-off value | Comments |
|---|---|---|---|---|---|---|---|---|
| Asuquo et al., 2020 | No | - | LFIA | IgG, IgM | SP | - | Both the quality control line (C) and the detection line M/G appeared (red) | - |
| Ward et al., 2020 | No | - | LFIA | IgG | - | - | - | - |
| Menezes et al., 2020 | No | - | LFIA | IgG, IgM | - | Sensitivity: 77.1% Specificity: 98.0% | - | - |
| Ariza et al., 2020 | Yes | - | CLIA | IgG | NP | - | Samples with a signal-to-cutoff (S/CO) ratio greater than or equal to 1.4 were considered positive | - |
| Majiya et al., 2020 | No | - | LFIA | IgG, IgM | - | Sensitivity: 100.0% Specificity: 100.0% | - | - |
| Javed et al., 2020 | No | - | Colloidal gold assay | IgG, IgM | - | - | - | - |
| Buonsenso et al., 2020 | No | - | ELISA | IgG | NP, SP | - | - | - |
| Khan et al., 2020 | - | - | CLIA | IgG | NP | - | - | - |
| Satpati et al., 2020 | - | - | ELISA | IgG | - | - | - | - |

| Reference | Paired serums | Days from last possible exposure to sampling (median/range) | Assay methods (screening methods/confirmatory methods) | Antibodies measured | Targeted antigen | Test performance (sensitivity , specificity)* | Reported positive cut-off value | Comments |
|---|---|---|---|---|---|---|---|---|
| Silva et al., 2020 | - | - | CLIA | IgG, IgM | NP | - | - | - |
| Calife et al., 2020 | - | - | CLIA | IgG, IgM | NP | - | - | - |
| Kumar et al., 2020 | No | - | CLIA | IgG | NP | | An index measurement ≥1.4 was considered positive | - |
| | | | ELISA | IgG | SP | | Results are evaluated semi-quantitatively by calculation of a ratio of the extinction of the control or patient sample over the extinction of the calibrator, and ratio of < 1.1 was positive | - |
| Official reports | | | | | | | | |
| Public Health Ontario, Canada, 2020 | - | - | CLIA | IgG | NP, SP | Sensitivity: 90.4% Specificity: 100.0% | - | - |

| Reference | Paired serums | Days from last possible exposure to sampling (median/range) | Assay methods (screening methods/confirmatory methods) | Antibodies measured | Targeted antigen | Test performance (sensitivity , specificity)* | Reported positive cut-off value | Comments |
|---|---|---|---|---|---|---|---|---|
| Office of National Statistics, UK, 2020 | No | - | CLIA | IgG | | - | - | - |
| the Government of Jersey, UK, 2020 | Yes | - | LFIA | IgG, IgM | - | - | - | - |
| Canadian Blood Services, 2020 | - | - | CLIA | IgG | NP | - | - | - |
| Ministry of Health, Labour and Welfare, Japan, 2020 | - | - | CLIA | IgG, IgM | NP | - | - | - |
| Islamic Republic of Afghanistan Ministry of Public Health, Afghanistan, 2020 | - | - | LFIA | IgG, IgM | - | - | - | - |
| Public Health England, 2020 | - | - | ELISA | - | SP | - | - | - |
| MedLife, Romania, 2020 | No | - | CLIA | - | | - | - | - |

| Reference | Paired serums | Days from last possible exposure to sampling (median/range) | Assay methods (screening methods/confirmatory methods) | Antibodies measured | Targeted antigen | Test performance (sensitivity , specificity)* | Reported positive cut-off value | Comments |
|---|---|---|---|---|---|---|---|---|
| | No | - | ELISA | - | | - | - | - |

Abbreviations: ELISA, Enzyme-linked immunosorbent assay; CLIA: Chemiluminescent immunoassay; LFIA: lateral flow immunoassays; MIA: Microsphere immunoassay; MN, Microneutralisation assay; OD value: Optical density value; POC: point of care; RLU: relative light unit

Notes: Chemiluminescence microparticle immunoassay (CMIA), a kind of CLIA assay, also named as "CLIA".

#### Appendix Table 4. Summary of studies reporting seroprevalence, seroconversion and seroincidence of human infections with SARS-CoV-2 included in systematic review

| Reference | Starting month | Study population | No. of positive/total no. of participants provided sera (seroprevalence rate, %) | Risk factors for SARS-CoV-2 infections (OR, 95%CI) |
|---|---|---|---|---|
| Peer-reviewed databases | | | | |
| Victoria et al., 2020 | Jan 2020 | Office co-workers<br>waiting room contacts<br>healthcare contacts | 0/8 (0)<br>0/14 (0)<br>0/6 (0) | - |
| To et al., 2020 | Jan 2020 | General population<br>Hong Kong resident evacuated from Hubei | IgG (either anti-NP or anti-RBD): 53/1938 (2.7);<br>Neutralizing antibodies: 0/1938 (0)<br>IgG (either anti-NP or anti-RBD): 13/452 (2.9);<br>Neutralizing antibodies: 15/452 (3.3) | - |
| Liang et al., 2020 | Jan 2020 | Inpatients and their healthy companions | Guangzhou: IgG or IgM: 52/8782 (0.6); IgG: 14/8782 (0.2);<br>IgM: 39/8782 (0.4)<br>Wuhan: IgG or IgM: 177/8272 (2.1); IgG: 123/8272 (1.5);<br>IgM: 71/8272 (0.9) | - |
| Fischer et al., 2020 | Jan 2020 | Blood donors | 29/3186 (0.9) | - |
| Hallowell et al., 2020 | Jan 2020 | Evacuees from Wuhan in a repatriation | 1/186 (0.5) | - |
| Sam et al., 2020 | Jan 2020 | Residual serum samples collected at a teaching hospital | IgG: 46/588 (7.8)<br>neutralizing antibodies: 3/588 (0.5) | - |

| Reference | Starting month | Study population | No. of positive/total no. of participants provided sera (seroprevalence rate, %) | Risk factors for SARS-CoV-2 infections (OR, 95%CI) |
|---|---|---|---|---|
| Chen et al., 2020 | Feb 2020 | Healthcare workers | IgG or IgM: 19/105 (18.1)<br>neutralizing antibodies: 18/105 (17.1) | Univariate analysis：<br>Exposure for more than 30 minutes at less than 1 meter: 3.478 (1.224-9.887), ref: no exposure;<br>Close contact with patient 2: 7.125(1.627-31.210), ref: close contact with patient 1;<br>Doctors: 3.850 (1.131- 13.105), ref: colleague;<br>Multivariate analysis:<br>Close contacts with patient 2: 6.605(1.123-38.830), ref: close contact with patient 1;<br>Doctor: 346.837 (8.924-13479.434), ref: colleague; Wearing disposable non-surgical face mask: 0.127 (0.017-0.968), ref: without wearing disposable non-surgical face mask |
| Cavicchiolo et al., 2020 | Feb 2020 | Neonates | 0/75 (0.0) | - |
| Plebani et al., 2020 | Feb 2020 | Healthcare worker | IgG: 343/8285 (4.1)<br>IgM: 82/8285 (1.0)<br>IgG or IgM: 378/8285 (4.6) | A significant higher seroprevalence could be observed in health care assistants compared to other groups ($\chi 2=5.34$, $p=0.021$) |
| Cox et al., 2020 | Feb 2020 | Household members of confirmed COVID-19 cases | 24/77 (31.2) | - |

| Reference | Starting month | Study population | No. of positive/total no. of participants provided sera (seroprevalence rate, %) | Risk factors for SARS-CoV-2 infections (OR, 95%CI) |
|---|---|---|---|---|
| Brandstetter et al., 2020 | Mar 2020 | Hospital staff with close contact<br>Hospital staff with moderate contact<br>Hospital staff with no contact | IgG: 1/50 (2.0); IgA: 3/50 (6.0); IgG or IgA: 4/50 (8.0)<br>IgG: 0/63 (0); IgA: 1/63 (1.6); IgG or IgA: 1/63 (1.6)<br>IgG: 0/50 (0); IgA: 6/50 (12.0) ; IgG or IgA: 6/50 (12.0) | - |
| Solodky et al., 2020 | Mar 2020 | Healthcare worker,<br>cancer patients | 13/244 (5.3)<br>5/85 (5.9) | - |
| Zhang et al., 2020 | Mar 2020 | Healthy individuals returning to Shenzhen | IgG: 6/1589 (0.4); IgA: 0/1589 (0); IgM: 0/1589 (0) | - |
| Suda et al., 2020 | Mar 2020 | Outpatients with liver disease | IgG (Immunochromatographic test): 2/300 (0.67)<br>IgG (CLIA) :1/600 (0.17) | - |
| Bogogiannidou et al., 2020 | Mar 2020 | Leftover blood samples from nationwide labs | 24/6586 (0.36) | - |
| Xu et al., 2020 | Mar 2020 | Hemodialysis Patients;<br>healthcare worker; | 51/1542 (3.3)<br>39/3205 (1.2) | Independent risk factors for SARS-CoV-2 infection were being older than 65 years, having manifestation of lung infection in imaging examinations, and having a lower level of serum albumin. |

| Reference | Starting month | Study population | No. of positive/total no. of participants provided sera (seroprevalence rate, %) | Risk factors for SARS-CoV-2 infections (OR, 95%CI) |
|---|---|---|---|---|
| Vena et al., 2020 | Mar 2020 | non-hospitalized participants in an outpatient setting | 398/3609 (11.0) | Factors Associated with Anti-Sars-CoV-2 Antibodies Positivity:<br>occupational exposure to the virus: 2.36 (1.59–3.50);<br>living in a long-term care facility: 4.53 (3.19–6.45);<br>reporting previous symptoms of influenza-like illness: 4.86 (3.75–6.30);<br>loss of sense of smell or taste: 41.00 (18.94–88.71) |
| Ng et al., 2020 | Mar 2020 | Blood donors;<br>Hospitalized patients admitted for non-respiratory indications | 1/1000 (0.1)<br>1/387 (0.3) | - |
| Dingens et al., 2020 | Mar 2020 | Residual serum samples from Seattle Children's Hospital | 8/1076 (0.7) | - |
| Brown et al., 2020 | Mar 2020 | Student who contacted with infected teacher | 1/21 (4.8) | - |
| Han et al., 2020 | Mar 2020 | Persons during work resumption screening | IgG: 813/22633 (3.6)<br>IgM: 236/22633 (2.0)<br>IgG and IgM: 196/22633 (0.9) | - |
| De et al., 2020 | Mar 2020 | Hemodialysis patients | 7/282 (2.5) | - |

| Reference | Starting month | Study population | No. of positive/total no. of participants provided sera (seroprevalence rate, %) | Risk factors for SARS-CoV-2 infections (OR, 95%CI) |
|---|---|---|---|---|
| Zhou et al., 2020 | Mar 2020 | Hospital staff | IgG or IgM: 89/3674 (2.4)<br>IgG: 73/3674 (2.0)<br>IgM: 26/3674 (0.7) | - |
| Tu et al., 2020 | Mar 2020 | Pediatric medical workers (close contact group) | ELISA:<br>IgG: 66/191 (34.6)<br>IgM: 16/191 (8.4)<br>dual-target immuno-fluorescence assay:<br>IgG: 79/191 (41.4) | - |
| | | Pediatric medical workers (Non-close contact group) | ELISA:<br>IgG: 12/110 (10.9)<br>IgM: 1/110 (0.9)<br>dual-target immuno-fluorescence assay:<br>IgG: 16/109 (14.7) | |
| | | Pediatric medical workers (Non-contact group) | ELISA:<br>IgG: 1/24 (4.2)<br>IgM: 0/24 (0)<br>dual-target immuno-fluorescence assay:<br>IgG: 3/24 (12.5) | |
| Fuereder et al., 2020 | Mar 2020 | Healthcare professionals<br>Cancer patients | 2/62 (3.2)<br>2/84 (2.3) | - |

| Reference | Starting month | Study population | No. of positive/total no. of participants provided sera (seroprevalence rate, %) | Risk factors for SARS-CoV-2 infections (OR, 95%CI) |
|---|---|---|---|---|
| Fusco et al., 2020 | Mar 2020 | Healthcare worker | IgG: 2/115 (1.7)<br>IgM: 0/115 (0.0) | - |
| Havers et al., 2020 | Mar 2020 | Residual patient sera collected for routine screening | Washington: 43/3264 (1.3)<br>New York City: 144/2482 (5.8)<br>Louisiana: 81/1184 (6.8)<br>South Florida: 38/1742 (2.2)<br>Pennsylvania: 20/824 (2.4)<br>Missouri: 54/1882 (2.9)<br>Utah: 26/1132 (2.3)<br>California: 12/1224 (1.0)<br>Connecticut: 70/1431 (4.9)<br>Minnesota: 14/860(1.6) | - |
| Xu et al., 2020 | Mar 2020 | Blood donors | IgG: 2/2199 (0.1)<br>IgA: 2/2199 (0.1)<br>Total antibodies: 7/2199 (0.3) | - |
| Behrens et al., 2020 | Mar 2020 | Firstline health care professional | IgG: 2/217 (0.9)<br>IgA: 9/217 (4.1)<br>Neutralizing antibodies: 1/217 (0.5) | - |
| Loconsole et al., 2020 | Mar 2020 | Patients admitted to Emergency Department | 70/819 (8.5) | - |
| Mansour et al., 2020 | Mar 2020 | Healthcare worker | 93/285 (32.6) | - |
| Gallian et al., 2020 | Mar 2020 | Blood donors | 27/998 (2.7) | - |

| Reference | Starting month | Study population | No. of positive/total no. of participants provided sera (seroprevalence rate, %) | Risk factors for SARS-CoV-2 infections (OR, 95%CI) |
|---|---|---|---|---|
| Korth et al., 2020 | Mar 2020 | High-risk Healthcare worker,<br>Intermediated-risk healthcare worker,<br>Low-risk healthcare worker, | 3/244 (1.2)<br>2/37 (5.4)<br>0/35 (0.0) | - |
| Bielecki et al., 2020 | Mar 2020 | Soldiers stationed at a Swiss Army Base<br>Company 1<br>Company 2 | <br>7/88 (8.0)<br>111/181 (61.3) | - |
| Tsaneva et al., 2020 | Mar 2020 | Outpatients | IgG: 22/586 (3.8); IgM: 13/586 (2.2); IgG or IgM: 28/586 (4.8) | |
| Houlihan et al., 2020 | Mar 2020 | First-line healthcare worker | 46/181 (25.4) | - |
| Basteiro et al., 2020 | Mar 2020 | Health care workers | IgG or IgM or IgA: 54/578 (9.3); IgG: 44/578 (7.6); IgM: 36/578 (6.2); IgA: 47/578 (8.1) | - |
| Isherwood et al., 2020 | Mar 2020 | Patients in a tertiary acute general surgical unit<br>Healthcare staff in the same healthcare setting | 71/1964 (3.6)<br>15/215 (7.0) | - |

| Reference | Starting month | Study population | No. of positive/total no. of participants provided sera (seroprevalence rate, %) | Risk factors for SARS-CoV-2 infections (OR, 95%CI) |
|---|---|---|---|---|
| Xu et al., 2020 | Mar 2020 | Healthcare worker in Wuhan<br>Healthcare worker in Hubei<br>Healthcare worker in Chongqing<br>Healthcare worker in Guangdong<br>Healthcare worker relative in Wuhan<br>Hemodialysis patient in Hubei<br>Hemodialysis patient in Guangdong<br>Outpatient in Chongqing<br>Hotel staff member in Wuhan<br>Community resident in Sichuan<br>Factory workers in Guangdong | IgG:27/714 (3.8); IgM:6/714 (0.8); IgG or IgM: 27/714 (3.8)<br>IgG:37/3091 (1.2); IgM:4/3091 (0.1); IgG or IgM: 41/3091 (1.3)<br>IgG:8/319 (2.5); IgM:2/319 (0.6); IgG or IgM: 10/319 (3.1)<br>IgG:1/260 (0.4); IgM:2/260 (0.8); IgG or IgM: 3/260 (1.2)<br>IgG:7/219 (3.2); IgM:3/219 (1.4); IgG or IgM: 7/219 (3.2)<br>IgG:19/979 (1.9); IgM:19/979 (1.9); IgG or IgM: 35/979 (3.6)<br>IgG:12/563 (2.1); IgM:7/563 (1.2); IgG or IgM: 16/563 (2.8)<br>IgG:37/993 (3.7); IgM:1/993 (0.1); IgG or IgM: 38/993 (3.8)<br>IgG:11/346 (3.2); IgM:8/346 (2.3); IgG or IgM: 13/346 (3.8)<br>IgG:26/9442 (0.3); IgM:29/9442 (0.3); IgG or IgM: 55/9442 (0.6)<br>IgG:4/442 (0.9); IgM: 4/442 (0.9); IgG or IgM:6 /442 (1.4) | - |
| Milani et al., 2020 | Mar 2020 | Personnel of the University of Milan | Total antibodies:5/197 (2.5)<br>IgM:5/197 (2.5)<br>IgG:11/197 (5.6) | - |
| Bryan et al., 2020 | Apr 2020 | Community resident | 87/4856 (1.8) | - |

| Reference | Starting month | Study population | No. of positive/total no. of participants provided sera (seroprevalence rate, %) | Risk factors for SARS-CoV-2 infections (OR, 95%CI) |
|---|---|---|---|---|
| Hains et al., 2020 | Apr 2020 | Hemodialysis patients<br>Healthcare worker | IgG:3/13 (23.1); IgM:2/13 (15.4)<br>IgG:7/25 (28.0); IgM:4/25 (16.0) | - |
| Liu et al., 2020 | Apr 2020 | Healthcare worker deployed to Wuhan<br>Healthcare professionals at home hospital | IgG:0/420 (0.0); IgM: 0/420 (0.0)<br>IgG:0/77 (0.0); IgM: 0/77 (0.0) | - |
| Malickova et al., 2020 | Apr 2020 | Inflammatory bowel disease healthcare professionals | 2/92 (2.2) | - |
| Lackermair et al., 2020 | Mar 2020 | Healthcare worker | 4/151 (2.6) | - |
| Sotgiu et al., 2020 | Mar 2020 | Healthcare worker | IgM: 29/202 (14.4)<br>IgG: 29/202 (7.4) | - |
| Wu et al., 2020 | Apr 2020 | People applying for a permission of resume<br>Hospitalized patients | IgG:98/1021 (9.6); IgM: 0/1021 (0.0)<br>IgG:40/381(10.5); IgM: 1/381 (0.0) | - |
| Stubblefield et al., 2020 | Apr 2020 | Healthcare worker worked in COVID-19 units | 19/249 (7.6) | - |
| Self et al., 2020 | Apr 2020 | frontline Health care personnel | 194/3248 (6.0) | Detection of SARS-CoV-2 antibodies was less common among participants who reported using a face covering for all clinical encounters (6%) than among those who did not (9%) (p = 0.012). |
| Patel et al., 2020 | Apr 2020 | health care personnel | 19/249 (7.6) | - |

| Reference | Starting month | Study population | No. of positive/total no. of participants provided sera (seroprevalence rate, %) | Risk factors for SARS-CoV-2 infections (OR, 95%CI) |
|---|---|---|---|---|
| Flannery et al., 2020 | Apr 2020 | Pregnant women presenting for delivery | IgG or IgM: 80/1293 (6.2)<br>IgG: 76/1293 (5.9)<br>IgM: 59/1293 (4.6) | Black/non-Hispanic and Hispanic/Latino women have higher SARS-CoV-2 seroprevalence rates relative to women of other races |
| Stock et al., 2020 | Apr 2020 | Adult clinicians | 15/98 (15) | - |
| Goldberg et al., 2020 | Apr 2020 | Staff members at a Skilled Nursing Facility;<br>residents at a Skilled Nursing Facility | 4/84 (4.8)<br>11/56 (19.6) | - |
| Stringhini et al., 2020 | Apr 2020 | General population | Week1: 12/341 (3.5)<br>Week2: 28/469 (6.0)<br>Week3: 61/577 (10.6)<br>Week4: 36/604 (6.0)<br>Week5: 82/775 (10.6)<br>Overall: 219/2766 (7.9) | Univariate analysis：<br>Aged 5–9 years:0.32 (0.11-0.63);<br>65 years and older:0.50 (0.28-0.78);<br>ref: aged 20–49 years |
| Erikstrup et al., 2020 | Apr 2020 | Blood donors | 412/20640 (2.0) | - |
| Lahner et al., 2020 | Apr 2020 | Healthcare worker | IgG: 8/1084 (0.7)<br>IgM: 0/1084 (0.0) | - |
| Pallett et al., 2020 | Apr 2020 | Health-care workers | IgG: 624/1704 (36.6)<br>IgM: 45/1704 (2.6) | - |
| Sood et al., 2020 | Apr 2020 | General population | 35/863 (4.1) | - |
| Madsen et al., 2020 | Apr 2020 | ED employees | 16/270 (6.0) | - |

| Reference | Starting month | Study population | No. of positive/total no. of participants provided sera (seroprevalence rate, %) | Risk factors for SARS-CoV-2 infections (OR, 95%CI) |
|---|---|---|---|---|
| Crovetto et al., 2020 | Apr 2020 | Pregnant women attending first trimester screening | 125/874 (14.3) | - |
| Gudbjartsson et al., 2020 | Apr 2020 | persons contact with the Icelandic health care system for reasons other than Covid-19<br>Icelandersi n the greater Reykjavik area<br>Residents of Vestmannaeyjar<br>Icelanders had been quarantined | 39/18609 (0.2)<br>21/4843 (0.4)<br>3/663 (0.5)<br>97/4222 (2.3) | - |
| Naranbhai et al. | Apr 2020 | Asymptomatic residents | IgG or IgM: 63/200 (31.5)<br>IgG: 45/200 (22.5)<br>IgM: 53/200 (26.5) | The number of cohabiting children: 1.057 (1.001-1.117);reduced sense of smell or taste: 1.519 (1.208-1.910) |
| Martin et al., 2020 | Apr 2020 | General population | 36/326 (11.0) | Presence of at least one comorbidity and symptoms at the time of collection increased the risk of a positive PCR and/or serology test (p<0.05) |
| Amendola et al., 2020 | Apr 2020 | Healthcare worker | 34/663(5.1) | - |

| Reference | Starting month | Study population | No. of positive/total no. of participants provided sera (seroprevalence rate, %) | Risk factors for SARS-CoV-2 infections (OR, 95%CI) |
|---|---|---|---|---|
| Iversen et al., 2020 | Apr 2020 | Healthcare worker | IgG: 808/28792 (2.8)<br>IgM: 768/28792 (2.7)<br>IgG or IgM: 1163/28792 (4.0) | Male health-care workers: RR=1.49 [1.31–1.68]; p<0·001; ref: female health-care workers;<br>Frontline health-care workers: RR 1.38 [1.22–1.56]; p<0·001; ref: health-care workers in other settings;<br>Health-care workers working on dedicated COVID-19 wards: RR 1.65 [1.34–2.03]; p<0·001); ref: other frontline health-care workers |
| | | Blood donors | IgG-only: 86/4672 (1.8)<br>IgM-only: 92/4672 (2.0)<br>IgG or IgM: 142/4672 (3.0) | - |
| Olalla et al., 2020 | Apr 2020 | Health care workers | 9/498 (1.8) | - |
| Cosma et al., 2020 | Apr 2020 | Pregnant women | IgG: 8/138 (5.9)<br>IgM: 4/138 (2.9)<br>IgG and IgM: 2/138 (1.4) | - |
| Caban-Martinez et al., 2020 | Apr 2020 | Frontline firefighter/paramedic workforce" | IgG or IgM: 18/203 (8.9)<br>IgM: 10/203 (4.9)<br>IgG: 10/203 (4.9) | - |
| Poletti et al., 2020 | Jun 2020 | Close contacts of COVID-19 cases | 2187/4120 (53.1) | - |

| Reference | Starting month | Study population | No. of positive/total no. of participants provided sera (seroprevalence rate, %) | Risk factors for SARS-CoV-2 infections (OR, 95%CI) |
|---|---|---|---|---|
| Racine-Brzostek et al., 2020 | Apr 2020 | health care workers | IgG or IgM: 805/2274 (35.4)<br>IgG: 798/2274 (35.1)<br>IgM: 232/2274 (10.2) | Ancillary: 2.12; administrative staff: 2.20; ref: physicians, nurse practitioners, and physician assistants |
| Rosenberg et al., 2020 | Apr 2020 | General population | 1887/15101 (12.5) | - |
| Daniel et al., 2020 | Apr 2020 | Service member | Total antibodies: 228/382 (60.0);<br>neutralizing antibodies: 135/382 (35.3) | Univariate analysis：<br>Hispanic/Latino participants were more likely to have positive microneutralization test results than were participants of non-Hispanic/Latino or unspecified ethnicity: 2.4 (1.1-5.1). |
| Schmidt et al., 2020 | Apr 2020 | Clinic staff | 11/385 (2.9) | - |
| Moscola et al., 2020 | Mar 2020 | Health Care Personnel in the New York City Area | 5523/40329 (13.7) | A previous positive PCR test result (relative risk, 1.52 [95% CI, 1.44-1.60]; P < .001) and reported high suspicion of virus exposure (rela- tive risk, 1.23 [95% CI, 1.18-1.28]; P < .001) were associated with seroprevalence |
| Montenegro et al., 2020 | Apr 2020 | Community individuals<br><br><br>Patients consulting the primary care physician | IgG: 11/311 (3.5)<br>IgM: 12/311 (3.9)<br>IgG or IgM: 17/311 (5.5)<br>IgG or IgM: 244/634 (38.5) | - |

| Reference | Starting month | Study population | No. of positive/total no. of participants provided sera (seroprevalence rate, %) | Risk factors for SARS-CoV-2 infections (OR, 95%CI) |
|---|---|---|---|---|
| Steensels et al., 2020 | Apr 2020 | Hospital staff | 197/3056 (6.4) | Univariate analysis：<br>Having a household contact: 3.15 (2.33-4.25); ref: without any household contact |
| Soriano et al., 2020 | Apr 2020 | University employees;<br>University employees' relatives<br>Social services and health care workers<br>Individuals living in communities<br>Other | 17/175 (9.7)<br>7/85 (8.2)<br>14/108 (13.0)<br>45/234 (19.2)<br>10/72 (13.9) | - |
| Eyre et al., 2020 | Apr 2020 | Healthcare worker | IgG (CLIA): 951/9958 (9.6)<br>IgG (ELISA): 905/9958 (9.1)<br>IgG (CLIA or ELISA): 1069/9958 (10.7) | Working in Covid-19 facing areas (2.47, 1.99-3.08, p<0.001) or throughout the hospital (1.39, 1.04-1.85, p=0.02) was associated with increased risk compared to non-Covid-19 areas |
| Shields et al., 2020 | Apr 2020 | Healthcare workers | 126/516 (24.4) | Black, Asian and minority ethnic ethnicity: 1.92 (1.14-3.23). |
| Menachemi et al., 2020 | Apr 2020 | Indiana residents derived from tax returns | 38/3518(1.1)<br>52/889 (5.8) | The overall prevalence was significantly higher among Hispanics (8.3%) than among non- Hispanics (2.3%) (p = 0.03). Participants who reported having a current household member who had previously been told by a provider that they had COVID-19 had a higher overall prevalence (33.6% versus 2.2%; p = 0.004). |

| Reference | Starting month | Study population | No. of positive/total no. of participants provided sera (seroprevalence rate, %) | Risk factors for SARS-CoV-2 infections (OR, 95%CI) |
|---|---|---|---|---|
| Marina et al., 2020 | Apr 2020 | General population | LFIA: 3054/51075 (5.0)<br>CLIA: 2390/51958 (4.6) | - |
| Petersen et al., 2020 | Apr 2020 | Inhabitants of the Faroe Islands | 6/1075(0.6) | - |
| Biggs et al., 2020 | Apr 2020 | Community household residents | 19/696 (2.7) | - |
| Sydney et al., 2020 | Apr 2020 | Healthcare workers | 327/1700 (19.2) | - |
| Hunter et al., 2020 | Apr 2020 | Healthcare worker | 12/734 (1.6) | - |
| Josè et al., 2020 | May 2020 | Healthy blood donors | IgG or IgM: 9/904 (1.0);<br>IgG: 9/904 (1.0)<br>IgM: 1/904 (1.0) | - |
| Paderno et al., 2020 | Apr 2020 | Healthcare worker in otolaryngology unit | 4/58 (6.9) | - |
| Merkely et al., 2020 | May 2020 | Hungarian population | 69/10474 (0.7) | - |
| Dioscoridi et al., 2020 | May 2020 | Family members<br>health care workers | 26/81 (32.1)<br>2/38 (5.3) | - |
| Péré et al., 2020 | May 2020 | Health care workers | 437/3569 (12.2) | - |
| Torres et al., 2020 | May 2020 | Students<br>staff members | 100/1009 (9.9)<br>36/235 (15.3) | - |
| Poulikakos et al., 2020 | May 2020 | Healthcare workers | 17/281 (6.0) | - |

| Reference | Starting month | Study population | No. of positive/total no. of participants provided sera (seroprevalence rate, %) | Risk factors for SARS-CoV-2 infections (OR, 95%CI) |
|---|---|---|---|---|
| Veerus et al., 2020 | May 2020 | Pregnant women | 2/433 (0.5) | - |
| Feehan et al., 2020 | May 2020 | General population | 183/2640 (6.9) | - |
| Sutton et al., 2020 | May 2020 | Patients visiting ambulatory, emergency, or inpatient health care setting | 9/897 (1.0) | - |
| Bampoe et al., 2020 | May 2020 | Maternity healthcare workers | 29/200 (14.5) | Presence of anosmia:18 (6-55) |
| Tong et al., 2020 | May 2020 | Medical staff who went to Wuhan city for support | 0/191 (0.0) | - |
| Mughal et al., 2020 | May 2020 | Healthcare personnel (HCP) in the ICU setting. | 1/121 (0.8) | - |
| Zhang et al., 2020 | May 2020 | Close contacts of COVID-19 patients | 17/120 (14.2) | - |
| Akinbami et al., 2020 | May 2020 | Healthcare, First Response, and Public Safety Personnel | 1131/16397 (6.9) | Exposure to a household member with confirmed COVID-19: 6.18 (4.81-7.93), ref: no or unknown exposure;<br>Working within 15 km of the Detroit center: 5.60 (3.98-7.89) |
| Kempen et al., 2020 | May 2020 | Local residents | 3/99 (3.0) | - |
| Pagani et al., 2020 | May 2020 | Population of Castiglione D'Adda | 115/509 (22.6) | - |
| Blairon et al., 2020 | May 2020 | Healthcare worker | 217/1485 (14.6) | - |

| **Reference** | **Starting month** | **Study population** | **No. of positive/total no. of participants provided sera (seroprevalence rate, %)** | **Risk factors for SARS-CoV-2 infections (OR, 95%CI)** |
|---|---|---|---|---|
| Noh et al., 2020 | May 2020 | Outpatients | IgG: 1/1500 (0.1)<br>Neutralizing antibodies: 1/1500 (0.1) | - |
| Lidström et al., 2020 | May 2020 | Healthcare staff | 577/8679 (6.6) | Lower age (0.984, 0.978–0.991) and male sex (1.334, 1.104–1.612) were both associated with an increased risk of infection. |
| Haizler-Cohen et al., 2020 | May 2020 | Pregnant women | 269/1671 (16.1) | - |
| Kassem et al., 2020 | Jun 2020 | Healthcare workers employed in the gastroenterology | IgG:3/74 (4.05)<br>IgM:9/74 (12.2)<br>IgG or IgM: 9/74 (12.2) | - |
| Dimcheff et al., 2020 | Jun 2020 | Employees of a Veterans Affairs Healthcare System | 72/1476 (4.9) | Employees who reported exposure to a known COVID-19 case outside of work: 4.53 (2.67-7.68), ref: those that did not. |
| Dodd et al., 2020 | Jun 2020 | Blood donors | 4786/160328 (3.0) | Donors who were aged 55 years and older: 2.43 (1.94-3.04);<br>African American: 2.58 (1.71-3.88), Hispanic: 2.31 (1.77-3.00), ref: White donors;<br>Donors from the Northeast: 1.83 (1.57-2.12), ref: West. |
| Lundkvist et al., 2020 | Jun 2020 | Randomly selected individuals | IgG:<br>5/123 (4.1)<br>26/90 (28.9) | - |
| Younas et al., 2020 | Jun 2020 | Blood donors | 81/370 (21.9) | - |

| Reference | Starting month | Study population | No. of positive/total no. of participants provided sera (seroprevalence rate, %) | Risk factors for SARS-CoV-2 infections (OR, 95%CI) |
|---|---|---|---|---|
| Del Brutto et al., 2020 | May 2020 | Inhabitants in Atahualpa | IgG: 294/673 (43.7)<br>IgM: 256/673 (38.0)<br>IgG or IgM: 303/673 (45.0) | - |
| Preprint database | | | | |
| Sughayer et al., 2020 | Jan 2020 | Healthy blood donors | 0/746 (0) | - |
| Germain et al., 2020 | Jan 2020 | Tissue donors | ELISA:1/144 (0.7)<br>CLIA: 0/144 (0.0) | - |
| Chang et al., 2020 | Jan 2020 | Blood donors in Wuhan<br><br><br>Blood donors in Shijiazhuang<br><br><br>Blood donors in Shenzhen | Total antibodies:590/17794 (3.3);<br>Neutralizing antibodies:407/17794 (2.3)<br><br>Total antibodies:60/13540 (0.4)<br>Neutralizing antibodies:1/13540 (0.0)<br><br>Total antibodies:28/6810 (0.4)<br>Neutralizing antibodies:2/6810 (0.0) | Multivariate regression analysis revealed that age and gender were independent risk factors for the presence of antibodies against SARS-CoV-2. |
| Li et al., 2020 | Jan 2020 | Individuals with different ocular diseases | IgG or IgM:11/1331 (0.8)<br>IgM:3/1331 (0.2)<br>IgG:9/1331 (0.7) | - |
| Buss et al., 2020 | Jan 2020 | Blood donor | 1880/13867 (13.6) | - |
| Stadlbauer et al., 2020 | Feb 2020 | Patients unrelated to COVID-19 | 145/3412 (4.2) | - |

| Reference | Starting month | Study population | No. of positive/total no. of participants provided sera (seroprevalence rate, %) | Risk factors for SARS-CoV-2 infections (OR, 95%CI) |
|---|---|---|---|---|
| Xiong et al., 2020 | Feb 2020 | Healthcare workers with intensive exposure to COVID-19 | IgG: 35/797 (4.4)<br>IgM: 3/797 (0.4) | - |
| Valenti et al., 2020 | Mar 2020 | Blood donors | 40/789 (5.1) | - |
| Yu et al., 2020 | Feb 2020 | Health Care Workers | 7/337 (2.1) | - |
| Liu et al., 2020 | Feb 2020 | Healthcare providers<br>general workers<br>other patients | IgG: 153/3832 (0.4); IgM: 57/3832 (1.5)<br>IgG: 900/19555 (4.6); IgM: 254/19555 (1.3)<br>IgG: 16/1616 (1.0); IgM: 3/1616 (0.2) | - |
| Tubiana et al., 2020 | Feb 2020 | Healthcare workers | 15/147 (10.2) | - |
| Skowronski et al., 2020 | May 2020 | Anonymized residual sera were obtained from patients | Neutralizing antibodies:<br>snapshot1: 0/869 (0.2)<br>snapshot2: 4/885 (0.5) | |
| Thompson et al., 2020 | Mar 2020 | Blood donors | 5/1000 (0.5) | - |
| Dietrich et al., 2020 | Mar 2020 | Children from a Children's Hospital | 62/812 (6.3) | - |
| Brehm et al., 2020 | Mar 2020 | Health care workers<br>non-health care workers; | 9/1026 (0.9)<br>1/217 (0.4) | - |
| Tang et al., 2020 | Mar 2020 | Outpatients in Zhongnan Hospital, Wuhan University (excluding COVID-19 patients) | IgG: 145/2952(4.9)<br>IgM: 51/2952 (1.7) | - |

| Reference | Starting month | Study population | No. of positive/total no. of participants provided sera (seroprevalence rate, %) | Risk factors for SARS-CoV-2 infections (OR, 95%CI) |
|---|---|---|---|---|
| Augusto et al., 2020 | Mar 2020 | Health care workers | 15/385 (3.9) | - |
| Wang et al., 2020 | Mar 2020 | HCWs who deployed to work in Wuhan<br>HCWs who deployed to work in Wuhan<br>HCWs who deployed to work in Wuhan<br>HCWs who remained in Hefei<br>HCWs who remained in Hefei<br>HCWs who remained in Hefei | IgM:0/142 (0.0)<br>IgG: 0/142 (0.0)<br>IgA: 0/142 (0.0)<br>IgM: 0/284 (0.0)<br>IgG: 0/284 (0.0)<br>IgA: 0/284 (0.0) | - |
| Ling et al., 2020 | Mar 2020 | Back-to-work participants | IgG: 627/18391 (3.4)<br>IgM: 89/18391 (0.5)<br>IgG or IgM: 657/18391 (3.5) | - |
| Paradiso et al., 2020 | Mar 2020 | Healthcare worker | IgG:1/606 (0.2)<br>IgM:3 /606 (0.5) | - |
| Herzog et al., 2020 | Mar 2020 | Persons with blood samples collected from clinical lab | Period 1:100/3910 (2.6)<br>Period 2:193/3397 (5.7) | Increasing age, male sex, smoking, and comorbidities such as cardiovascular diseases and diabetes have been identified as risk factors for developing severe illness. |
| Dopico et al., 2020 | Mar 2020 | Blood donor and Pregnant women | 129/1900 (6.8) | - |
| Streeck et al., 2020 | Mar 2020 | Local inhabitants | IgG: 106/919 (11.5)<br>IgA: 170/919 (18.5) | - |
| Doi et al., 2020 | Mar 2020 | Patients who visited outpatient clinics with blood samples | 33/1000 (0.3) | - |

| Reference | Starting month | Study population | No. of positive/total no. of participants provided sera (seroprevalence rate, %) | Risk factors for SARS-CoV-2 infections (OR, 95%CI) |
|---|---|---|---|---|
| Tosato et al., 2020 | Apr 2020 | Healthcare professionals | IgG: 6/133 (4.5)<br>IgM: 0/133 (0.0) | - |
| Shakiba et al., 2020 | Apr 2020 | Local inhabitants | IgG: 110/528 (20.8)<br>IgM: 100/528 (19.0)<br>IgG or IgM: 117/528 (22.2) | - |
| Bastiani et al., 2020 | Apr 2020 | Adults who lived in Italy | IgG: 108/472 (22.9)<br>IgM: 49/421 (11.6) | - |
| Carozzi et al., 2020 | Apr 2020 | Health care workers | IgG: 240/17098 (1.4)<br>IgM:109/17098 (0.6) | - |
| Siddiqui et al., 2020 | Apr 2020 | Staff of a tertiary care hospital<br>individuals visiting that hospital for COVID-19 testing | 74/448 (16.5)<br>78/332 (23.5) | - |
| Kammon et al., 2020 | Apr 2020 | Community residents<br>healthcare workers | 6/142 (2.8)<br>0/77 (0.0) | - |
| Bendavid et al., 2020 | Apr 2020 | Local residents | 50/3330 (1.5) | - |
| Egerup et al., 2020 | Apr 2020 | Parturient women<br>partners of parturient women<br>newborns | 29/1313 (2.2)<br>34/1189 (2.9)<br>17/1206 (1.4) | - |
| Krähling et al., 2020 | Apr 2020 | Employees in the Frankfurt metropolitan area | 5/1000 (0.5) | - |

| Reference | Starting month | Study population | No. of positive/total no. of participants provided sera (seroprevalence rate, %) | Risk factors for SARS-CoV-2 infections (OR, 95%CI) |
|---|---|---|---|---|
| Nopsopon et al., 2020 | Apr 2020 | Hospital staff,<br>patients who needed procedural treatment or operation | IgM:<br>25/675 (3.7)<br>22/182 (12.1)<br>IgG:<br>1/675 (0.1)<br>1/182 (0.5) | Participants with present upper respiratory tract symptoms had a higher rate of positive IgM than those without (9.6% vs. 4.5%) |
| Leidner et al., 2020 | Apr 2020 | Healthcare workers | 253/10019 (2.5) | Significantly increased seropositivity among HCW age 50 and above, with odds ratio of 1.51 (95% CI 1.17-1.94) |
| Fujita et al., 2020 | Apr 2020 | Healthcare workers | 5/92 (5.4) | Univariate analysis：<br>Participants working at the otolaryngology department and/or having a history of seasonal common cold symptoms had a significantly higher titer of SARS-CoV-2 IgG antibody (p=0.046, p=0.046, respectively). |
| Psichogiou et al., 2020 | Apr 2020 | Healthcare workers from two hospitals | 15/1495 (1.0) | - |
| Thomas et al., 2020 | | Health Care Workers<br>Asymptomatic outpatients | 38/1282 (3.0)<br>106/2379 (4.5) | - |
| Cohen et al., 2020 | Apr 2020 | Children consulting an ambulatory pediatrician | 63/543 (11.6) | Contact with a person with proven COVID-19: OR 15.1 (95%CI 6.6-34.6). |

| Reference | Starting month | Study population | No. of positive/total no. of participants provided sera (seroprevalence rate, %) | Risk factors for SARS-CoV-2 infections (OR, 95%CI) |
|---|---|---|---|---|
| Rudberg et al., 2020 | Apr 2020 | Healthcare worker | 410/2149 (19.1) | Seroprevalence was strongly associated with patient-related work (OR 2.9), covid-19 patient contact (OR 1.43), and occupation assisting nurse (OR 3.67). |
| Sikora et al., 2020 | Apr 2020 | Cancer center staff | IgM: 10/161 (6.2)<br>IgG: 5/161 (3.1)<br>IgG or IgM: 12/161 (7.5) | - |
| Galán et al., 2020 | Apr 2020 | Healthcare workers wearing PPE<br>Healthcare workers with wearing PPE | 818/2590 (31.6) | Multivariate analysis:<br>Being physicians (OR 2.37, CI95% 1.61-3.49), nurses (OR 1.67, 95%CI 1.14-2.46), or nurse-assistants (OR 1.84, 95%CI 1.24-2.73), HCW working at COVID-19 hospitalization areas (OR 1.71, 95%CI 1.22-2.40), non-COVID-19 hospitalization areas (OR 1.88, 95%CI 1.30-2.73), and at the Emergency Room (OR 1.51, 95%CI 1.01-2.27). |
| Frank et al., 2020 | Apr 2020 | Residents of nursing home<br>Staff of nursing home | IgG: 15/100 (1.5); IgM: 13/100 (1.3); IgG or IgM: 17/100 (1.7)<br>IgG: 14/88 (16.0); IgM: 11/88 (12.5); IgG or IgM: 18/88 (20.5) | - |
| Garralda Fernandez et al., 2020 | Apr 2020 | Health care workers | IgG: 411/2439 (16.9)<br>IgM: 32/2439 (1.3) | - |
| Snoeck et al., 2020 | Apr 2020 | General population | IgG: 35/1820 (2.0), IgA: 201/1820 (11.0) | - |

| Reference | Starting month | Study population | No. of positive/total no. of participants provided sera (seroprevalence rate, %) | Risk factors for SARS-CoV-2 infections (OR, 95%CI) |
|---|---|---|---|---|
| Comar et al., 2020 | Apr 2020 | Healthcare worker | 52/727 (7.2) | Multivariate analysis:<br>Being medical doctor: 1.82 |
| Nisar et al., 2020 | Apr 2020 | Households | Apr:2/1000 (0.2)<br>Jun:164/1004 (16.3) | - |
| Wang et al., 2020 | Apr 2020 | Communities residents | IgG: 13/2184 (0.6)<br>IgG, IgM: 3/2184 (0.1)<br>Neutralizing antibodies: 0/2184 (0.0) | - |
| Waterfield et al., 2020 | Apr 2020 | Healthy children of healthcare workers | 68/992 (6.9%) | - |
| Zou et al., 2020 | Apr 2020 | Local residents | IgG: 3/127 (2.4)<br>IgM: 6/127 (4.7)<br>IgG or IgM: 9/127 (7.1) | - |
| Nopsopon et al., 2020 | Apr 2020 | Healthcare staff | IgG: 0/844 (0)<br>IgM: 7/844 (0.8)<br>IgG or IgM: 7/844 (0.8) | Female staff seemed to have higher rate of positive IgM (1.0%, 95% CI: 0.5%, 2.1%) than male (0.5%, 95% CI: 0.1%, 2.6%) |
| McDade et al., 2020 | Apr 2020 | Household members of essential workers | 33/202 (16.3) | - |
| Appa et al., 2020 | Apr 2020 | Residents and county essential workers | CLISA:9/1810 (0.5)<br>ELISA:4/1810 (0.2) | - |
| Tönshoff et al., 2020 | Apr 2020 | Children and their parents | IgG:70/4964 (1.4)<br>Neutralizing antibodies: 66/4964 (1.3) | - |
| Jerković et al., 2020 | Apr 2020 | Industry workers | IgG: 13/1494 (0.9)<br>IgM: 9/1494 (0.6)<br>IgG or IgM: 19/1494 (1.3) | - |

| Reference | Starting month | Study population | No. of positive/total no. of participants provided sera (seroprevalence rate, %) | Risk factors for SARS-CoV-2 infections (OR, 95%CI) |
|---|---|---|---|---|
| Alessandro et al., 2020 | Apr 2020 | Health Care Workers<br>General population | 400/2415 (16.6)<br>534/1792 (29.8) | - |
| Halatoko et al., 2020 | Apr 2020 | healthcare (doctors, nurses, pharmacy auxiliaries, hospital administrators), air transport, police, road transport (taxi and moto-taxi drivers) and informal (market sellers and craftsmen). | IgG: 8/955 (0.8)<br>IgM: 2/955 (0.2) | - |
| Dillner et al., 2020 | Apr 2020 | Healthy hospital employees | 1481/12928 (11.5) | - |
| Aziz et al., 2020 | Apr 2020 | Community residents | ELISA:46/4755 (1.0)<br>Immunofluorescent test:26/4755 (0.6)<br>Plaque reduction neutralization test:17/4755 (0.4) | - |
| Chamie et al., 2020 | Apr 2020 | All residents (>4 years) and workers in census tract | 131/3861 (3.4) | - |
| Nesbitt et al., 2020 | Apr 2020 | Blood donor | LFIA: IgM: 68/1996 (3.4); IgG: 13/1996 (0.7)<br>CLIA:14/1996(0.7) | - |
| Wells et al., 2020 | Apr 2020 | Members of the Twins K cohort | 51/431 (11.8) | Seropositive participants were older (median age seropositive 48, median age seronegative 36; p = 0.046). No difference in sex (% female of seropositive participants 72, and 87 for seronegative) or BMI (median 23.8 seropositive; 22.8 seronegative) was evident between the groups. |
| Fontanet et al., 2020 | Apr 2020 | Pupils, their parents and relatives, and staff of primary schools | 139/1340 (10.4) | - |

| Reference | Starting month | Study population | No. of positive/total no. of participants provided sera (seroprevalence rate, %) | Risk factors for SARS-CoV-2 infections (OR, 95%CI) |
|---|---|---|---|---|
| Sandri et al., 2020 | Apr 2020 | Healthcare workers | 447/3985 (11.2) | - |
| Uyoga et al., 2020 | Apr 2020 | Blood donors | 174/3098 (5.6) | - |
| Brant-Zawadzki et al., 2020 | May 2020 | Healthcare workers | 31/2932 (1.1) | Significant differences between observed negative and positive cases were found for age (z = 2.65, p = 0.008), race (p = 0.037), presence of fever (p < 0.001), and loss of smell (p < 0.001) |
| Addetia et al., 2020 | May 2020 | Ship's crew | CLIA: 6/120 (5.0)<br>Neutralization assay: 3/120 (2.5) | - |
| Barallat et al., 2020 | May 2020 | Healthcare worker | IgG (S1/S2): 712/7563 (9.4)<br>IgG (S1/S2 or N): 779/7563 (10.3) | - |
| Tess et al., 2020 | May 2020 | Local inhabitants | IgG: 21/517 (4.1)<br>IgM: 7/517 (1.4)<br>IgG or IgM: 27/517 (5.2) | - |
| Takita et al., 2020 | May 2020 | Inhabitants | 115/509 (22.6) | Univariate analysis：<br>The central Tokyo of 23 special wards exhibited a significantly higher prevalence compared to the other area of Tokyo (p =0.02, 4.68% (95%CI: 3.08-6.79) versus 1.83 (0.68-3.95) in central and suburban Tokyo. |
| Mattern et al., 2020 | May 2020 | All patients admitted to the delivery room | 20/249 (8.0) | - |

| Reference | Starting month | Study population | No. of positive/total no. of participants provided sera (seroprevalence rate, %) | Risk factors for SARS-CoV-2 infections (OR, 95%CI) |
|---|---|---|---|---|
| Carrat et al., 2020 | May 2020 | General adult population | IgG-SP: 983/14628 (6.7)<br>IgG-NP: 511/14628 (3.5)<br>Neutralizing:424/14628 (2.90%) | - |
| McBride et al., 2020 | May 2020 | Outpatients coming into the Department of Radiation Oncology | 44/919 (4.8) | - |
| Ebinger et al., 2020 | May 2020 | Health Care Workers | 212/6062 (3.5) | The strongest self-reported symptom associated with greater odds of seropositive status was anosmia (11.53 [7.51, 17.70], P<0.001) |
| Hurk et al., 2020 | May 2020 | Blood donor | 419/7150 (5.9) | - |
| Weis et al., 2020 | May 2020 | Community residents | 52/620 (8.4) | - |
| Rigatti et al., 2020 | May 2020 | Life insurance applicants | 1520/50025 (3.0) | - |
| Gomes et al., 2020 | May 2020 | Maternity healthcare workers | 97/4608 (2.1) | - |
| Hallal et al., 2020 | Apr 2020 | Community residents | 347/24955 (1.4)<br>746/31128 (2.4) | Indigenous individuals: 5.89 (95%CI 2.99-10.66) ref: the white. |
| Nakamura et al., 2020 | May 2020 | Healthcare workers | CLIA (Abbott): 4/1000 (0.4); CLIA (Roche): 0/1000 (0.0); POC qualitative test: 33/1000 (3.3) | - |
| Jespersen et al., 2020 | May 2020 | All healthcare workers and administrative personnel at the hospitals (including the pre-hospital services) and specialist practitioner clinics) | 668/17948 (3.7) | Nursing staff (7.3, 3.5–14.9), medical doctors (4.0, 1.8–8.9), and biomedical laboratory (5.0, 2.1–11.6) scientists; ref: medical secretaries |

| Reference | Starting month | Study population | No. of positive/total no. of participants provided sera (seroprevalence rate, %) | Risk factors for SARS-CoV-2 infections (OR, 95%CI) |
|---|---|---|---|---|
| Tsertsvadze et al., 2020 | May 2020 | Adult residents of capital city of Tbilisi | 9/1068 (0.8) | - |
| Chibwana et al., 2020 | May 2020 | Health care workers | 84/500 (16.8) | - |
| Armann et al., 2020 | May 2020 | Students and their teacher without household contacts of COVID-19 patients;<br>Students and their teacher with household contacts of COVID-19 patients | 17/2016 (0.8)<br>1/24 (4.2) | - |
| Hibino et al., 2020 | May 2020 | Healthy volunteers working for a Japanese company | IgG: 95/350 (27.1)<br>IgM: 90/350 (25.7) | - |
| Wilkins et al., 2020 | May 2020 | Healthcare workers<br>. | 316/6510 (4.9) | Known out-of-hospital exposure was 4.7 (3.5-6.4), ref: without out-of-hospital exposure; Participants with a family member who tested positive for COVID-19: 26.8 (17.3-41.8), ref: Participants without a positive family; Services (3.0, 1.2-6.4); medical assistants (2.9, 1.4-5.5); nurses (2.12, 1.5-3.2) had higher odds: ref : administrators;<br>Participating in the care of COVID-19 patients: 2.19 (1.61-3.01), ref: participants who did not report participating in the care of COVID-19 patients. |
| Alkurt et al., 2020 | May 2020 | Healthcare workers | 22/813 (2.7) | - |
| Vassallo et al., 2020 | May 2020 | Blood Donors | 2948/189656 (1.6) | - |

| Reference | Starting month | Study population | No. of positive/total no. of participants provided sera (seroprevalence rate, %) | Risk factors for SARS-CoV-2 infections (OR, 95%CI) |
|---|---|---|---|---|
| Melo et al., 2020 | May 2020 | Healthcare workers | IgM: 28/471 (5.9)<br>IgG: 64/471 (13.6) | - |
| Favara et al., 2020 | May 2020 | Hospital staff working in an oncology department | Rapid POC serology: 34/434 (7.8)<br>Microsphere-based assay: 80/434 (18.4) | - |
| Silva et al., 2020 | May 2020 | Health care workers from public facilities | 5/738 (0.7) | - |
| Ray et al., 2020 | May 2020 | Patients who were admitted to the medicine wards and intensive care unit (ICU) | 42/212 (19.8) | - |
| Bardai et al., 2020 | May 2020 | children patients<br>accompanying persons<br>hospital employees | 3/39 (7.7)<br>7/61 (11.5)<br>12/99 (12.1) | - |
| Mahajan et al., 2020 | Jun 2020 | Community residents | 23/567 (4.1) | - |
| Nishida et al., 2020 | Jun 2020 | Hospital staff | 4/925 (0.4) | - |
| Nawa et al., 2020 | Jun 2020 | Households randomly selected from Utsunomiya City's basic resident registry | 3/742 (0.7) | - |
| Qutob et al., 2020 | Jun 2020 | Palestinian population residing in the West Bank<br>individuals visiting medical laboratories | 0/1319 (0.0)<br>4/1136 (0.4) | - |
| Ulyte et al., 2020 | Jun 2020 | School children | 74/2484 (3.0) | - |
| Asuquo et al., 2020 | Jun 2020 | clinic staff and patients | 17/66 (25.8) | - |
| Ward et al., 2020 | Jun 2020 | Community adults | 5544/99908 (5.6) | - |

| Reference | Starting month | Study population | No. of positive/total no. of participants provided sera (seroprevalence rate, %) | Risk factors for SARS-CoV-2 infections (OR, 95%CI) |
|---|---|---|---|---|
| Menezes et al., 2020 | Jun 2020 | Community residents | 849/31869 (2.7) | - |
| Ariza et al., 2020 | Apr 2020 | medical trainees or medical doctors | 8/351 (2.3) | - |
| Majiya et al., 2020 | Jun 2020 | Residents | IgG: 47/185 (25.4)<br>IgM:4/185 (2.2) | |
| Javed et al., 2020 | Jun 2020 | Working population | IgG:2543/24210 (10.5)<br>IgM:2783/24210 (11.5)<br>IgG, IgM:4234/24210 (17.5) | - |
| Buonsenso et al., 2020 | Jun 2020 | Household contacts of index patients | 44/80 (55.0) | - |
| Khan et al., 2020 | Jun 2020 | Hospital visitors | 111/2906 (3.8) | - |
| Satpati et al., 2020 | Jun 2020 | Population of Paschim Medinipur District | 19/458 (4.2) | - |
| Silva et al., 2020 | Jun 2020 | Residents | 1167/3156 (37.0) | - |
| Calife et al., 2020 | Jun 2020 | Residents | 33/2342 (1.4) | - |
| Kumar et al., 2020 | Jun 2020 | Healthcare worker | CLIA: 14/996 (1.4)<br>ELISA: 22/996 (2.2) | - |
| Offical report | | | | |
| Public Health Ontario, Canada, 2020 | Mar 2020 | Serum or plasma left over after diagnostic testing | March 2020: 3/827 (0.4)<br>May 2020: 15/1061 (1.4)<br>June 2020: 79/7014 (1.1) | - |

| Reference | Starting month | Study population | No. of positive/total no. of participants provided sera (seroprevalence rate, %) | Risk factors for SARS-CoV-2 infections (OR, 95%CI) |
|---|---|---|---|---|
| Office of National Statistics, UK, 2020 | Apr 2020 | General population | 153/3298 (6.3) | - |
| the Government of Jersey, UK, 2020 | Apr 2020 | Adult resident population living in private households in Jersey | 24/855 (2.9) | - |
| Canadian Blood Services, 2020 | May 2020 | Blood donor | 275/37737 (0.7) | - |
| Ministry of Health, Labour and Welfare, Japan, 2020 | Jun 2020 | Residents | 2/1971 (0.1)<br>5/2970 (0.2)<br>1/3009 (0.0) | - |
| Islamic Republic of Afghanistan Ministry of Public Health, Afghanistan, 2020 | Jul 2020 | General population | 2997/9514 (31.5) | |
| Public Health England, 2020 | Aug 2020 | Blood donor | 457/7899 (5.8) | - |
| MedLife, Romania, 2020 | | Healthcare workers | 11/371 (3.0) | - |

Abbreviations: ELISA, Enzyme-linked immunosorbent assay; CLIA: Chemiluminescent immunoassay; LFIA: lateral flow immunoassays; MIA: Microsphere immunoassay; MN, Microneutralisation assay; POC: point of care;

* The sensitivity and specificity validated by the authors rather than manufactures.

**Appendix Table 5. Definition of subjects included in meta-analysis**

| Type of exposure | Population | Definition |
|---|---|---|
| Exposed to laboratory-confirmed or suspected COVID-19 patients | Close contact | A person or a group of people who lived with or cared for a virologically-confirmed or suspected COVID-19 patients during the infectious period (e.g. household members, family contacts and relatives.), as well as other persons who worked with or had close contact with the virologically-confirmed or suspected COVID-19 patients during the infectious period (e.g. office co-workers, people sharing same waiting room, service member in the same aircraft carrier, patients in the same hemodialysis unit, and other potential social contacts). Specifically, clustering cases (excluding the patient) in the community or working place were also considered as close contacts. |
| | High-risk healthcare worker | A group of persons who provided routine medical care for virologically-confirmed or suspected COVID-19 patients during the infectious period without wearing personal protective equipment (including protective suits, mask, gloves, goggles, face shields, and gowns). |
| Exposed to laboratory-confirmed /suspected/non COVID-19 patients | Low-risk healthcare worker | A group of persons who provided routine medical care for virologically-confirmed or suspected COVID-19 patients during the infectious period with the use of personal protective equipment (including protective suits, mask, gloves, goggles, face shields, and gowns), as well as those people who provided medical care for non-COVID-19 patients. |
| Without known exposure to laboratory-confirmed or suspected COVID-19 patients | General population | Persons without known exposure to laboratory-confirmed or suspected COVID-19 patients (e.g. community residents). |
| Indeterminate exposure to laboratory- | Poorly-defined population | Persons with undefined or unknown exposure to laboratory-confirmed or suspected COVID-19 patients, as well as |

| | | |
|---|---|---|
| confirmed or suspected COVID-19 patients | | those participants cannot be categorized as the study populations mentioned above due to limited exposure information. |

**Appendix Table 6. Scoring system used for evaluation of published reports describing seroevidence of human infection with SARS-CoV-2**

| | Parameter | Maximum score | Individual score | | | |
|---|---|---|---|---|---|---|
| | | | 0 | 1 | 2 | 3 |
| Study design | Representativeness of samples | 3 | Without reporting the method of recruitment of study participants or the selection of study sites | Convenience samples without randomly selecting study participants (e.g. archived specimens from clinical labs, or healthcare workers in single center) | Randomly-selected samples in communities or multiple healthcare settings | Multi-stage/stratified samples from communities or universal samples from healthcare settings |
| Laboratory method | Approval by National Regulatory Authority | 1 | No | Yes | NA | NA |
| | Validation prior to assay for surveillance | 2 | No | NA | Yes | NA |
| | Confirmation methods | 2 | No | Second serological assay (except the VNT or pVNT) | VNT or pVNT | NA |
| Outcomes | Correction for age or sex* | 2 | No | NA | Yes | NA |

| | | | | | | |
|---|---|---|---|---|---|---|
| | Correction for testing performance (sensitivity and specificity) | 2 | No | NA | Yes | NA |
| | Total | 12 | NA | NA | NA | NA |

Note: VNT, Virus neutralization tests (such as the plaque-reduction neutralization test (PRNT) and microneutralization); pVNT, Pseudovirus neutralization tests;

* Studies stratified their findings in separate age groups or sex will be assigned with 2 points.

**Appendix Table 7. Quality assessment of serological studies describing subclinical and clinically mild human infections with SARS-CoV-2**

| Reference | Study characteristics | | Laboratory method | | | Outcome | | Total | Grade |
|---|---|---|---|---|---|---|---|---|---|
| | Study population | Representativeness of samples | Approval for NRA | pre-experiment validations | Confirmation methods | Correction for age/sex or other socio-demographic factors | Correction for test performance | | |
| Peer-reviewed databases | | | | | | | | | |
| Victoria et al., 2020 | Close contacts; Low-risk healthcare workers | 1 | 1 | 2 | 0 | 0 | 0 | 4 | C |
| To et al., 2020 | Poorly-defined population | 1 | 0 | 2 | 2 | 2 | 0 | 7 | B |
| Liang et al., 2020 | Poorly-defined population | 2 | 1 | 0 | 0 | 2 | 0 | 5 | C |
| Hallowell et al., 2020 | Close contacts; Poorly-defined population | 1 | 1 | 0 | 2 | 0 | 0 | 4 | C |
| Sam et al., 2020 | Poorly-defined population | 1 | 1 | 2 | 1 | 2 | 0 | 7 | B |
| Chen et al., 2020 | High-risk healthcare workers | 1 | 1 | 2 | 2 | 2 | 0 | 8 | B |
| Cavicchiolo et al., 2020 | Poorly-defined population | 1 | 1 | 0 | 0 | 0 | 0 | 2 | D |
| Plebani et al., 2020 | Poorly-defined population | 2 | 1 | 2 | 0 | 2 | 0 | 7 | B |
| Cox et al., 2020 | Close contacts | 1 | 1 | 0 | 0 | 0 | 0 | 2 | D |
| Brandstetter et al., 2020 | High-risk healthcare workers | 1 | 1 | 0 | 0 | 0 | 0 | 2 | D |

| | | | | | | | | | |
|---|---|---|---|---|---|---|---|---|---|
| Solodky et al., 2020 | Low-risk healthcare workers; Poorly-defined population | 1 | 0 | 0 | 0 | 0 | 0 | 1 | D |
| Zhang et al., 2020 | Poorly-defined population | 1 | 0 | 0 | 0 | 0 | 0 | 1 | D |
| Suda et al., 2020 | Poorly-defined population | 1 | 1 | 2 | 1 | 0 | 0 | 5 | C |
| Bogogiannidou et al., 2020 | Poorly-defined population | 1 | 1 | 2 | 1 | 2 | 2 | 9 | B |
| Xu et al., 2020 | Poorly-defined population | 2 | 1 | 2 | 0 | 2 | 0 | 7 | B |
| Vena et al., 2020 | Poorly-defined population | 1 | 1 | 2 | 1 | 2 | 0 | 7 | B |
| Ng et al., 2020 | Poorly-defined population | 1 | 1 | 2 | 2 | 0 | 0 | 6 | C |
| Dingens et al., 2020 | Poorly-defined population | 1 | 1 | 0 | 2 | 2 | 0 | 6 | C |
| Fischer et al., 2020 | Poorly-defined population | 1 | 1 | 0 | 1 | 0 | 0 | 3 | D |
| Brown et al., 2020 | Close contacts | 1 | 1 | 0 | 0 | 0 | 0 | 2 | D |
| Han et al., 2020 | General population | 1 | 1 | 0 | 0 | 0 | 0 | 2 | D |
| De et al., 2020 | Poorly-defined population | 0 | 0 | 0 | 0 | 0 | 0 | 0 | D |
| Zhou et al., 2020 | Low-risk healthcare workers | 1 | 1 | 0 | 0 | 2 | 0 | 4 | C |

| | | | | | | | | | |
|---|---|---|---|---|---|---|---|---|---|
| Tu et al., 2020 | Low-risk healthcare workers; Poorly-defined population | 1 | 1 | 0 | 1 | 0 | 0 | 3 | D |
| Fuereder et al., 2020 | Poorly-defined population | 1 | 1 | 0 | 1 | 0 | 0 | 3 | D |
| Fusco et al., 2020 | Low-risk healthcare workers | 1 | 1 | 0 | 0 | 2 | 0 | 4 | C |
| Havers et al., 2020 | Poorly-defined population | 1 | 1 | 0 | 1 | 2 | 2 | 7 | B |
| Xu et al., 2020 | Poorly-defined population | 1 | 1 | 0 | 1 | 0 | 0 | 3 | D |
| Behrens et al., 2020 | Low-risk healthcare workers | 1 | 1 | 0 | 2 | 0 | 0 | 4 | C |
| Loconsole et al., 2020 | Poorly-defined population | 1 | 1 | 0 | 0 | 0 | 0 | 2 | D |
| Mansour et al., 2020 | Low-risk healthcare workers | 1 | 1 | 0 | 0 | 2 | 0 | 4 | C |
| Gallian et al., 2020 | Poorly-defined population | 1 | 1 | 2 | 2 | 0 | 0 | 6 | C |
| Korth et al., 2020 | Low-risk healthcare workers | 1 | 1 | 0 | 0 | 0 | 0 | 2 | D |
| Bielecki et al., 2020 | Close contacts; Poorly-defined population | 1 | 1 | 2 | 0 | 0 | 0 | 4 | C |
| Tsaneva et al., 2020 | Poorly-defined population | 1 | 1 | 0 | 0 | 2 | 0 | 4 | C |
| Houlihan et al., 2020 | Low-risk healthcare workers | 1 | 0 | 0 | 1 | 0 | 0 | 2 | D |

| | | | | | | | | | |
|---|---|---|---|---|---|---|---|---|---|
| Basteiro et al., 2020 | Low-risk healthcare workers | 2 | 1 | 2 | 0 | 2 | 0 | 7 | B |
| Isherwood et al., 2020 | Low-risk healthcare workers; Poorly-defined population | 1 | 1 | 0 | 0 | 2 | 0 | 4 | C |
| Xu et al., 2020 | Low-risk healthcare workers; General population; Poorly-defined population | 1 | 1 | 2 | 0 | 0 | 0 | 4 | C |
| Milani et al., 2020 | Poorly-defined population | 1 | 1 | 0 | 0 | 0 | 0 | 2 | D |
| Bryan et al., 2020 | General population | 1 | 1 | 2 | 0 | 2 | 0 | 6 | C |
| Hains et al., 2020 | Close contacts; Low-risk healthcare workers | 1 | 1 | 0 | 1 | 2 | 0 | 5 | C |
| Liu et al., 2020 | Low-risk healthcare workers | 1 | 1 | 2 | 0 | 2 | 0 | 6 | C |
| Malickova et al., 2020 | Poorly-defined population | 1 | 1 | 0 | 0 | 0 | 0 | 2 | D |
| Lackermair et al., 2020 | Low-risk healthcare workers | 1 | 1 | 0 | 0 | 0 | 0 | 2 | D |
| Sotgiu et al., 2020 | Poorly-defined population | 1 | 1 | 0 | 0 | 2 | 0 | 4 | C |
| Wu et al., 2020 | Poorly-defined population | 1 | 1 | 0 | 0 | 0 | 0 | 2 | D |
| Stubblefield et al., 2020 | Low-risk healthcare workers | 1 | 1 | 0 | 0 | 2 | 0 | 4 | C |

| | | | | | | | | | |
|---|---|---|---|---|---|---|---|---|---|
| Self et al., 2020 | Low-risk healthcare workers | 2 | 1 | 2 | 0 | 2 | 0 | 7 | B |
| Patel et al., 2020 | Poorly-defined population | 1 | 1 | 0 | 0 | 0 | 0 | 2 | D |
| Flannery et al., 2020 | Poorly-defined population | 1 | 1 | 2 | 0 | 0 | 0 | 4 | C |
| Stock et al., 2020 | Low-risk healthcare workers | 1 | 1 | 0 | 0 | 2 | 0 | 4 | C |
| Goldberg et al., 2020 | Poorly-defined population | 1 | 1 | 0 | 0 | 0 | 0 | 2 | D |
| Stringhini et al., 2020 | General population | 3 | 1 | 2 | 1 | 2 | 2 | 11 | A |
| Erikstrup et al., 2020 | Poorly-defined population | 1 | 1 | 2 | 0 | 2 | 2 | 8 | B |
| Lahner et al., 2020 | Low-risk healthcare workers | 1 | 1 | 2 | 0 | 0 | 0 | 4 | C |
| Pallett et al., 2020 | Poorly-defined population | 2 | 1 | 2 | 1 | 0 | 0 | 6 | C |
| Sood et al., 2020 | General population | 3 | 1 | 2 | 0 | 2 | 2 | 10 | A |
| Madsen T et al., 2020 | Low-risk healthcare workers | 1 | 1 | 2 | 0 | 0 | 0 | 4 | C |
| Crovetto et al., 2020 | Poorly-defined population | 0 | 1 | 0 | 1 | 0 | 0 | 2 | D |
| Gudbjartsson et al., 2020 | General population; Low-risk healthcare workers | 2 | 1 | 2 | 1 | 2 | 0 | 8 | B |

| | | | | | | | | | |
|---|---|---|---|---|---|---|---|---|---|
| Naranbhai et al., 2020 | General population | 1 | 1 | 2 | 0 | 2 | 2 | 8 | B |
| Martin et al., 2020 | Low-risk healthcare workers | 1 | 1 | 0 | 0 | 2 | 0 | 4 | C |
| Amendola et al., 2020 | Low-risk healthcare workers | 1 | 1 | 0 | 0 | 2 | 0 | 4 | C |
| Iversen et al., 2020 | Low-risk healthcare workers; Poorly-defined population | 3 | 1 | 2 | 0 | 2 | 2 | 10 | A |
| Olalla et al., 2020 | Low-risk healthcare workers | 1 | 1 | 0 | 0 | 0 | 0 | 2 | D |
| Cosma et al., 2020 | Poorly-defined population | 1 | 1 | 0 | 1 | 0 | 0 | 3 | D |
| Caban-Martinez et al., 2020 | Poorly-defined population | 1 | 1 | 0 | 0 | 2 | 0 | 4 | C |
| Poletti et al., 2020 | Close contacts | 1 | 1 | 0 | 0 | 2 | 0 | 4 | C |
| Racine-Brzostek et al., 2020 | Poorly-defined population | 1 | 1 | 0 | 0 | 2 | 0 | 4 | C |
| Rosenberg et al., 2020 | General population | 1 | 1 | 2 | 0 | 2 | 2 | 8 | B |
| Daniel et al., 2020 | Close contacts | 1 | 1 | 0 | 2 | 0 | 0 | 4 | C |
| Schmidt et al., 2020 | Low-risk healthcare workers | 1 | 1 | 0 | 0 | 2 | 0 | 4 | C |
| Moscola et al., 2020 | Low-risk healthcare workers | 3 | 1 | 2 | 1 | 2 | 0 | 9 | B |

| | | | | | | | | | |
|---|---|---|---|---|---|---|---|---|---|
| Montenegro et al., 2020 | General population; Poorly-defined population | 2 | 1 | 0 | 1 | 2 | 0 | 6 | C |
| Steensels et al., 2020 | Low-risk healthcare workers | 1 | 1 | 2 | 0 | 0 | 0 | 4 | C |
| Soriano et al., 2020 | Close contacts; Poorly-defined population | 1 | 1 | 0 | 0 | 0 | 0 | 2 | D |
| Eyre et al., 2020 | Low-risk healthcare workers | 2 | 1 | 0 | 1 | 0 | 0 | 4 | C |
| Shields et al., 2020 | Low-risk healthcare workers | 1 | 0 | 0 | 0 | 2 | 0 | 3 | D |
| Menachemi et al., 2020 | General population | 3 | 0 | 0 | 0 | 2 | 0 | 5 | C |
| Marina et al., 2020 | General population | 3 | 1 | 2 | 1 | 2 | 0 | 9 | B |
| Petersen et al., 2020 | General population | 2 | 1 | 0 | 0 | 2 | 2 | 7 | B |
| Biggs et al., 2020 | General population | 3 | 1 | 2 | 0 | 2 | 0 | 8 | B |
| Sydney et al., 2020 | Poorly-defined population | 1 | 1 | 0 | 0 | 0 | 0 | 2 | D |
| Hunter et al., 2020 | Low-risk healthcare workers | 2 | 1 | 0 | 0 | 2 | 0 | 5 | C |
| Josè et al., 2020 | Poorly-defined population | 1 | 1 | 0 | 0 | 2 | 0 | 4 | C |
| Paderno et al., 2020 | Low-risk healthcare workers | 1 | 1 | 0 | 0 | 0 | 0 | 2 | D |

| | | | | | | | | | |
|---|---|---|---|---|---|---|---|---|---|
| Merkely et al., 2020 | General population | 3 | 1 | 0 | 0 | 2 | 0 | 6 | C |
| Dioscoridi et al., 2020 | Close contacts; Low-risk healthcare workers | 1 | 1 | 0 | 1 | 0 | 0 | 3 | D |
| Péré et al., 2020 | Poorly-defined population | 1 | 1 | 2 | 1 | 0 | 0 | 5 | C |
| Torres et al., 2020 | Close contacts | 2 | 1 | 0 | 0 | 2 | 0 | 5 | C |
| Poulikakos et al., 2020 | Low-risk healthcare workers | 1 | 1 | 0 | 0 | 0 | 0 | 2 | D |
| Veerus et al., 2020 | Poorly-defined population | 1 | 1 | 0 | 0 | 2 | 0 | 4 | C |
| Feehan et al., 2020 | General population | 3 | 1 | 0 | 0 | 2 | 0 | 6 | C |
| Sutton et al., 2020 | Poorly-defined population | 1 | 1 | 0 | 0 | 2 | 0 | 4 | C |
| Bampoe et al., 2020 | High-risk healthcare workers | 2 | 1 | 0 | 0 | 0 | 0 | 3 | D |
| Tong et al., 2020 | Low-risk healthcare workers | 1 | 1 | 0 | 0 | 2 | 0 | 4 | C |
| Mughal et al., 2020 | Low-risk healthcare workers | 1 | 1 | 0 | 0 | 0 | 0 | 2 | D |
| Zhang et al., 2020 | Close contacts | 2 | 1 | 0 | 0 | 0 | 0 | 3 | D |
| Akinbami et al., 2020 | Poorly-defined population | 1 | 1 | 0 | 0 | 2 | 0 | 4 | C |

| | | | | | | | | | |
|---|---|---|---|---|---|---|---|---|---|
| Kempen et al., 2020 | General population | 1 | 1 | 0 | 0 | 2 | 0 | 4 | C |
| Pagani et al., 2020 | General population | 2 | 1 | 0 | 0 | 2 | 0 | 5 | C |
| Blairon et al., 2020 | Poorly-defined population | 2 | 1 | 2 | 1 | 0 | 0 | 6 | C |
| Noh et al., 2020 | Poorly-defined population | 1 | 1 | 0 | 2 | 0 | 0 | 4 | C |
| Lidström et al., 2020 | Low-risk healthcare workers | 3 | 1 | 0 | 0 | 2 | 0 | 6 | C |
| Haizler-Cohen et al., 2020 | Poorly-defined population | 2 | 1 | 0 | 0 | 0 | 0 | 3 | D |
| Kassem et al., 2020 | Low-risk healthcare workers | 1 | 1 | 0 | 0 | 0 | 0 | 2 | D |
| Dimcheff et al., 2020 | Low-risk healthcare workers | 1 | 1 | 0 | 0 | 2 | 0 | 4 | C |
| Dodd et al., 2020 | Poorly-defined population | 1 | 1 | 0 | 0 | 2 | 0 | 4 | C |
| Lundkvist et al., 2020 | General population | 2 | 1 | 2 | 0 | 0 | 0 | 5 | C |
| Younas et al., 2020 | Poorly-defined population | 1 | 1 | 2 | 1 | 0 | 0 | 5 | C |
| Del Brutto et al., 2020 | General population | 3 | 1 | 0 | 0 | 2 | 0 | 6 | C |
| Preprint servers | | | | | | | | | |
| Sughayer et al., 2020 | Poorly-defined population | 1 | 1 | 0 | 0 | 0 | 0 | 2 | D |

| | | | | | | | | | |
|---|---|---|---|---|---|---|---|---|---|
| Germain et al., 2020 | Poorly-defined population | 1 | 1 | 0 | 1 | 0 | 0 | 3 | D |
| Chang et al., 2020 | Poorly-defined population | 1 | 1 | 0 | 2 | 2 | 0 | 6 | C |
| Li et al., 2020 | Poorly-defined population | 1 | 1 | 0 | 0 | 0 | 0 | 2 | D |
| Buss et al., 2020 | Poorly-defined population | 1 | 1 | 2 | 0 | 2 | 2 | 8 | B |
| Stadlbauer et al., 2020 | Poorly-defined population | 1 | 1 | 2 | 1 | 0 | 0 | 5 | C |
| Xiong et al., 2020 | Low-risk healthcare workers | 0 | 0 | 0 | 0 | 0 | 0 | 0 | D |
| Valenti et al., 2020 | Poorly-defined population | 1 | 1 | 2 | 0 | 2 | 2 | 8 | B |
| Yu et al., 2020 | Low-risk healthcare workers | 1 | 1 | 0 | 0 | 0 | 0 | 2 | D |
| Liu et al., 2020 | High-risk healthcare workers; General population; Poorly-defined population | 1 | 1 | 0 | 0 | 2 | 0 | 4 | C |
| Tubiana et al., 2020 | High-risk healthcare workers | 2 | 1 | 0 | 1 | 2 | 0 | 6 | C |
| Skowronski et al., 2020 | Poorly-defined population | 1 | 1 | 0 | 2 | 2 | 2 | 8 | B |
| Thompson et al., 2020 | Poorly-defined population | 1 | 1 | 2 | 2 | 0 | 0 | 6 | C |

| | | | | | | | | | |
|---|---|---|---|---|---|---|---|---|---|
| Dietrich et al., 2020 | Poorly-defined population | 1 | 1 | 2 | 0 | 2 | 0 | 6 | C |
| Brehm et al., 2020 | Low-risk healthcare workers | 1 | 1 | 2 | 0 | 0 | 0 | 4 | C |
| Tang et al., 2020 | Poorly-defined population | 1 | 1 | 0 | 0 | 2 | 0 | 4 | C |
| Augusto et al., 2020 | Low-risk healthcare workers | 2 | 1 | 0 | 0 | 0 | 0 | 3 | D |
| Wang et al., 2020 | Low-risk healthcare workers | 1 | 1 | 0 | 0 | 2 | 0 | 4 | C |
| Ling et al., 2020 | General population | 1 | 1 | 2 | 0 | 2 | 2 | 8 | B |
| Paradiso et al., 2020 | Poorly-defined population | 1 | 1 | 0 | 1 | 0 | 0 | 3 | D |
| Herzog et al., 2020 | Poorly-defined population | 1 | 1 | 0 | 0 | 2 | 0 | 4 | C |
| Dopico et al., 2020 | Poorly-defined population | 1 | 1 | 2 | 0 | 0 | 0 | 4 | C |
| Streeck et al., 2020 | General population | 2 | 1 | 2 | 2 | 0 | 2 | 9 | B |
| Doi et al., 2020 | Poorly-defined population | 1 | 0 | 0 | 0 | 2 | 0 | 3 | D |
| Tosato et al., 2020 | Low-risk healthcare workers | 1 | 1 | 0 | 0 | 0 | 0 | 2 | D |
| Shakiba et al., 2020 | General population | 3 | 1 | 2 | 0 | 2 | 2 | 10 | A |

| | | | | | | | | | |
|---|---|---|---|---|---|---|---|---|---|
| Bastiani et al., 2020 | Poorly-defined population | 1 | 0 | 0 | 0 | 2 | 0 | 3 | D |
| Carozzi et al., 2020 | Low-risk healthcare workers | 2 | 1 | 2 | 1 | 0 | 0 | 6 | C |
| Siddiqui et al., 2020 | Low-risk healthcare workers; Poorly-defined population | 1 | 1 | 0 | 0 | 2 | 0 | 4 | C |
| Kammon et al., 2020 | Low-risk healthcare workers; General population | 2 | 1 | 0 | 0 | 2 | 0 | 5 | C |
| Bendavid et al., 2020 | General population | 1 | 1 | 2 | 0 | 2 | 2 | 8 | B |
| Egerup et al., 2020 | Poorly-defined population | 1 | 1 | 0 | 0 | 2 | 2 | 6 | C |
| Krähling et al., 2020 | Poorly-defined population | 1 | 0 | 2 | 2 | 0 | 0 | 5 | C |
| Nopsopon et al., 2020 | Low-risk healthcare workers; Poorly-defined population | 3 | 1 | 2 | 0 | 2 | 0 | 8 | B |
| Leidner et al., 2020 | Low-risk healthcare workers | 2 | 1 | 2 | 0 | 0 | 0 | 5 | C |
| Fujita et al., 2020 | Low-risk healthcare workers | 1 | 1 | 0 | 0 | 0 | 0 | 2 | D |
| Psichogiou et al., 2020 | Low-risk healthcare workers | 1 | 0 | 2 | 0 | 2 | 0 | 5 | C |
| Thomas et al., 2020 | Close contacts; Low-risk healthcare workers | 2 | 0 | 2 | 0 | 0 | 0 | 4 | C |

| | | | | | | | | | |
|---|---|---|---|---|---|---|---|---|---|
| Cohen et al., 2020 | Close contacts; Poorly-defined population | 2 | 1 | 0 | 0 | 0 | 0 | 3 | D |
| Rudberg et al., 2020 | Low-risk healthcare workers | 1 | 1 | 2 | 0 | 2 | 0 | 6 | C |
| Sikora et al., 2020 | Poorly-defined population | 1 | 1 | 0 | 0 | 0 | 0 | 2 | D |
| Galán et al., 2020 | High-risk healthcare workers | 1 | 1 | 0 | 0 | 2 | 0 | 4 | C |
| Frank et al., 2020 | Poorly-defined population | 1 | 1 | 0 | 0 | 0 | 0 | 2 | D |
| Garralda et al., 2020 | Low-risk healthcare workers | 1 | 1 | 0 | 0 | 2 | 0 | 4 | C |
| Snoeck et al., 2020 | General population | 3 | 1 | 2 | 0 | 2 | 0 | 8 | B |
| Comar et al., 2020 | Low-risk healthcare workers | 1 | 1 | 2 | 0 | 0 | 0 | 4 | C |
| Nisar et al., 2020 | General population | 2 | 1 | 2 | 0 | 2 | 0 | 7 | B |
| Wang et al., 2020 | General population | 3 | 1 | 2 | 2 | 2 | 0 | 10 | A |
| Waterfield et al., 2020 | Poorly-defined population | 2 | 1 | 0 | 1 | 2 | 0 | 6 | C |
| Zou et al., 2020 | General population | 1 | 1 | 0 | 0 | 0 | 0 | 2 | D |
| Nopsopon et al., 2020 | Low-risk healthcare workers | 1 | 1 | 2 | 0 | 2 | 0 | 6 | C |

| | | | | | | | | | |
|---|---|---|---|---|---|---|---|---|---|
| McDade et al., 2020 | Poorly-defined population | 0 | 1 | 2 | 0 | 0 | 0 | 3 | D |
| Appa et al., 2020 | General population | 3 | 1 | 2 | 1 | 0 | 2 | 9 | B |
| Tönshoff et al. et al., 2020 | Poorly-defined population | 1 | 1 | 0 | 2 | 2 | 0 | 6 | C |
| Jerković et al., 2020 | Poorly-defined population | 1 | 1 | 0 | 0 | 2 | 0 | 4 | C |
| Alessandro et al., 2020 | General population; Low-risk healthcare workers | 2 | 1 | 0 | 0 | 2 | 0 | 5 | C |
| Halatoko et al., 2020 | Low-risk healthcare workers; Poorly-defined population | 2 | 1 | 2 | 0 | 0 | 0 | 5 | C |
| Dillner et al., 2020 | Poorly-defined population | 2 | 1 | 2 | 0 | 2 | 0 | 7 | B |
| Aziz et al., 2020 | General population | 3 | 1 | 0 | 2 | 0 | 0 | 6 | C |
| Chamie et al., 2020 | General population | 1 | 1 | 0 | 0 | 2 | 2 | 6 | C |
| Nesbitt et al., 2020 | Poorly-defined population | 1 | 1 | 0 | 1 | 2 | 0 | 5 | C |
| Wells et al., 2020 | General population | 1 | 1 | 2 | 1 | 0 | 0 | 5 | C |
| Fontanet et al., 2020 | Poorly-defined population | 1 | 1 | 0 | 0 | 2 | 0 | 4 | C |

| | | | | | | | | | |
|---|---|---|---|---|---|---|---|---|---|
| Sandri et al., 2020 | Poorly-defined population | 1 | 1 | 0 | 0 | 2 | 0 | 4 | C |
| Uyoga et al., 2020 | Poorly-defined population | 1 | 1 | 2 | 0 | 2 | 2 | 8 | B |
| Brant et al., 2020 | Low-risk healthcare workers | 1 | 1 | 0 | 0 | 2 | 2 | 6 | C |
| Addetia et al., 2020 | Poorly-defined population | 1 | 1 | 0 | 2 | 0 | 0 | 4 | C |
| Barallat et al., 2020 | Low-risk healthcare workers | 1 | 1 | 0 | 1 | 2 | 0 | 5 | C |
| Tess et al., 2020 | General population | 2 | 1 | 2 | 0 | 2 | 0 | 7 | B |
| Takita et al., 2020 | Poorly-defined population | 1 | 0 | 2 | 0 | 2 | 0 | 5 | C |
| Mattern et al., 2020 | Poorly-defined population | 1 | 1 | 0 | 0 | 0 | 0 | 2 | D |
| Carrat et al., 2020 | General population | 2 | 1 | 0 | 2 | 2 | 0 | 7 | B |
| McBride et al., 2020 | Poorly-defined population | 1 | 1 | 0 | 0 | 0 | 0 | 2 | D |
| Ebinger et al., 2020 | Poorly-defined population | 2 | 1 | 0 | 0 | 2 | 0 | 5 | C |
| Hurk et al., 2020 | Poorly-defined population | 1 | 1 | 2 | 0 | 2 | 0 | 6 | C |
| Weis et al., 2020 | General population | 3 | 1 | 0 | 1 | 0 | 0 | 5 | C |

| | | | | | | | | | |
|---|---|---|---|---|---|---|---|---|---|
| Rigatti et al., 2020 | Poorly-defined population | 1 | 1 | 0 | 0 | 2 | 0 | 4 | C |
| Gomes et al., 2020 | General population | 3 | 1 | 0 | 0 | 2 | 0 | 6 | C |
| Hallal et al., 2020 | General population | 3 | 1 | 2 | 0 | 2 | 2 | 10 | A |
| Nakamura et al., 2020 | Low-risk healthcare workers | 1 | 1 | 0 | 1 | 0 | 0 | 3 | D |
| Jespersen et al., 2020 | Poorly-defined population | 2 | 1 | 0 | 0 | 2 | 2 | 7 | B |
| Tsertsvadze et al., 2020 | General population | 1 | 1 | 0 | 0 | 2 | 2 | 6 | C |
| Chibwana et al., 2020 | Poorly-defined population | 1 | 1 | 0 | 0 | 0 | 2 | 4 | C |
| Armann et al., 2020 | Close contacts; General population | 2 | 1 | 0 | 1 | 0 | 0 | 4 | C |
| Hibino et al., 2020 | Poorly-defined population | 1 | 1 | 0 | 0 | 0 | 0 | 2 | D |
| Wilkins et al., 2020 | Low-risk healthcare workers | 2 | 1 | 0 | 0 | 2 | 0 | 5 | C |
| Alkurt et al., 2020 | Poorly-defined population | 2 | 1 | 0 | 0 | 0 | 0 | 3 | D |
| Vassallo et al., 2020 | Poorly-defined population | 1 | 1 | 0 | 0 | 2 | 0 | 4 | C |
| Melo et al., 2020 | Poorly-defined population | 2 | 1 | 2 | 0 | 0 | 0 | 5 | C |

| | | | | | | | | | |
|---|---|---|---|---|---|---|---|---|---|
| Favara et al., 2020 | High-risk healthcare workers | 2 | 1 | 2 | 1 | 0 | 0 | 6 | C |
| Silva et al., 2020 | Low-risk healthcare workers | 2 | 1 | 2 | 0 | 0 | 0 | 5 | C |
| Ray et al., 2020 | Poorly-defined population | 1 | 1 | 2 | 1 | 2 | 0 | 7 | B |
| Bardai et al., 2020 | Low-risk healthcare workers; Poorly-defined population | 1 | 1 | 0 | 0 | 0 | 0 | 2 | D |
| Mahajan et al., 2020 | General population | 2 | 1 | 2 | 1 | 2 | 0 | 8 | B |
| Nishida et al., 2020 | Low-risk healthcare workers | 1 | 1 | 0 | 0 | 2 | 0 | 4 | C |
| Nawa et al., 2020 | General population | 2 | 1 | 0 | 0 | 2 | 0 | 5 | C |
| Qutob et al., 2020 | General population; Poorly-defined population | 3 | 1 | 0 | 0 | 2 | 0 | 6 | C |
| Ulyte et al., 2020 | Poorly-defined population | 2 | 1 | 2 | 0 | 2 | 2 | 9 | B |
| Asuquo et al., 2020 | Poorly-defined population | 2 | 1 | 0 | 0 | 0 | 0 | 3 | D |
| Ward et al., 2020 | General population | 3 | 1 | 0 | 0 | 2 | 2 | 8 | B |
| Menezes et al., 2020 | General population | 3 | 1 | 2 | 0 | 0 | 0 | 6 | C |

| | | | | | | | | | |
|---|---|---|---|---|---|---|---|---|---|
| Ariza et al., 2020 | Low-risk healthcare workers | 1 | 1 | 2 | 1 | 2 | 0 | 7 | B |
| Majiya et al., 2020 | General population | 3 | 1 | 2 | 0 | 2 | 0 | 8 | B |
| Javed et al., 2020 | General population | 3 | 1 | 0 | 0 | 0 | 0 | 4 | C |
| Buonsenso et al., 2020 | Close contacts | 1 | 1 | 0 | 0 | 0 | 0 | 2 | D |
| Khan et al., 2020 | Poorly-defined population | 1 | 1 | 0 | 0 | 2 | 0 | 4 | C |
| Satpati et al., 2020 | General population | 3 | 1 | 0 | 0 | 2 | 0 | 6 | C |
| Silva et al., 2020 | General population | 3 | 1 | 0 | 0 | 2 | 0 | 6 | C |
| Calife et al., 2020 | General population | 3 | 1 | 0 | 0 | 0 | 0 | 4 | C |
| Kumar et al., 2020 | Low-risk healthcare workers | 1 | 1 | 0 | 1 | 2 | 0 | 5 | C |
| Official reports | | | | | | | | | |
| Public Health Ontario, Canada, 2020 | Poorly-defined population | 1 | 1 | 2 | 0 | 2 | 2 | 8 | B |
| Office of National Statistics, UK, 2020 | General population | 2 | 1 | 2 | 0 | 2 | 0 | 7 | B |
| the Government of Jersey, UK, 2020 | General population | 3 | 1 | 0 | 0 | 2 | 2 | 8 | B |

| | | | | | | | | | |
|---|---|---|---|---|---|---|---|---|---|
| Canadian Blood Services, 2020 | Poorly-defined population | 1 | 1 | 0 | 0 | 2 | 2 | 6 | C |
| Ministry of Health, Labour and Welfare, Japan, 2020 | General population | 2 | 1 | 0 | 1 | 0 | 0 | 4 | C |
| Islamic Republic of Afghanistan Ministry of Public Health, Afghanistan, 2020 | General population | 3 | 1 | 0 | 0 | 0 | 0 | 4 | C |
| Public Health England, 2020 | Poorly-defined population | 1 | 1 | 0 | 0 | 2 | 2 | 6 | C |
| MedLife, Romania, 2020 | Poorly-defined population | 1 | 1 | 0 | 0 | 0 | 0 | 2 | D |

**Appendix Table 8. The summary of fifty-one grade A and grade B studies included into the main analysis on the basis of WHO regions and pre-defined study populations**

| Author, Country | Study population | No. of specimens tested | Grade |
|---|---|---|---|
| **African Region** | | | |
| Majiya et al., Nigeria | General population | 185 | B |
| Uyoga et al., Kenya | Poorly-defined population | 3098 | B |
| **Region of the Americas** | | | |
| Self et al., USA | Low-risk Healthcare worker | 3248 | B |
| Ariza et al., Colombia | Low-risk Healthcare worker | 351 | B |
| Moscola et al., USA | Low-risk Healthcare worker | 40329 | B |
| Sood et al., USA | General population | 863 | A |
| Hallal et al., Brazil | General population | 56123 | A |
| Naranbhai et al., USA | General population | 200 | B |
| Mahajan et al., USA | General population | 567 | B |
| Tess et al., Brazil | General population | 517 | B |
| Rosenberg et al., USA | General population | 15101 | B |
| Bendavid et al., USA | General population | 3330 | B |
| Biggs et al., USA | General population | 696 | B |
| Appa et al., USA | General population | 1880 | B |
| Public Health Ontario, Canada | Poorly-defined population | 8902 | B |
| Buss et al., Brazil | Poorly-defined population | 13867 | B |
| Skowronski et al., Canada | Poorly-defined population | 1754 | B |
| Havers et al., USA | Poorly-defined population | 16025 | B |
| **Eastern Mediterranean Region** | | | |
| Shakiba et al., Iran | General population | 528 | A |
| Nisar et al., Pakistan | General population | 2004 | B |
| **European Region** | | | |
| Iversen et al., Denmark | Low-risk Healthcare worker | 28792 | A |
| Basteiro et al., Spain | Low-risk Healthcare worker | 578 | B |
| Stringhini et al., Switzerland | General population | 2766 | A |
| Office of National Statistics, UK | General population | 9343 | B |
| Gudbjartsson et al., Iceland | General population | 5506 | B |
| Carrat et al., France | General population | 14628 | B |
| Streeck et al., Germany | General population | 919 | B |
| Snoeck et al., Luxembourg | General population | 1820 | B |
| Marina et al., Spain | General population | 51958 | B |
| Petersen et al., Denmark | General population | 1075 | B |
| Government of Jersey, UK | General population | 855 | B |
| Ward et al., UK | General population | 99908 | B |

| | | | |
|---|---|---|---|
| Iversen et al., Denmark | Poorly-defined population | 4672 | A |
| Gudbjartsson et al., Iceland | Poorly-defined population | 22831 | B |
| Vena et al., Italy | Poorly-defined population | 3609 | B |
| Ulyte et al., Switzerland | Poorly-defined population | 2484 | B |
| Dillner et al., Sweden | Poorly-defined population | 12928 | B |
| Valenti et al., Italy | Poorly-defined population | 789 | B |
| Erikstrup et al., Denmark | Poorly-defined population | 20640 | B |
| Bogogiannidou et al., Greece | Poorly-defined population | 6586 | B |
| Jespersen et al., Denmark | Poorly-defined population | 17948 | B |
| Plebani et al., Italy | Poorly-defined population | 8285 | B |
| **South-East Asia Region** | | | |
| Nopsopon et al., Thailand | Low-risk Healthcare worker | 675 | B |
| Ray et al., India | Poorly-defined population | 212 | B |
| Nopsopon et al., Thailand | Poorly-defined population | 182 | B |
| **Western Pacific Region** | | | |
| Chen et al., China | High-risk Healthcare worker | 105 | B |
| Wang et al., China | General population | 2184 | B |
| Ling et al., China | General population | 18721 | B |
| To et al., China | Poorly-defined population | 1265 | B |
| Sam et al., Malaysia | Poorly-defined population | 588 | B |
| Xu et al., China | Poorly-defined population | 4747 | B |

#### Appendix Table 9. Estimated seroprevalence of antibodies to SARS-CoV-2 by WHO regions and study population among fifty-one grade A and grade B studies

| | All infections | | | | | Symptomatic infections | | | | | Asymptomatic infections | | | | |
|---|---|---|---|---|---|---|---|---|---|---|---|---|---|---|---|
| Study population | No. of studies | Total no. of positive | Total no. of participants provided serum | Estimated seroprevalence (95% confidence interval) | $I^2$ (P) | No. of studies | Total no. of positive | Total no. of participants provided serum | Estimated seroprevalence (95% confidence interval) | $I^2$ (P) | No. of studies | Total no. of positive | Total no. of participants provided serum | Estimated seroprevalence (95% confidence interval) | $I^2$ (P) |
| **Overall** | | | | | | | | | | | | | | | |
| Close contacts | 0 | - | - | - | - | 0 | - | - | - | - | 0 | - | - | - | - |
| High-risk HCWs | 1 | 18 | 105 | 17.1 [9.9-24.4] | - | 1 | 4 | 105 | 3.8 [0.1-7.5] | - | 1 | 14 | 105 | 13.3 [6.8-19.8] | - |
| Low-risk HCWs | 6 | 6538 | 73973 | 5.4 [0.7-10.1] | 99.9 (p<0.001) | 3 | 143 | 4274 | 1.8 [0.0-4.7] | 98.3 (p<0.001) | 3 | 60 | 4274 | 0.9 [0.0-2.3] | 95.9 (p<0.001) |
| General population | 24 | 13380 | 291607 | 5.3 [4.2-6.4] | 99.8 (p<0.001) | 11 | 5109 | 178832 | 1.9 [1.1-2.8] | 99.8 (p<0.001) | 11 | 3661 | 178832 | 1.7 [0.9-2.5] | 99.5 (p<0.001) |
| Poorly-defined population | 20 | 6476 | 151412 | 4.1 [3.0-5.1] | 99.6 (p<0.001) | 2 | 0 | 971 | 0.0 [0.0-0.2] | 0.0 (p=1.000) | 2 | 41 | 971 | 2.8 [0.0-7.2] | 95.5 (p<0.001) |
| **African Region** | | | | | | | | | | | | | | | |
| Close contacts | 0 | - | - | - | - | 0 | - | - | - | - | 0 | - | - | - | - |
| High-risk HCWs | 0 | - | - | - | - | 0 | - | - | - | - | 0 | - | - | - | - |
| Low-risk HCWs | 0 | - | - | - | - | 0 | - | - | - | - | 0 | - | - | - | - |
| General population | 1 | 47 | 185 | 25.4 [19.1-31.7] | - | 1 | 25 | 185 | 13.5 [8.6-18.4] | - | 1 | 22 | 185 | 11.9 [7.2-16.6] | - |
| Poorly-defined population | 1 | 174 | 3098 | 5.6 [4.8-6.4] | - | 0 | - | - | - | - | 0 | - | - | - | - |
| **Region of the Americas** | | | | | | | | | | | | | | | |
| Close contacts | 0 | - | - | - | - | 0 | - | - | - | - | 0 | - | - | - | - |

| | | | | | | | | | | | | | | | |
|---|---|---|---|---|---|---|---|---|---|---|---|---|---|---|---|
| High-risk HCWs | 0 | - | - | - | - | 0 | - | - | - | - | 0 | - | - | - | - |
| Low-risk HCWs | 3 | 5725 | 43928 | 7.3 [0.5-14.2] | 99.6 (p<0.001) | 2 | 142 | 3599 | 2.7 [0.0-5.8] | 95.4 (p<0.001) | 2 | 60 | 3599 | 1.6 [1.2-2.1] | 0.0 (p=0.339) |
| General population | 9 | 3182 | 79207 | 5.2 [2.9-7.5] | 99.5 (p<0.001) | 3 | 56 | 2126 | 2.5 [1.7-3.3] | 29.6 (p=0.242) | 3 | 21 | 2126 | 1.0 [0.6-1.4] | 0.0 (p=0.813) |
| Poorly-defined population | 4 | 2483 | 40548 | 4.5 [1.0-8.0] | 99.8 (p<0.001) | 0 | - | - | - | - | 0 | - | - | - | - |
| **Eastern Mediterranean Region** | | | | | | | | | | | | | | | |
| Close contacts | 0 | - | - | - | - | 0 | - | - | - | - - | 0 | - | - | - | - |
| High-risk HCWs | 0 | - | - | - | - | 0 | - | - | - | - - | 0 | - | - | - | - |
| Low-risk HCWs | 0 | - | - | - | - | 0 | - | - | - | - - | 0 | - | - | - | - |
| General population | 2 | 276 | 2532 | 14.4 [2.2-26.7] | 97.8 (p<0.001) | 0 | - | - | - | - - | 0 | - | - | - | - |
| Poorly-defined population | 0 | - | - | - | - | 0 | - | - | - | - - | 0 | - | - | - | - |
| **European Region** | | | | | | | | | | | | | | | |
| Close contacts | 0 | - | - | - | - | 0 | - | - | - | - | 0 | - | - | - | - |
| High-risk HCWs | 0 | - | - | - | - | 0 | - | - | - | - | 0 | - | - | - | - |
| Low-risk HCWs | 2 | 812 | 29370 | 5.0 [0.2-9.9] | 95.0 (p<0.001) | 0 | - | - | - | - | 0 | - | - | - | - |
| General population | 10 | 9248 | 188778 | 4.2 [2.7-5.8] | 99.6 (p<0.001) | 5 | 5028 | 155616 | 1.9 [0.6-3.1] | 99.5 (p<0.001) | 5 | 2991 | 155616 | 1.5 [1.0-2.1] | 97.6 (p<0.001) |
| Poorly-defined population | 10 | 3668 | 100772 | 4.3 [2.9-5.6] | 99.7 (p<0.001) | 1 | 0 | 789 | 0.0 [0.0-0.2] | - | 1 | 40 | 789 | 5.1 [3.5-6.6] | - |
| **South-East Asia Region** | | | | | | | | | | | | | | | |
| Close contacts | 0 | - | - | - | - | 0 | - | - | - | - | 0 | - | - | - | - |
| High-risk HCWs | 0 | - | - | - | - | 0 | - | - | - | - | 0 | - | - | - | - |
| Low-risk HCWs | 1 | 1 | 675 | 0.1 [0.0-0.4] | - | 1 | 1 | 675 | 0.1 [0.0-0.4] | - | 1 | 0 | 675 | 0.0 [0.0-0.2] | - |
| General population | 0 | - | - | - | - | 0 | - | - | - | - | 0 | - | - | - | - |
| Poorly-defined population | 2 | 43 | 394 | 10.0 [0.0-28.9] | 99.6 (p<0.001) | 1 | 0 | 182 | 0.0 [0.0-0.8] | - | 1 | 1 | 182 | 0.5 [0.0-1.6] | - |

| | | | | | | | | | | | | | | | |
|---|---|---|---|---|---|---|---|---|---|---|---|---|---|---|---|
| **Western Pacific Region** | | | | | | | | | | | | | | | |
| Close contacts | 0 | - | - | - | - | 0 | - | - | - | - | 0 | - | - | - | - |
| High-risk HCWs | 1 | 18 | 105 | 17.1 [9.9-24.4] | - | 1 | 4 | 105 | 3.8 [0.1-7.5] | - | 1 | 14 | 105 | 13.3 [6.8-19.8] | - |
| Low-risk HCWs | 0 | - | - | - | - | 0 | - | - | - | - | 0 | - | - | - | - |
| General population | 2 | 627 | 20905 | 1.7 [0.0-5.0] | 99.8 (p<0.001) | 2 | 0 | 20905 | 0.0 [0.0-0.0] | 0.0 (p=1.000) | 2 | 627 | 20905 | 1.7 [0.0-5.0] | 99.8 (p<0.001) |
| Poorly-defined population | 3 | 108 | 6600 | 1.2 [0.4-2.1] | 87.5 (p<0.001) | 0 | - | - | - | - | 0 | - | - | - | - |

#### Appendix Table 10. Sensitivity analysis of seroprevalence of antibodies to SARS-CoV-2 among fifty-one grade A and grade B studies, considering alternative serological assays used in the same study

| | Studies using original serological assays | | | | | Studies using alternative serological assays | | | | |
|---|---|---|---|---|---|---|---|---|---|---|
| **Study population** | **No. of studies** | **Total no. of positive** | **Total no. of participants provided serum** | **Estimated seroprevalence (95% confidence interval)** | **$I^2$ (P)** | **No. of studies** | **Total no. of positive** | **Total no. of participants provided serum** | **Estimated seroprevalence (95% confidence interval)** | **$I^2$ (P)** |
| **Overall** | | | | | | | | | | |
| Close contacts | 0 | - | - | - | - | 0 | - | - | - | - |
| High-risk HCWs | 1 | 18 | 105 | 17.1 [9.9-24.4] | - | 1 | 18 | 105 | 17.1 [9.9-24.4] | - |
| Low-risk HCWs | 6 | 6538 | 73973 | 5.4 [0.7-10.1] | 99.9 (p<0.001) | 6 | 6538 | 73973 | 5.4 [0.7-10.1] | 99.9 (p<0.001) |
| General population | 24 | 13380 | 291607 | 5.3 [4.2-6.4] | 99.8 (p<0.001) | 24 | 13828 | 296629 | 5.3 [4.2-6.4] | 99.8 (p<0.001) |
| Poorly-defined population | 20 | 6476 | 151412 | 4.1 [3.0-5.1] | 99.6 (p<0.001) | 20 | 6476 | 151412 | 4.1 [3.0-5.1] | 99.6 (p<0.001) |
| **African Region** | | | | | | | | | | |
| General population | 1 | 47 | 185 | 25.4 [19.1-31.7] | - | 1 | 47 | 185 | 25.4 [19.1-31.7] | - |
| Poorly-defined population | 1 | 174 | 3098 | 5.6 [4.8-6.4] | - | 1 | 174 | 3098 | 5.6 [4.8-6.4] | - |
| **Region of the Americas** | | | | | | | | | | |
| Low-risk HCWs | 3 | 5725 | 43928 | 7.3 [0.5-14.2] | 99.6 (p<0.001) | 3 | 5725 | 43928 | 7.3 [0.5-14.2] | 99.6 (p<0.001) |
| General population | 9 | 3182 | 79207 | 5.2 [2.9-7.5] | 99.5 (p<0.001) | 9 | 3177 | 79207 | 5.0 [3.0-7.1] | 99.6 (p<0.001) |
| Poorly-defined population | 4 | 2483 | 40548 | 4.5 [1.0-8.0] | 99.8 (p<0.001) | 4 | 2483 | 40548 | 4.5 [1.0-8.0] | 99.8 (p<0.001) |
| **Eastern Mediterranean Region** | | | | | | | | | | |
| General population | 2 | 276 | 2532 | 14.4 [2.2-26.7] | 97.8 (p<0.001) | 2 | 276 | 2532 | 14.4 [2.2-26.7] | 97.8 (p<0.001) - |
| **European Region** | | | | | | | | | | |
| Low-risk HCWs | 2 | 812 | 29370 | 5.0 [0.2-9.9] | 95.0 (p<0.001) | 2 | 812 | 29370 | 5.0 [0.2-9.9] | 95.0 (p<0.001) |
| General population | 10 | 9248 | 188778 | 4.2 [2.7-5.8] | 99.6 (p<0.001) | 10 | 9701 | 193800 | 4.3 [2.7-5.8] | 99.7 (p<0.001) |
| Poorly-defined population | 10 | 3668 | 100772 | 4.3 [2.9-5.6] | 99.7 (p<0.001) | 10 | 3668 | 100772 | 4.3 [2.9-5.6] | 99.7 (p<0.001) |
| **South-East Asia Region** | | | | | | | | | | |
| Low-risk HCWs | 1 | 1 | 675 | 0.1 [0.0-0.4] | - | 1 | 1 | 675 | 0.1 [0.0-0.4] | - |

| | | | | | | | | | | |
|---|---|---|---|---|---|---|---|---|---|---|
| Poorly-defined population | 2 | 43 | 394 | 10.0 [0.0-28.9] | 99.6 (p<0.001) | 2 | 43 | 394 | 10.0 [0.0-28.9] | 99.7 (p<0.001) |
| **Western Pacific Region** | | | | | | | | | | |
| High-risk HCWs | 1 | 18 | 105 | 17.1 [9.9-24.4] | - | 1 | 18 | 105 | 17.1 [9.9-24.4] | - |
| General population | 2 | 627 | 20905 | 1.7 [0.0-5.0] | 99.8 (p<0.001) | 2 | 627 | 20905 | 1.7 [0.0-5.0] | 99.8 (p<0.001) |
| Poorly-defined population | 3 | 108 | 6600 | 1.2 [0.4-2.1] | 87.5 (p<0.001) | 3 | 108 | 6600 | 1.2 [0.4-2.1] | 87.5 (p<0.001) |

**Appendix Table 11. Multivariable meta-regression for change in the seroprevalence of human antibodies to SARS-CoV-2 among fifty-one grade A and grade B studies**

| Study characteristics | Change in the seroprevalence (coefficient β†) (95% CI) |
|---|---|
| WHO regions | |
| African Region | 1 |
| Region of the Americas | -8.6 (-17.0, -0.2)* |
| Eastern Mediterranean Region | -0.2 (-11.5, 11.1) |
| European Region | -9.7 (-17.9, -1.5)* |
| South-East Asia Region | -8.0 (-18.2, 2.2) |
| Western Pacific Region | -12.6 (-21.8, -3.4)** |
| Study populations | |
| General population | 1 |
| High-risk healthcare worker | 15.3 (1.6, 28.9)* |
| Low-risk healthcare worker | -0.2 (-5.2, 4.8) |
| Poorly-defined group | -0.8 (-4.2, 2.5) |
| Study quality | |
| Grade A | 1 |
| Grade B | 0.1 (-4.7, 4.9) |

*** $p<0.001$; **$0.001<p<0.01$; *$0.01<p<0.05$.
† The regression coefficient β refers to the change in the seroprevalence of human antibodies to SARS-CoV-2. A negative sign for the coefficient β corresponds to a reduction in the seroprevalence of SARS-CoV-2 specific antibodies for given changes in the covariate, while a positive sign corresponds to an increase in the seroprevalence of SARS-CoV-2 specific antibodies.

**Appendix Table 12. Relative risk of infections with SARS-CoV-2 by age groups and sex among grade A and grade B studies**

| Categories | Relative risk (RR, 95% CI) |
|---|---|
| **Overall** | |
| Age group† | |
| Young | 0.764 (0.691-0.845)* |
| Middle-age | Ref |
| Old | 0.708 (0.536-0.937) * |
| Sex | |
| Female | Ref |
| Male | 1.033 (0.946-1.127) |
| **Region of the Americas** | |
| Age group† | |
| Young | 0.714 (0.599-0.850)* |
| Middle-age | Ref |
| Old | 0.736 (0.604-0.897)* |
| Sex | |
| Female | Ref |
| Male | 1.103 (0.936-1.301) |
| **European Region** | |
| Age group† | |
| Young | 0.790 (0.699-0.893)* |
| Middle-age | Ref |
| Old | 0.713 (0.495-1.027) |
| Sex | |
| Female | Ref |
| Male | 1.006 (0.940-1.077) |

* $p<0.05$

† The age groups between each study were not perfectly aligned. Specially, the Young represent participants younger than 20 years, while the old represent participants older than 65 years. The Middle-age group represent participants aged 20 – 64 years.

**Appendix Table 13. The cumulative incidence and estimated number of serological infections of selected grade A an**

| Author | Location, Country | Age of participants | Unadjusted seroprevalence (%) (a) | Adjusted seroprevalence (%) (factors) (a') * | Total population (b) | Age proportion for population (%) (c) |
|---|---|---|---|---|---|---|
| **Region of the Americas** | | | | | | |
| Hallal et al. ‖ | Brazil | ≥ 1 yrs | 1.95 | - | 213863051 | 98.7 |
| Biggs et al. ‡ | DeKalb, Fulton County, Georgia, USA | All ages | 2.7 | 2.5 | 1806672 | 100 |
| Appa et al. † | Marin, California, USA | ≥ 4 yrs | 0.5 | 0.29 | 258826 | 95.5 |
| Mahajan et al. † | Connecticut, USA | ≥ 18 yrs | 4.1 | 4.0 | 3565287 | 76.7 |
| **European Region** | | | | | | |
| Petersen et al. ‡ | Faroe Islands, Denmark | All ages | 0.6 | 0.7 | 52154 | 100 |
| Marina et al. ¶ | Spain | All ages | 4.6 | - | 46459218 | 100 |
| Stringhini et al. ‖ | Geneva, Switzerland | ≥ 5 yrs | 7.9 | - | 504128 | 94.8 |
| Ward et al. ‡ | England, UK | ≥ 18 yrs | 5.5 | 6.0 | 56286961 | 78.6 |
| Office of National Statistics.‡ | England, UK | ≥ 16 yrs | 5.1 | 6.2 | - | - |
| Government of Jersey ‡ | Jersey, UK | ≥ 16 yrs | 2.9 | 3.1 | - | - |
| Streeck et al. ‡§ | Gangelt, Kreis Heinsberg, Germany | All ages | 11.5 | 14.1 | 12597 | 100 |
| Snoeck et al. † | Luxembourg | ≥ 18 yrs | 1.9 | 2.09 | 603951 | - |

* Adjust factors mainly include demographic factors (age and/or sex) and test performance (sensitivity and specificity of assays).

‖ We aggregated the multiple sampling results to calculate the crude estimated during the whole study period, and adjusted seroprevalence could not be calculated.

‡ The estimated number of infections or population size were reported in their own study.

¶ The seroprevalence of immunoassay were used in main analysis

†The age proportion used to calculate population size in specific age groups were not perfectly aligned with the age group reported in original study.

§ The number of COVID-19 cases in the local as of Mar 30 were extracted in the original study to represent the cumulative number of cases as of Mar 20.

#### Appendix Table 14. The data source of population size and COVID-19-related epidemiological data of grade A and gr

| Author | Location | Institution | Source of population | Institution |
|---|---|---|---|---|
| Biggs et al. | Georgia, USA | National Center for Health Statistics | https://www.cdc.gov/nchs/nvss/bridged_race/data_documentation. htm#Vintage2018. | JHU CSSE COVID-19 Dashboard |
| Appa et al. | Marin, California, USA | U.S. Census Bureau, Population Division | https://archive.vn/20200214061229/https://factfinder.census.gov/faces/tableservices/jsf/pages/productview.xhtml?pid=PEP_2018_PEPANNRES&prodType=table https://www.census.gov/quickfacts/marincountycalifornia | JHU CSSE COVID-19 Dashboard |
| Mahajan et al. | Connecticut, USA | United State Census Bureau/ Statista | https://www2.census.gov/programs-surveys/popest/tables/2010-2019/state/totals/nst-est2019-01.xlsx?# https://www.statista.com/statistics/1021891/connecticut-population-share-age-group/ | JHU CSSE COVID-19 Dashboard/ Connecticut Open Data |
| Petersen et al. | Faroe Islands, Denmark | STATBANK | https://statbank.hagstova.fo/pxweb/en/H2/H2__IB__IB01/fo_aldbyg d.px/ | JHU CSSE COVID-19 Dashboard |
| Carrat et al. | France | World pop | https://www.worldpop.org/ | JHU CSSE COVID-19 Dashboard/ Ministère des Solidarités et de la Santé |
| Gudbjartsson et al. | Iceland | World pop | https://www.worldpop.org/ | JHU CSSE COVID-19 Dashboard |
| Marina et al. | Spain | World pop | https://www.worldpop.org/ | JHU CSSE COVID-19 Dashboard |
| Stringhini et al. | Geneva, Switzerland | STAT-TAB | https://www.pxweb.bfs.admin.ch/pxweb/en/px-x-0103010000_101/-/px-x-0103010000_101.px/?rxid=34873e36-d320-4c20-b931-8f0596e0e667 | Federal Office of Public Health |
| Ward et al. | England, UK | Office for National Statistics | https://www.ons.gov.uk/peoplepopulationandcommunity/populationandmigration/populationestimates | Public Health England |

| | | | | |
|---|---|---|---|---|
| Office of National Statistics. | England, UK | Office for National Statistics | https://www.ons.gov.uk/peoplepopulationandcommunity/populationandmigration/populationestimates | Public Health England |
| Government of Jersey | Jersey, UK | - | - | Government of Jersey |
| Streeck et al. | Gangelt, Kreis Heinsberg, Germany | - | Streeck et al. Infection fatality rate of SARS-CoV-2 infection in a German community with a super-spreading event. medRxiv. https://doi.org/10.1101/2020.05.11.20092916 | - |
| Snoeck et al. | Luxembourg | World pop | https://www.worldpop.org/ | JHU CSSE COVID-19 Dashboard/The Luxembourg government |

#### Appendix Table 15. Estimated seroprevalence of antibodies to SARS-CoV-2 by WHO regions and study population an

| Study population | All infections | | | | | Symptomatic infections | | | | |
|---|---|---|---|---|---|---|---|---|---|---|
| | No. of studies | Total no. of positive | Total no. of participants provided serum | Estimated seroprevalence (95% confidence interval) | I² (P) | No. of studies | Total no. of positive | Total no. of participants provided serum | Estimated seroprevalence (95% confidence interval) | I² (P) |
| **Overall** | | | | | | | | | | |
| Close contacts | 16 | 2901 | 9349 | 22.9 [11.1-34.7] | 99.6 (p<0.001) | 8 | 218 | 2082 | 7.6 [2.3-12.9] | 96.8 (p<0.001) |
| High-risk HCWs | 9 | 525 | 5369 | 14.8 [6.2-23.4] | 98.0 (p<0.001) | 5 | 56 | 831 | 5.5 [3.0-8.1] | 55.2 (p=0.063) |
| Low-risk HCWs | 69 | 12488 | 178176 | 5.6 [4.7-6.4] | 99.4 (p<0.001) | 38 | 1430 | 59926 | 0.5 [0.4-0.7] | 97.7 (p<0.001) |
| General population | 55 | 24525 | 468399 | 6.5 [5.8-7.2] | 99.8 (p<0.001) | 24 | 6951 | 255521 | 1.3 [1.2-1.5] | 99.7 (p<0.001) |
| Poorly-defined population | 111 | 25230 | 779004 | 5.3 [4.9-5.7] | 99.3 (p<0.001) | 43 | 2278 | 150518 | 0.1 [0.1-0.2] | 98.5 (p<0.001) |
| **African Region** | | | | | | | | | | |
| Close contacts | 0 | - | - | - | - | 0 | - | - | - | - |
| High-risk HCWs | 0 | - | - | - | - | 0 | - | - | - | - |
| Low-risk HCWs | 1 | 5 | 370 | 1.4 [0.2-2.5] | 99.6 (p<0.001) | 0 | - | - | - | - |
| General population | 2 | 50 | 284 | 14.1 [0.0-36.0] | 98.0 (p<0.001) | 2 | 26 | 284 | 7.0 [0.0-19.3] | 95.3 (p<0.001) |
| Poorly-defined population | 4 | 278 | 4249 | 9.8 [4.6-14.9] | 99.4 (p<0.001) | 1 | 0 | 500 | 0.0 [0.0-0.3] | - |
| **Region of the Americas** | | | | | | | | | | |
| Close contacts | 7 | 381 | 4092 | 10.1 [3.5-16.8] | 97.0 (p<0.001) | 5 | 198 | 1700 | 8.1 [0.0-17.2] | 95.9 (p<0.001) |
| High-risk HCWs | 0 | - | - | - | - | 0 | - | - | - | - |
| Low-risk HCWs | 19 | 6626 | 69768 | 6.4 [3.8-9.0] | 99.5 (p<0.001) | 10 | 170 | 15925 | 1.1 [0.6-1.7] | 94.9 (p<0.001) |
| General population | 19 | 6116 | 137746 | 7.3 [5.9-8.7] | 99.5 (p<0.001) | 6 | 1727 | 41759 | 6.4 [3.2-9.5] | 99.5 (p<0.001) |
| Poorly-defined population | 29 | 15263 | 519830 | 5.9 [5.2-6.6] | 99.6 (p<0.001) | 9 | 271 | 55839 | 0.7 [0.4-1.1] | 97.5 (p<0.001) |
| **Eastern Mediterranean Region** | | | | | | | | | | |
| Close contacts | 0 | - | - | - | - | 0 | - | - | - | - |
| High-risk HCWs | 0 | - | - | - | - | 0 | - | - | - | - |
| Low-risk HCWs | 2 | 3 | 151 | 1.5 [0.0-5.3] | 63.0 (p=0.100) | 0 | - | - | - | - |
| General population | 6 | 5822 | 37717 | 12.6 [4.0-21.1] | 99.9 (p<0.001) | 1 | 1 | 142 | 0.7 [0.0-2.1] | - |
| Poorly-defined population | 3 | 85 | 2252 | 3.1 [1.2-5.0] | 98.1 (p<0.001) | 1 | 0 | 746 | 0.0 [0.0-0.2] | - |
| **European Region** | | | | | | | | | | |

| | | | | | | | | | | |
|---|---|---|---|---|---|---|---|---|---|---|
| Close contacts | 8 | 2503 | 5137 | 34.4 [19.0-49.7] | 98.6 (p<0.001) | 2 | 11 | 262 | 6.3 [0.0-19.5] | 92.1 (p<0.001) |
| High-risk HCWs | 7 | 354 | 1432 | 16.1 [5.1-27.2] | 97.2 (p<0.001) | 4 | 52 | 726 | 6.0 [2.9-9.1] | 60.3 (p=0.056) |
| Low-risk HCWs | 33 | 5562 | 93244 | 7.2 [5.9-8.5] | 99.1 (p<0.001) | 18 | 1246 | 35521 | 1.4 [1.2-1.7] | 98.8 (p<0.001) |
| General population | 20 | 10141 | 210967 | 6.1 [4.9-7.3] | 99.7 (p<0.001) | 10 | 5197 | 168598 | 2.6 [1.5-3.7] | 99.6 (p<0.001) |
| Poorly-defined population | 53 | 8087 | 168119 | 6.1 [5.4-6.9] | 99.2 (p<0.001) | 23 | 1951 | 30153 | 3.6 [3.1-4.1] | 99.1 (p<0.001) |
| **South-East Asia Region** | | | | | | | | | | |
| Close contacts | 0 | - | - | - | - | 0 | - | - | - | - |
| High-risk HCWs | 0 | - | - | - | - | 0 | - | - | - | - |
| Low-risk HCWs | 3 | 75 | 1967 | 2.2 [0.7-3.6] | 97.7 (p<0.001) | 3 | 9 | 1967 | 0.2 [0.0-0.7] | 76.4 (p=0.014) |
| General population | 1 | 19 | 458 | 4.1 [2.3-6.0] | - | 1 | 0 | 458 | 0.0 [0.0-0.3] | - |
| Poorly-defined population | 4 | 232 | 3632 | 10.9 [5.6-16.2] | 97.8 (p<0.001) | 3 | 56 | 3420 | 2.8 [0.5-5.1] | 95.7 (p<0.001) |
| **Western Pacific Region** | | | | | | | | | | |
| Close contacts | 1 | 17 | 120 | 14.2 [7.9-20.4] | - | 1 | 9 | 120 | 7.5 [2.8-12.2] | - |
| High-risk HCWs | 2 | 171 | 3937 | 10.1 [0.0-22.9] | 92.1 (p<0.001) | 1 | 4 | 105 | 3.8 [0.1-7.5] | - |
| Low-risk HCWs | 11 | 217 | 12676 | 1.2 [0.6-1.8] | 94.8 (p<0.001) | 7 | 5 | 6513 | 0.0 [0.0-0.0] | 0.0 (p=0.547) |
| General population | 7 | 2377 | 81227 | 1.8 [0.9-2.6] | 99.7 (p<0.001) | 4 | 0 | 44280 | 0.0 [0.0-0.0] | 0.0 (p=1.000) |
| Poorly-defined population | 18 | 1285 | 80922 | 2.0 [1.6-2.5] | 98.0 (p<0.001) | 6 | 0 | 59860 | 0.0 [0.0-0.0] | 0.0 (p=1.000) |

#### Appendix figures

**Appendix Figure 1. Quality scores assigned to SARS-CoV-2 serological studies by study populations, December 2019-September 2020.**

**(A)** Median quality score and range from assessment of serological studies of close contacts, high-risk healthcare workers, low-risk healthcare workers, general population, and Poorly-defined population. **(B)** Quality of studies by grade category (i.e. A, B, C and D). Category A included studies with scores ranging from 10 to 12, category B from 7 to 9, category C from 4 to 6, and category D from 0 to 3.

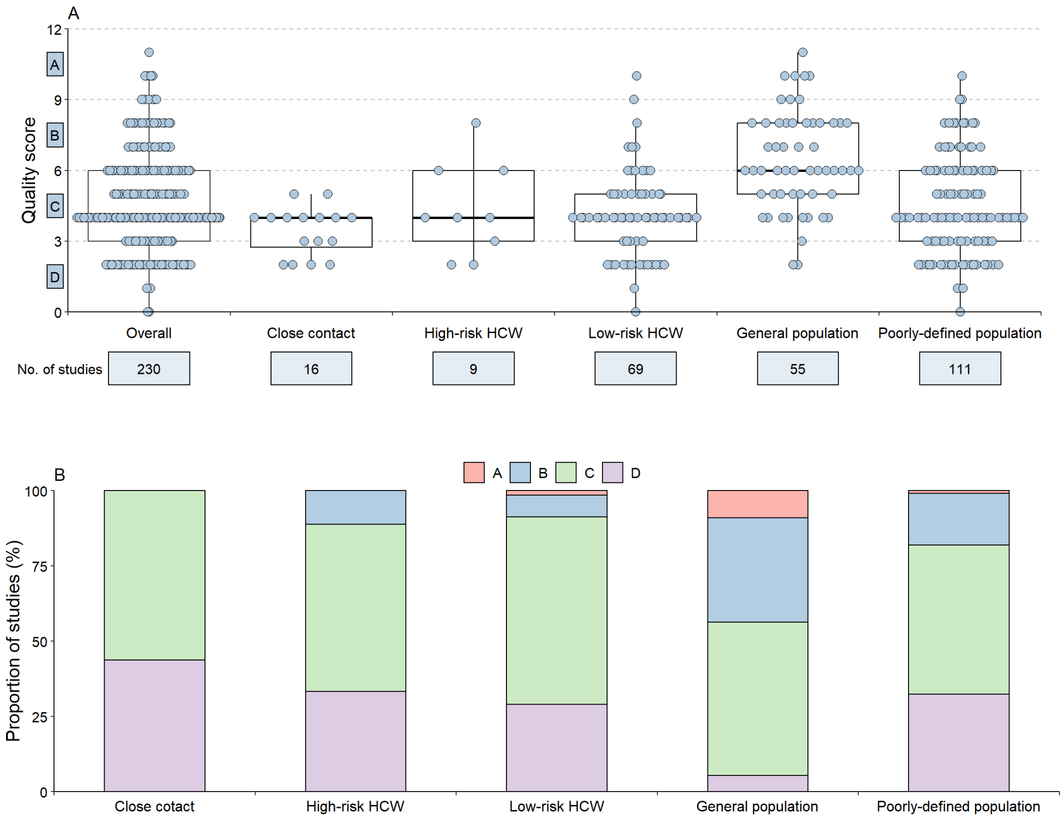

#### Appendix Figure 2. The starting sampling date for each serological study included in this meta-analysis in African Region

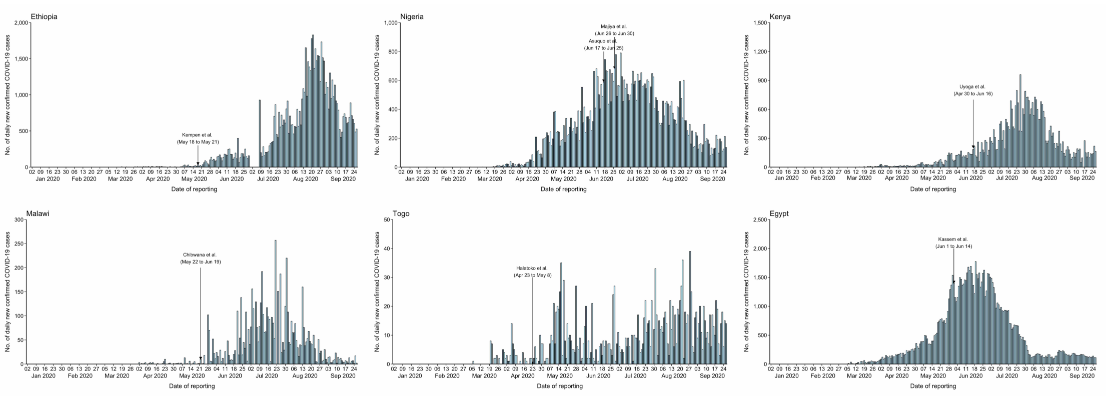

#### Appendix Figure 3. The starting sampling date for each serological study included in this meta-analysis in region of the Americas

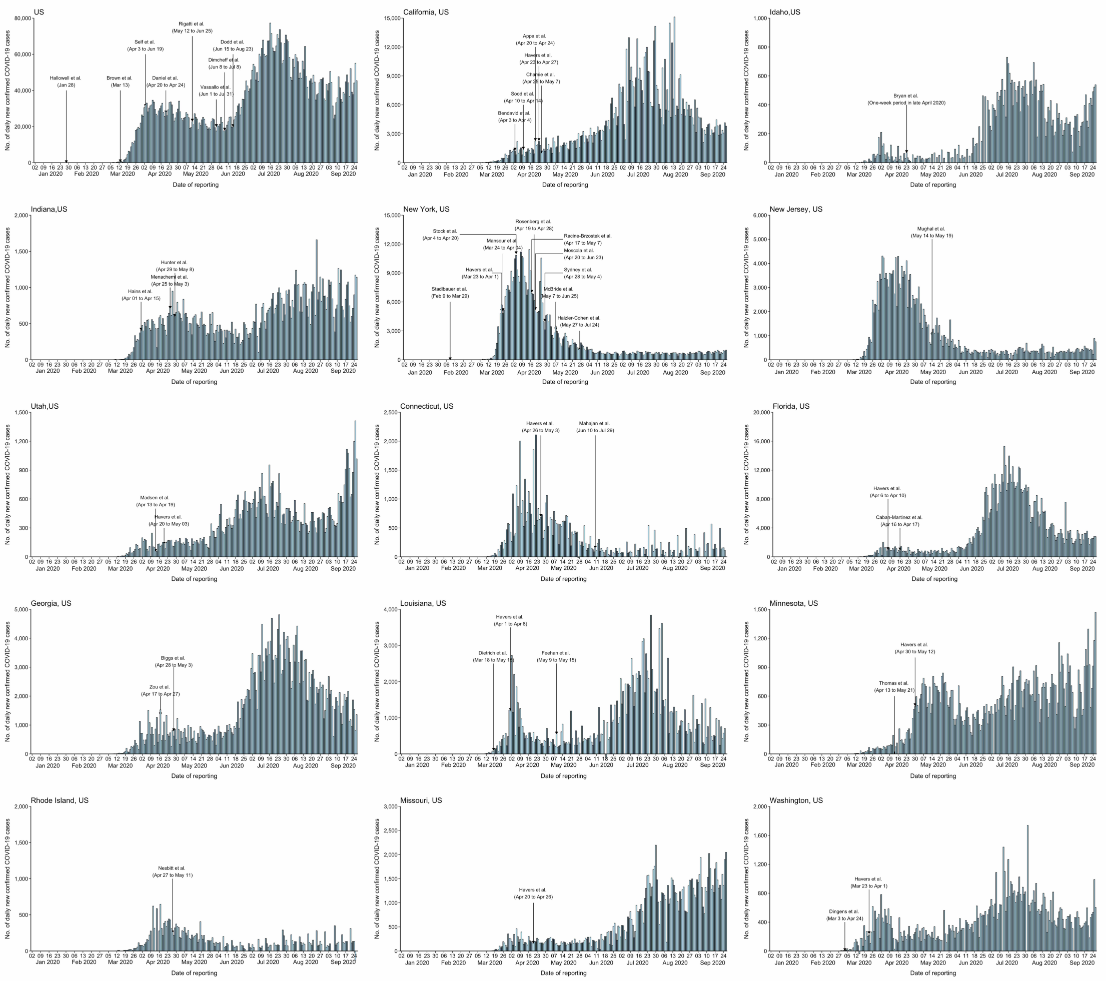

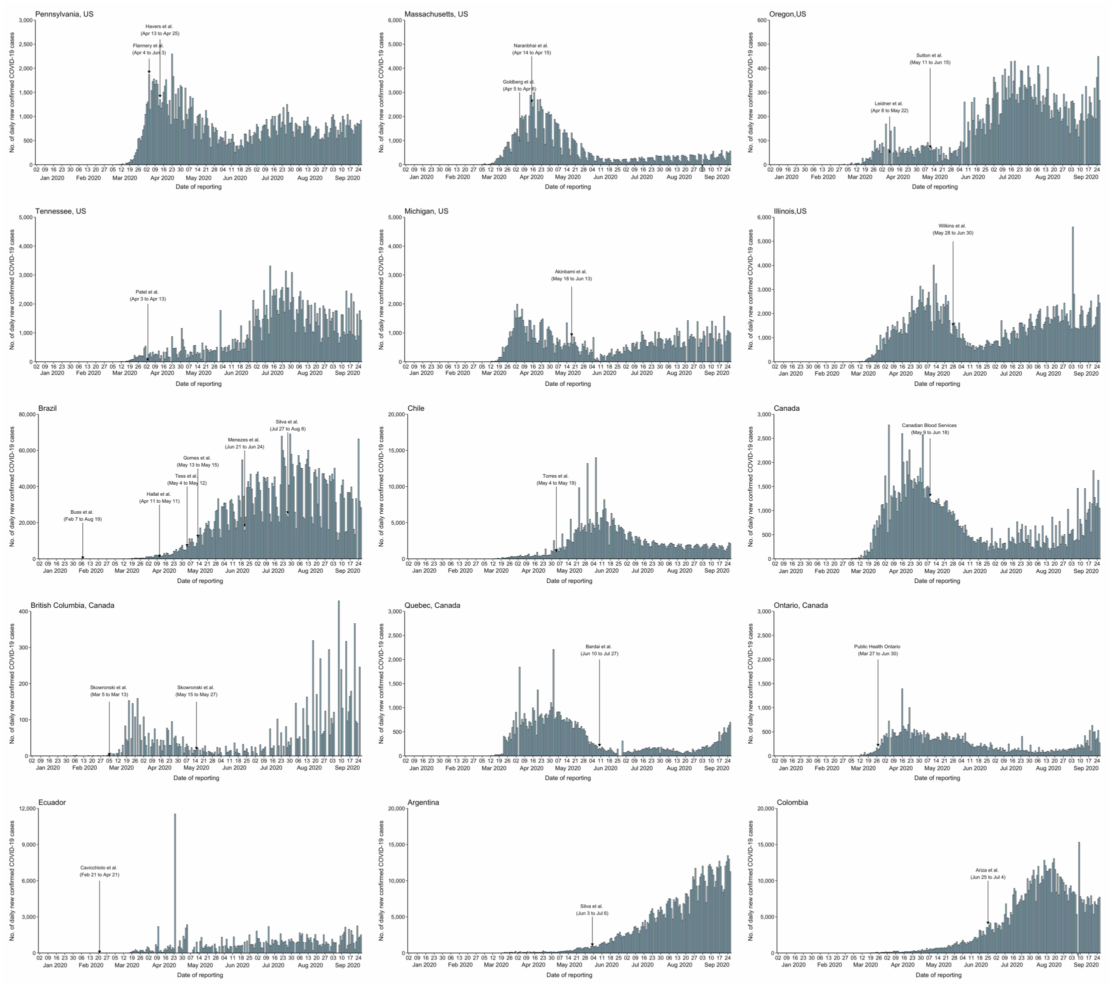
Pennsylvania, US
Havers et al. (Apr 13 to Apr 25)
Flannery et al. (Apr 4 to Jun 3)
Massachusetts, US
Naranbhai et al. (Apr 14 to Apr 15)
Goldberg et al. (Apr 5 to Apr 6)
Oregon,US
Sutton et al. (May 11 to Jun 15)
Leidner et al. (Apr 8 to May 22)
Tennessee, US
Patel et al. (Apr 3 to Apr 13)
Michigan, US
Akinbami et al. (May 18 to Jun 13)
Illinois,US
Wilkins et al. (May 28 to Jun 30)
Brazil
Buss et al. (Feb 7 to Aug 19)
Hallal et al. (Apr 11 to May 11)
Tess et al. (May 4 to May 12)
Gomes et al. (May 13 to May 15)
Menezes et al. (Jun 21 to Jun 24)
Silva et al. (Jul 27 to Aug 8)
Chile
Torres et al. (May 4 to May 19)
Canada
Canadian Blood Services (May 9 to Jun 18)
British Columbia, Canada
Skowronski et al. (Mar 5 to Mar 13)
Skowronski et al. (May 15 to May 27)
Quebec, Canada
Bardai et al. (Jun 10 to Jul 27)
Ontario, Canada
Public Health Ontario (Mar 27 to Jun 30)
Ecuador
Cavicchiolo et al. (Feb 21 to Apr 21)
Argentina
Silva et al. (Jun 3 to Jul 6)
Colombia
Ariza et al. (Jun 25 to Jul 4)
No. of daily new confirmed COVID-19 cases
Date of reporting
Jan 2020 Feb 2020 Mar 2020 Apr 2020 May 2020 Jun 2020 Jul 2020 Aug 2020 Sep 2020

#### Appendix Figure 4. The starting sampling date for each serological study included in this meta-analysis in Eastern Mediterranean Region

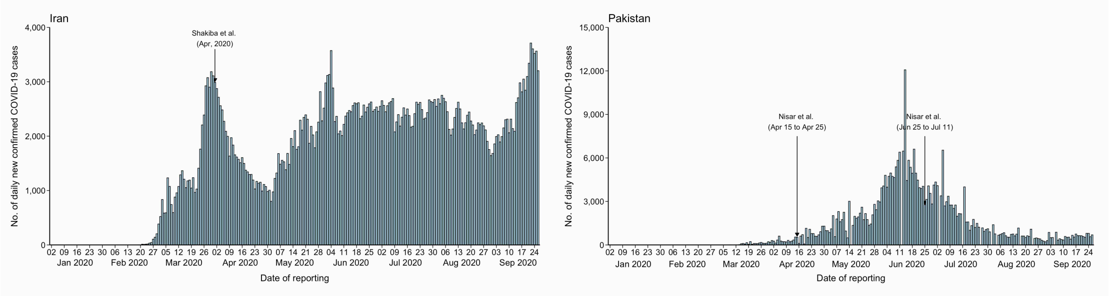

### Appendix Figure 5. The starting sampling date for each serological study included in this meta-analysis in European Region

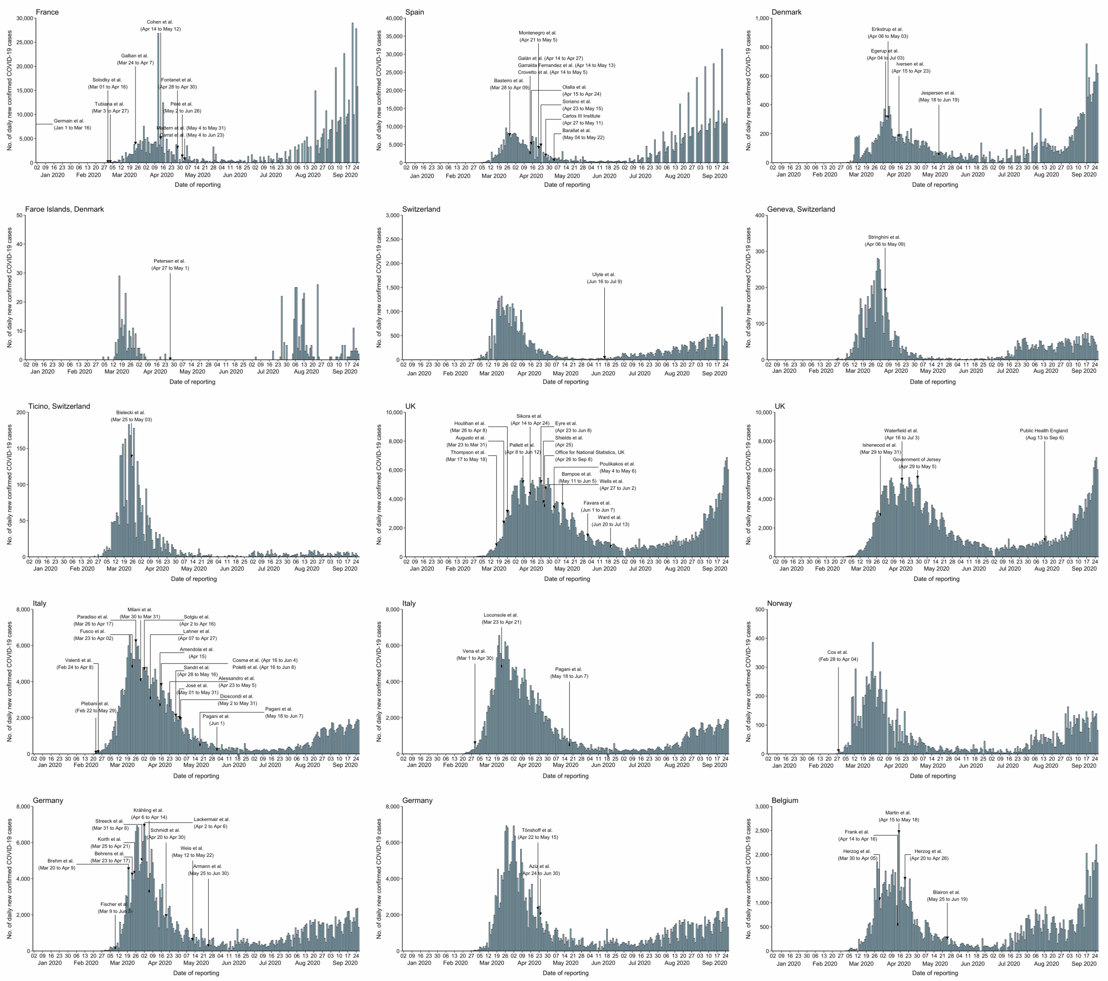

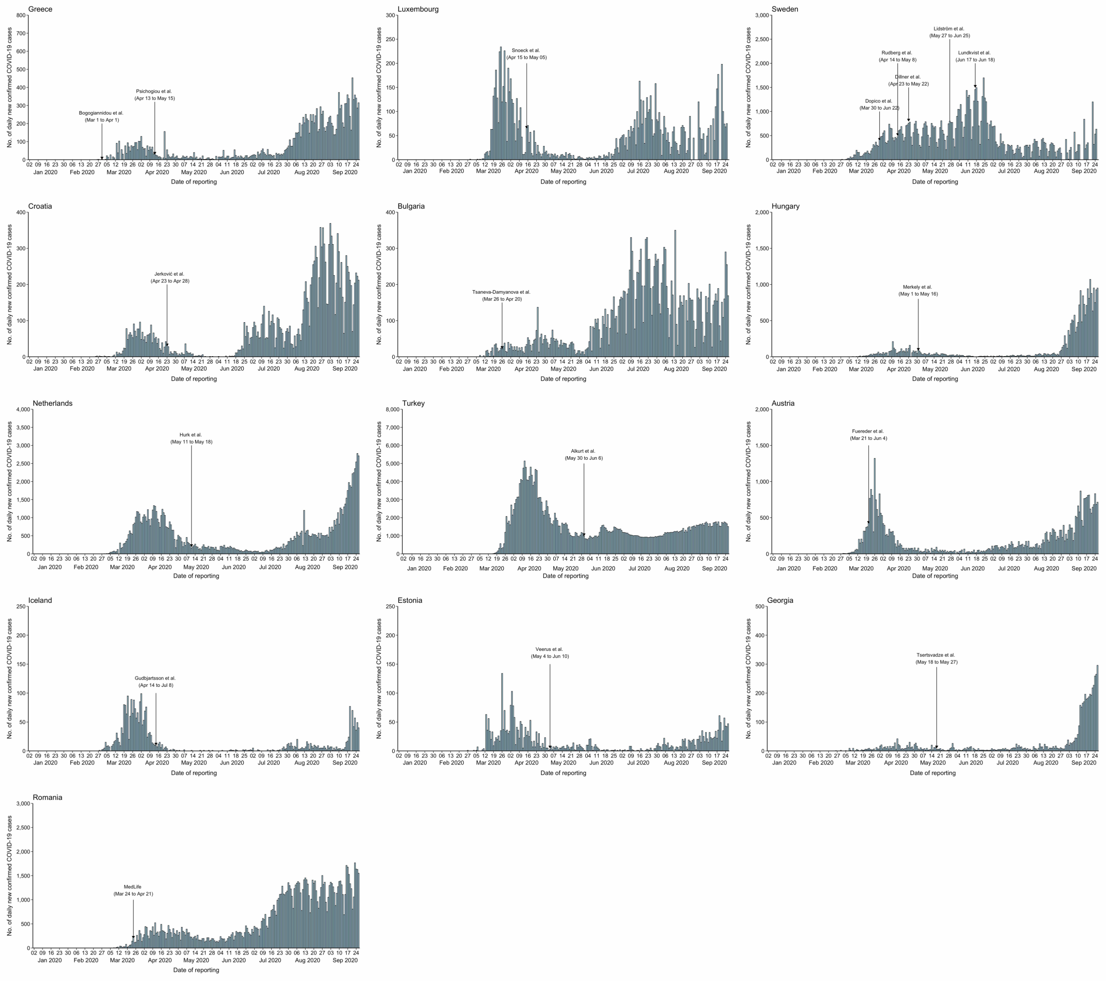
Greece
Bogogiannidou et al.
(Mar 1 to Apr 1)
Psichogiou et al.
(Apr 13 to May 15)
Luxembourg
Snoeck et al.
(Apr 15 to May 05)
Sweden
Dopico et al.
(Mar 30 to Jun 22)
Rudberg et al.
(Apr 14 to May 8)
Dillner et al.
(Apr 23 to May 22)
Lidström et al.
(May 27 to Jun 25)
Lundkvist et al.
(Jun 17 to Jun 18)
Croatia
Jerković et al.
(Apr 23 to Apr 28)
Bulgaria
Tsaneva-Damyanova et al.
(Mar 26 to Apr 20)
Hungary
Merkely et al.
(May 1 to May 16)
Netherlands
Hurk et al.
(May 11 to May 18)
Turkey
Alkurt et al.
(May 30 to Jun 6)
Austria
Fuereder et al.
(Mar 21 to Jun 4)
Iceland
Gudbjartsson et al.
(Apr 14 to Jul 8)
Estonia
Veerus et al.
(May 4 to Jun 10)
Georgia
Tsertsvadze et al.
(May 18 to May 27)
Romania
MedLife
(Mar 24 to Apr 21)
No. of daily new confirmed COVID-19 cases
Date of reporting
Jan 2020
Feb 2020
Mar 2020
Apr 2020
May 2020
Jun 2020
Jul 2020
Aug 2020
Sep 2020

### Appendix Figure 6. The starting sampling date for each serological study included in this meta-analysis in South-East Asia Region

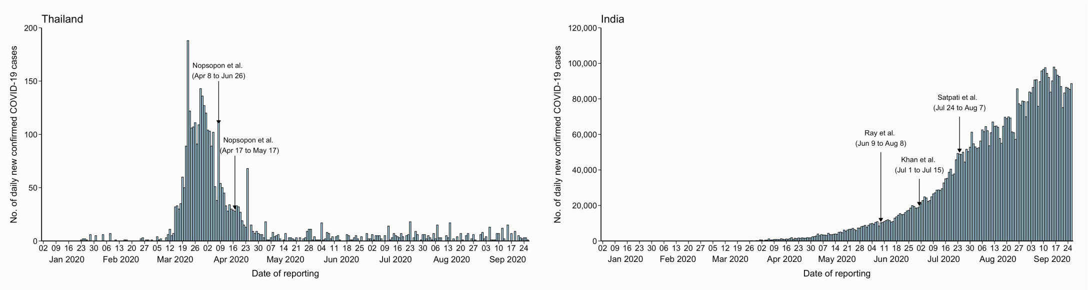

### Appendix Figure 7. The starting sampling date for each serological study included in this meta-analysis in Western Pacific Region

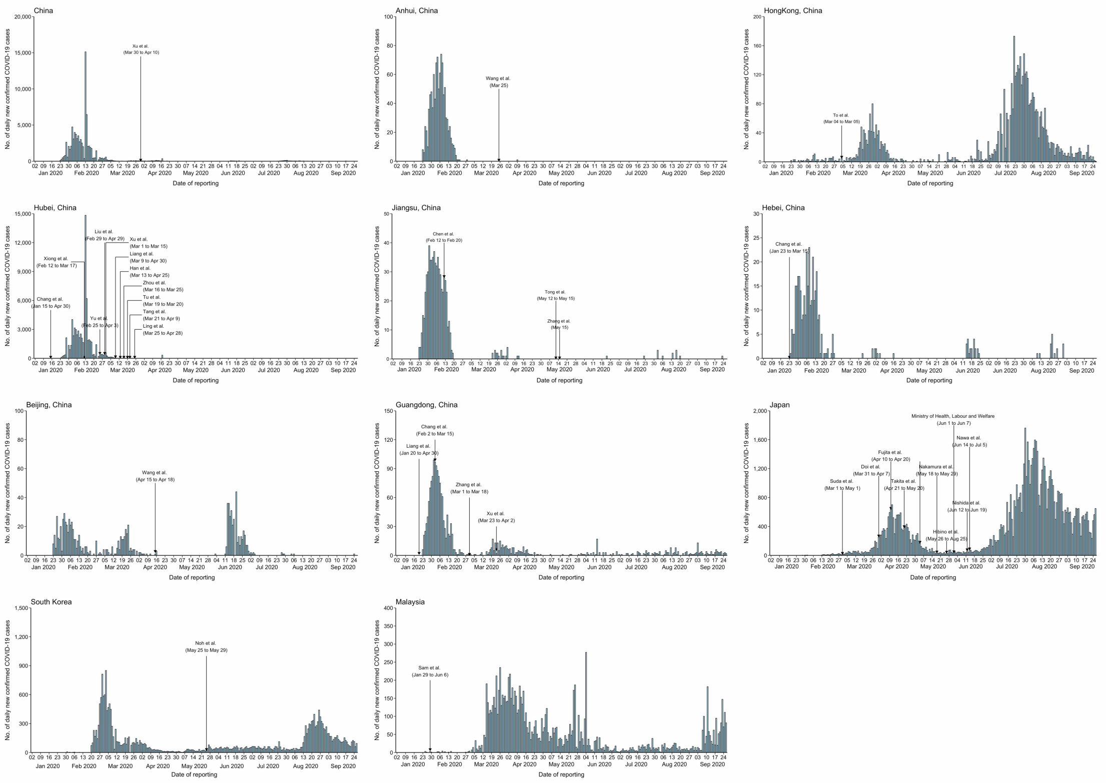

**Appendix Figure 8. The proportion of reported cases that occurred in each area by 2 weeks before the middle time point of each population-based serosurvey**

We calculate the proportion of reported COVID-19 cases among all cases up to Sept 25 that occurred in each area with available epidemiological data among representative population-based serosurveys.

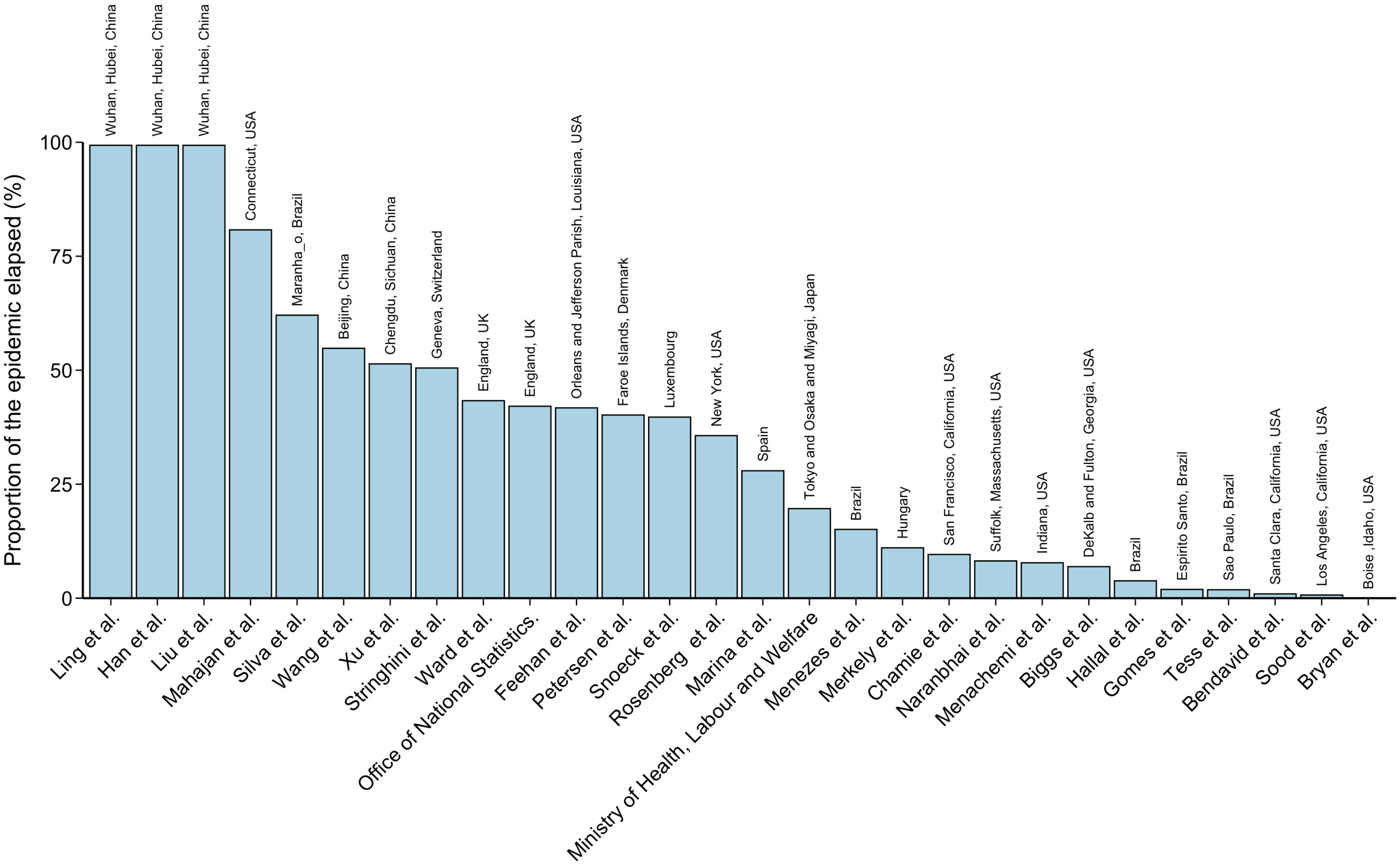
Proportion of the epidemic elapsed (%)
100
75
50
25
0
Wuhan, Hubei, China
Wuhan, Hubei, China
Wuhan, Hubei, China
Connecticut, USA
Maranha_o, Brazil
Beijing, China
Chengdu, Sichuan, China
Geneva, Switzerland
England, UK
England, UK
Orleans and Jefferson Parish, Louisiana, USA
Faroe Islands, Denmark
Luxembourg
New York, USA
Spain
Tokyo and Osaka and Miyagi, Japan
Brazil
Hungary
San Francisco, California, USA
Suffolk, Massachusetts, USA
Indiana, USA
DeKalb and Fulton, Georgia, USA
Brazil
Espirito Santo, Brazil
Sao Paulo, Brazil
Santa Clara, California, USA
Los Angeles, California, USA
Boise ,Idaho, USA
Ling et al.
Han et al.
Liu et al.
Mahajan et al.
Silva et al.
Wang et al.
Xu et al.
Stringhini et al.
Ward et al.
Office of National Statistics.
Feehan et al.
Petersen et al.
Snoeck et al.
Rosenberg et al.
Marina et al.
Ministry of Health, Labour and Welfare
Menezes et al.
Merkely et al.
Chamie et al.
Naranbhai et al.
Menachemi et al.
Biggs et al.
Hallal et al.
Gomes et al.
Tess et al.
Bendavid et al.
Sood et al.
Bryan et al.

**Appendix Figure 9. Geographical distribution of SARS-CoV-2 serosurveys in humans by study populations, December 2019-September 2020.**

(A) Serological studies in the whole world. (B) Serological studies in Europe. The color of the map indicates the cumulative incidence of reported cases with darker colors representing higher values.

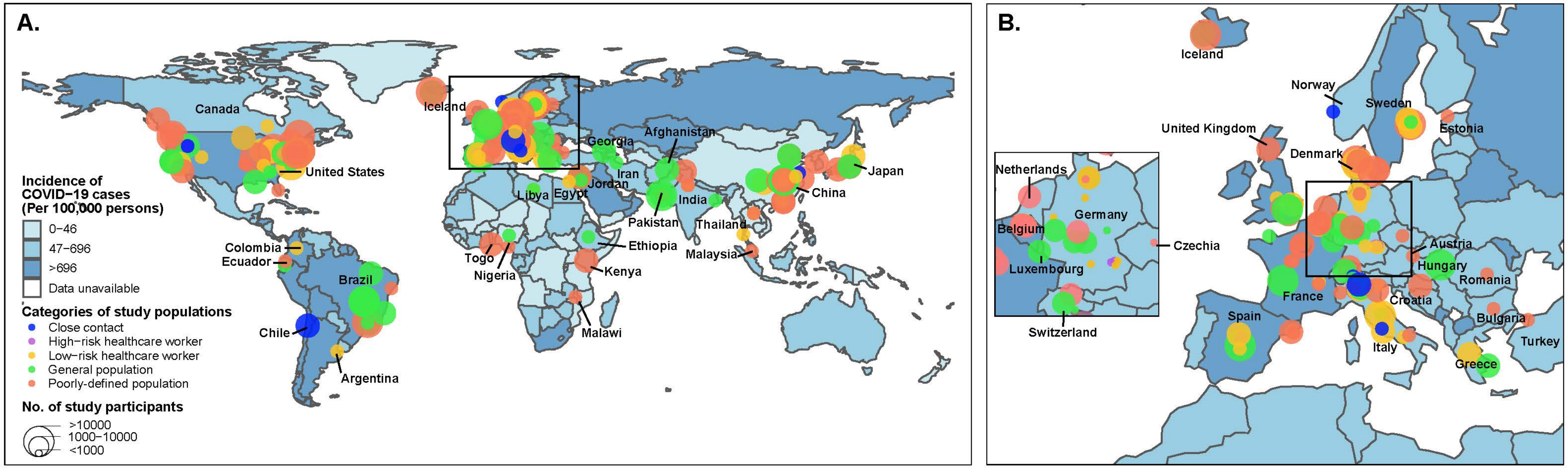

**Appendix Figure 10. Estimated seroprevalence of antibodies to SARS-CoV-2 among grade A and grade B studies involving general populations by age and sex**

**(A)** Seroprevalence of infections with SARS-CoV-2 by age groups; **(B)** Seroprevalence of infections with SARS-CoV-2 by age groups in the Region of the Americas; **(C)** Seroprevalence of infections with SARS-CoV-2 by age groups in the European region; **(D)** Seroprevalence of infections with SARS-CoV-2 by sex; **(E)** Seroprevalence of infections with SARS-CoV-2 by sex in the African region; **(F)** Seroprevalence of infections with SARS-CoV-2 by sex in the Region of the Americas; **(G)** Seroprevalence of infections with SARS-CoV-2 by sex in the European region; (H) Seroprevalence of infections with SARS-CoV-2 by sex in the Western Pacific region;

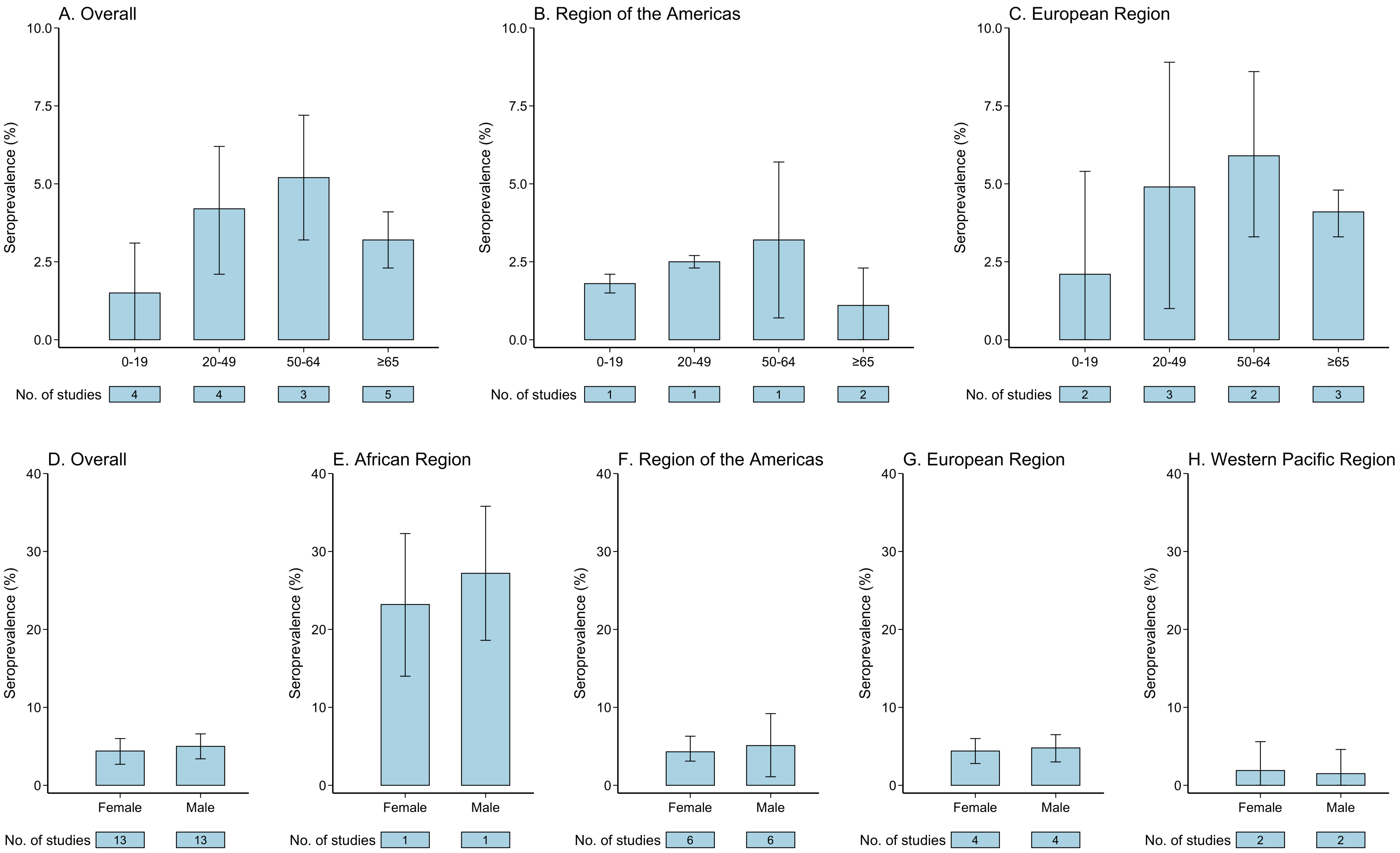
A. Overall
B. Region of the Americas
C. European Region
Seroprevalence (%)
10.0
7.5
5.0
2.5
0.0
0-19
20-49
50-64
≥65
No. of studies
D. Overall
E. African Region
F. Region of the Americas
G. European Region
H. Western Pacific Region
40
30
20
10
0
Female
Male

**Appendix Figure 11. Regression analysis between seroprevalence and local cumulative incidence among grade A and grade B studies involving general populations**

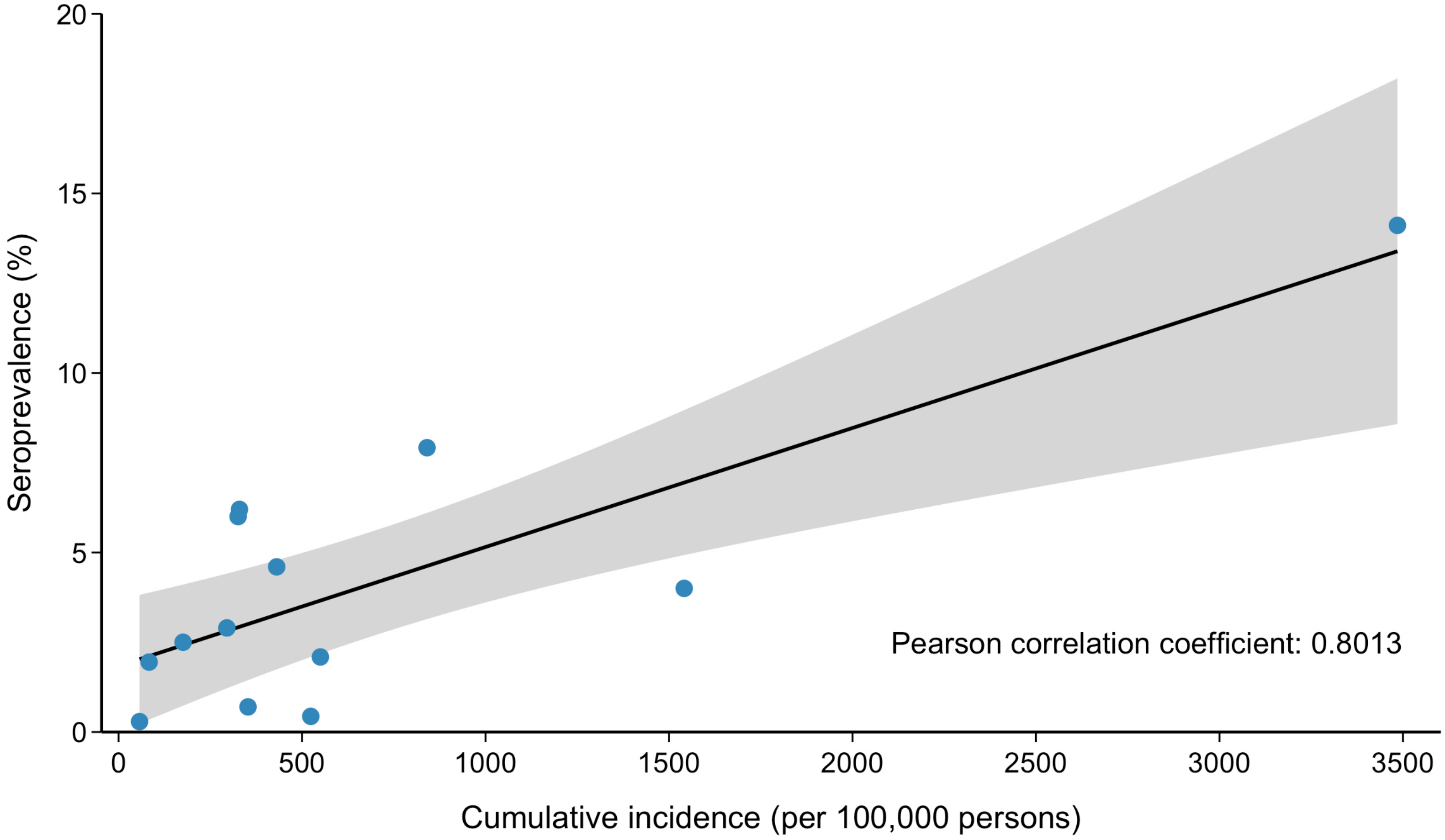
Seroprevalence (%)
20
15
10
5
0
0
500
1000
1500
2000
2500
3000
3500
Cumulative incidence (per 100,000 persons)
Pearson correlation coefficient: 0.8013

### Appendix Figure 12. Estimated seroprevalence by WHO regions and study populations among all 230 studies

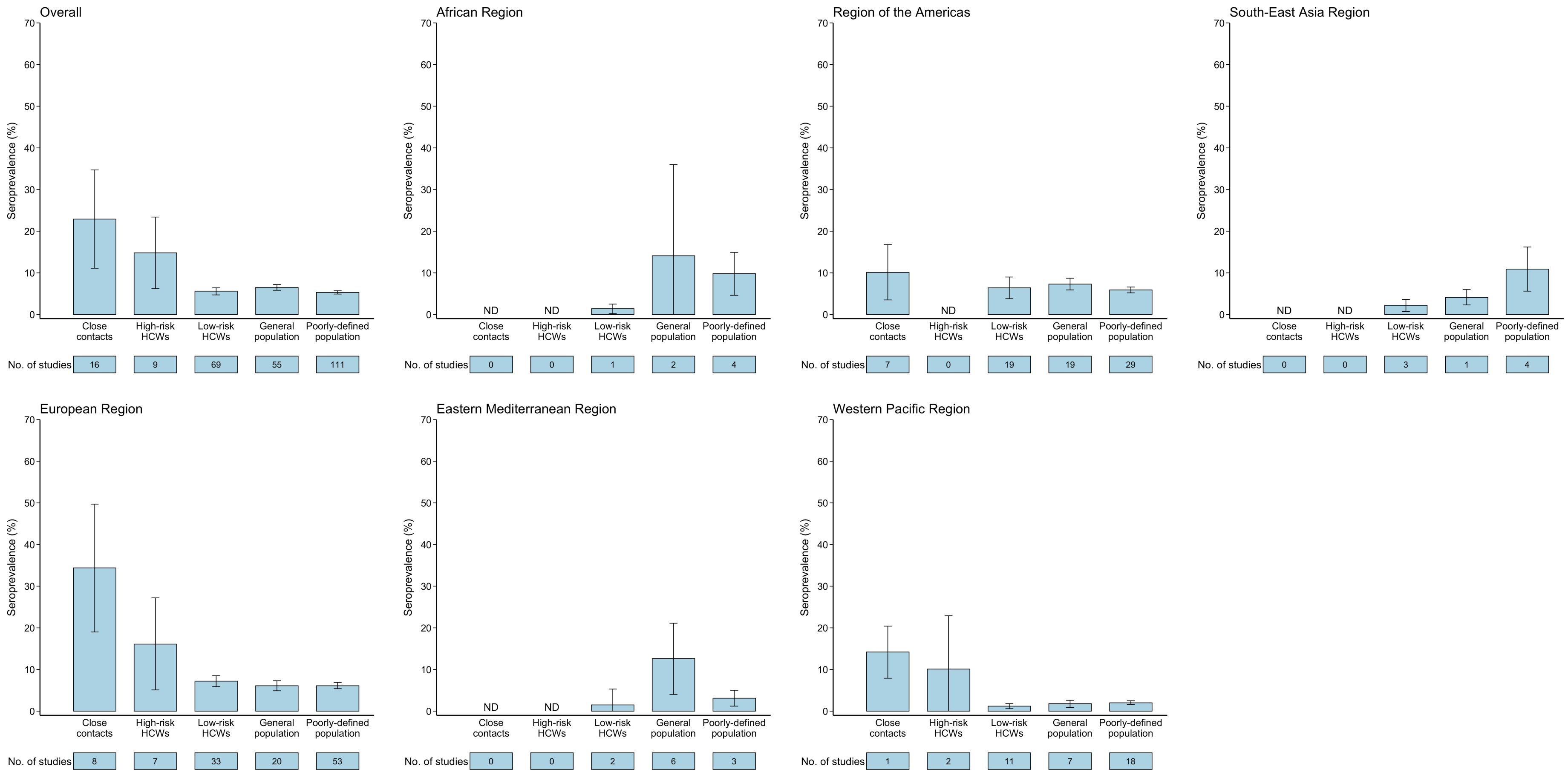
